## Supplemental Text for "Cost-effectiveness of sleeping sickness elimination campaigns in five settings of the Democratic Republic of Congo"

#### Contents

**S1 Supplementary methods** **S2**
S1.1 Locations . . . S2
S1.2 Models . . . S3
S1.3 Health outcomes denominated as disability-adjusted life-years (DALYs) . . . S8
S1.4 Cost functions . . . S10
S1.5 Cost-effectiveness analysis . . . S22
S1.6 Incremental cost-effectiveness ratios, dominance, and weak dominance . . . S24
S1.7 Sensitivity Analysis . . . S25

**S2 Glossary of technical terms** **S26**

**References for S1 & S2** **S27**

**S3 Supplementary results** **S29**
S3.1 Supplemental outcome summaries . . . S30
S3.2 Supplemental cost summaries . . . S30
S3.3 Supplementary cost-effectiveness acceptability curves . . . S39
S3.4 Sensitivity analysis: time-horizon and time discounting . . . S41
S3.5 Sensitivity analysis: vector control . . . S45

**S4 PRIME-NTD criteria** **S48**

**S5 CHEERS checklist** **S49**

**S6 Parameter Glossary** **S53**
S6.1 Organization of parameters . . . S55
S6.2 Principles for parameterization . . . S55
S6.3 Summary of health outcome parameters . . . S56
S6.4 Summary of cost parameters . . . S61
S6.5 Notes on effect parameters . . . S65
S6.6 Notes on cost parameters . . . S79

**References for Parameter Glossary** **S87**

### S1 Supplementary methods

#### S1.1 Locations

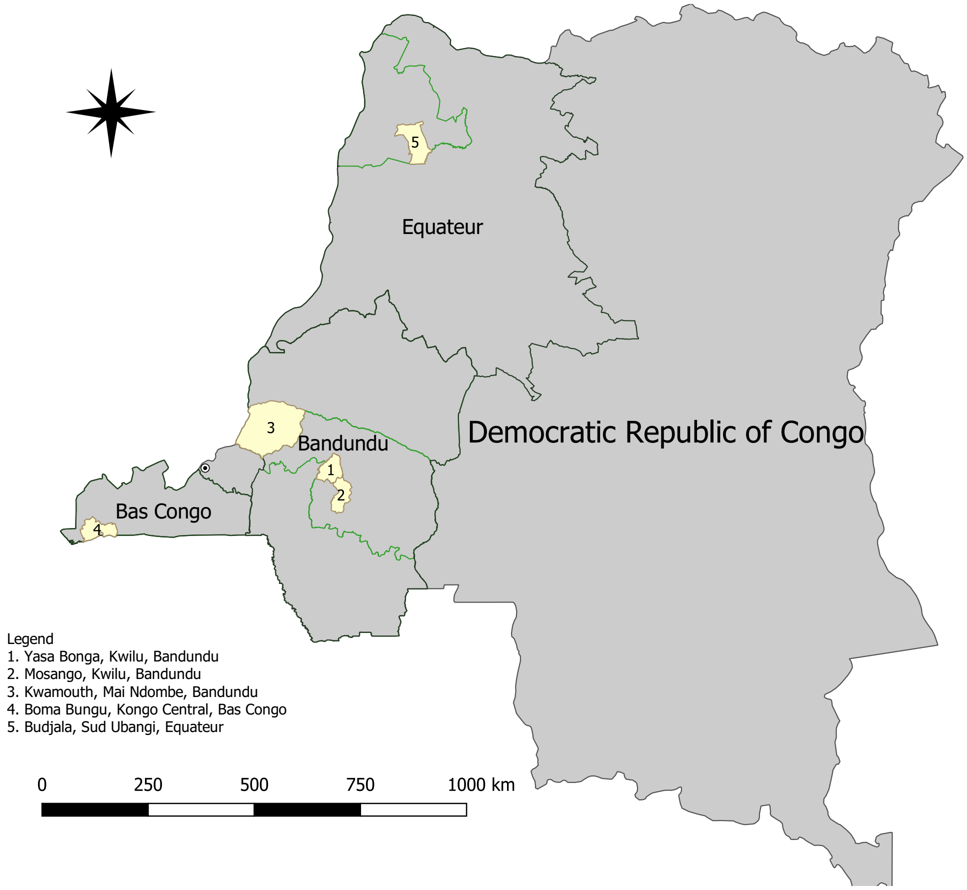


Figure S1: Locations of the specific health zones considered in this study are shown in yellow. New provincial boundaries are denoted in green, and former provincial designations are denoted in black.

#### S1.2 Models

##### S1.2.1 Transmission model

The epidemiological model [1, 2], as described in the methods of the main manuscript, assumes that only low-risk people participate in AS, and high-risk individuals who become diseased are only detected in PS. The proportion of the population considered low-risk is estimated during the fitting process [1] based on historic data (2000–2016). Previous modelling exercises have demonstrated that this is a reasonable modelling assumption based on observed case trends in some parts of DRC and Chad [3, 4], and this type of heterogeneity is necessary to provide a good match to data.

Trends in case reporting and infection levels from 2018, the final year of the period for which data were available, were simulated under a range of alternative intervention strategies, to which cost-effectiveness analysis could then be applied.

Case detection as well as the time lived with disease for cases that were never detected was simulated assuming that the observed prevalence is a combination of simulated prevalence and diagnostic accuracy. Our model assumes that suspects in AS activities are first screened by the card agglutination test for trypanosomiasis (CATT), although sometimes this could be done by a rapid diagnostic test (RDT) instead. Suspects that are screened in fixed facilities (passive screening) are first tested with RDTs. CATT-positive or RDT-positive suspects then have blood drawn for microscopic examination searching for trypanosomes. For patients who qualify for fexinidazole treatment according to WHO guidelines, no further testing is assumed, and for other suspects a lumbar puncture to stage their disease would be performed to determine whether pentamidine or NECT treatment would be administered.

Elimination of transmission (EOT) is assumed when the underlying transmission (not detected cases) falls below 1 new infection per year. Such a proxy threshold is necessary when using deterministic transmission models which represent average dynamics and asymptote to zero rather than a stochastic model including chance events which can hit zero. The metric of interest in this paper is regional EOT (where activities must remain to prevent re-establishment) rather than eradication because we are not treating the issue of importation of cases [5]. This is also related to the WHO's gHAT goal for 2030 which is global EOT to humans [6, 7].

**Uncertainty.** Uncertainty in fitted epidemiological parameter values is incorporated by drawing from their joint posterior distributions as estimated by adaptive MCMC [1, 2]. Most of the fitted parameters correspond to features of the model that can be expected to vary between health zones: including the proportion of the population at low-risk vs high-risk of infection, the rate at which tsetse bite humans rather than other non-human animals, and the reporting rate by passive screening. By contrast, parameters that correspond to features conserved throughout all health zones are assumed to be fixed at available literature estimates: e.g. human mortality rate, tsetse biting rate, and stage 1 to stage 2 disease progression in humans. As well as the variability among joint posterior samples of the fitted parameters; some uncertainty around the parameters that we assume to be fixed, measurement and recording error, and other variation from unknown sources is partially included by sampling detected cases and deaths around the deterministic model outputs from over-dispersed beta-binomial distributions.

##### S1.2.2 Strategy components

**Active screening**.

Each active screening team consists of 8–9 individuals that travel by truck to a village [8–10]. A team has a screening capacity of about 60,000 people per year although there is formidable variance in the literature (see section S6.5.7).

*Coverage.* Coverage of active screening is simulated at "mean" levels and "max" levels. "Mean AS" indicates that the number of people screened is the mean of the AS for the period 2014–2018 whereas under a "Max AS" strategy, the number of people screened is the maximum screened in any single year between 2000–2018. The simulations for the decision analysis were started on 2020 (as no decision can now be made about activities in 2019). The actual screening numbers are applied in 2017 and 2018 and "Mean AS" is assumed for all strategies in 2019.

*Activities.* AS teams can perform initial screening tests with a CATT test or RDT. For those that are CATT- or RDT-positive, the team then examines lymph node glands and performs microscopy exams, consisting of a blood sample taken and examined via mAECT (mini-anion exchange centrifugation technique).

Traditionally, the stage of disease is then determined by microscopy examination of the cerebro-spinal fluid, which is extracted via lumbar puncture for cases in which trypanosomes are present in the blood. Depending on whether trypanosomes are present in the cerebro-spinal fluid (stage 2) or not (stage 1) patients are referred to the appropriate health center or health district hospital for treatment. In the context of fexinidazole treatment, we do not expect that lumbar punctures will be performed by the active screening team, but rather that patients are referred to a health center that will determine eligibility for fexinidazole treatment. However, fexinidazole has only recently been approved for use and how eligibility takes place and lumbar punctures are administered is to be seen.

We note here that while mini-mobile teams are in operation in selected health zones of DRC, their effectiveness at linking cases to care is still under investigation. The potential implications of strategies that employ mini-mobile teams is therefore beyond the scope of this paper.

**Reactive screening**. Reactive screening (RS) is equivalent to AS, but it occurs after a case has been identified in a health zone where AS had ceased after a three-year period of no cases.

**Passive surveillance**. PS, or detection in fixed health facilities is assumed to take place with RDT diagnostic tests used for initial serological screening, followed by confirmatory testing (possibly at another facility with greater diagnostic capacity). The epidemiological model assumes that people are more likely to be detected through the passive system in stage 2 disease rather than stage 1, although in recent years the time until detection has decreased in some locations. For each health zone we have parameterised the stage 1 and 2 detection rates based on historic data for the health zone and the former province it is contained in [1]. In former Bandundu and Bas Congo provinces, we have simulated improvements in PS as there was a strong signal for this in the data. Unlike AS which is assumed to only identify low-risk cases, we assume any person with symptoms may self-present for passive detection regardless of risk status.

*Coverage.* Coverage of PS was assumed to depend on the number of health centres that can perform a serological confirmation of gHAT. We assumed that health zones were served by the same number of centers that were operational for serological confirmation in 2014, according to a publication by Simarro *et al* ([11]), see table 1. Based on unpublished data by colleagues at the Institute of Tropical Medicine-Antwerp, our assumption was that between 0.003 and 0.01 of the population could be screened for gHAT by each clinic, most of which are expected to be gHAT-negative (see section S6.5.3). We note that the Report of the third WHO stakeholder's meeting on gHAT elimination, indicated that 437,402 patients were screened in 514 centers in 2017. We do not know the source population in the areas where the population was screened, but it indicates that approximately 851 patients were screened per center, in line with our estimates (see sections S1.4.2 and S6.5.3).

*Activities.* A selection of fixed health centers can perform screening for gHAT in each health zone. Generally, these centers perform screening with RDT tests rather than CATT because CATT kits (of fifty tests) spoil within a week if unused. For suspects that are RDT-positive, the health worker examines lymph node glands and performs microscopy exams, consisting of a blood sample taken and examined via mAECT (mini-anion exchange centrifugation technique).

Traditionally, the stage of disease is determined by microscopy examination of the cerebro-spinal fluid, which is extracted via lumbar puncture for cases in which trypanosomes are present in the blood. Depending on whether trypanosomes are present in the cerebro-spinal fluid (stage 2) or not (stage 1) patients are referred to the appropriate health center or health district hospital for treatment. In the context of fexinidazole treatment, we do not expect that lumbar punctures will be performed at the health center where diagnosis exists, but rather at the treatment center where eligibility for fexinidazole treatment is determined.

We note here that while there are now some health centres that can administer RDT tests and refer patients to health district hospitals (the referral centres) for confirmation and staging (when necessary) we did not evaluate the impact of additional health centres because the overall impact on disease detection is still under investigation [12].

**Vector control**. In all places, deployments are assumed to occur twice per year, based on the frequency at which it is expected that targets are knocked down or washed away [13–15]. However, other characteristics are more location-specific, which we describe below.

First, the locations where activities are ongoing (Yasa Bonga) or are planned (Kwamouth).

- Yasa Bonga: Our default assumption is that vector control must be deployed along 210 kilometers with 60 targets per kilometer with 90% tsetse reduction, which is the reduction that was observed in field studies in this health zone [15].
- Kwamouth: Our default assumption is that vector control must be deployed along 432 kilometers with 20 targets per kilometer, which is planned in this health zone (personal communication: Prof. Steve Torr). We have assumed 80% tsetse reduction, which is conservative compared to what has been observed in other regions [13, 15].

Second, in the locations where activities are neither ongoing or planned:

- In Mosango, Boma Bungu, and Budjala: Our default assumption is that 100 kilometers of riverbank must be protected with uniform distribution of 40 targets per kilometer. Our rationale for choosing 100 km is that the incidence in these areas is rather low, and so a similar location to use as an example is Arua, in Northwestern Uganda, where only about 77 km of riverbank were protected with 20 targets per kilometer [13].

Sensitivity analyses are described in section S1.7.

##### S1.2.3 Treatment model

We combined the case detections from the dynamic model with a model of treatment outcomes. The model was conceived as a probability tree (Fig-1). The probability of any of the outcomes at the tips of the tree (convalescence, death, treatment failure & S2, rescue treatment with NECT – all with and without serious adverse events) were the result of a series of conditional probabilities along the steps of treatment, serious adverse events, and diagnosis of treatment failure.

We assumed the treatment algorithm based on the WHO interim recommendations of 2019 [16].

- **Step 1, Group A. Patients without clinical symptoms of severe gHAT.** These patients might be eligible for fexinidazole treatment according to the following criteria:
  - **Patients under 6 years old or under 20 kg of weight.** There patients are considered ineligible for fexinidazole treatment. We assumed that all patients over 6 years old were also over 20 kg. We made this assumption because little good data on the patient characteristics are available and it made little different in the costs and effects of the current analysis. See Step 2, Group A.
  - **Patients over 6 years old and over 20 kg of weight.** There patients are considered eligible for fexinidazole treatment. See Step 2, Group B. The recommendations also stipulated that a doctor ought to be certain of adherence in the part of the patient in order to prescribe fexinidazole on an outpatient basis, but we have assumed that this is not an issue for simplicity and because it doesn't make a substantial difference in the costs or effects of this particular analysis.
- **Step 1, Group B. Patients with clinical symptoms of severe HAT.** Patients whose clinical assessment would be consistent with severe gHAT (see Annex 1 of the WHO Interim guidelines [16]) would undergo a lumbar puncture to determine the amount of WBC in the cerebro-spinal fluid. If they were to find fewer than 100 WBC per microliter, then the patient would be referred to fexinidazole treatment. In our model, we assume that none of the stage 1 patients will show more than 100 WBC/$\mu$L of CSF as this is a feature of stage 2 disease, and we have assumed that some proportion of stage 2 patients will be in late-stage disease (see tables S1 and S2). Patients with less than 100 WBC per microliter of CSF would be considered for fexinidazole eligibility based on age and weight, just as those patients in Step 1, Group A.
- **Step 2, Group A. Patients ineligible for fexinidazole treatment.** Patients who were either under 6 years old or under 20 kg in weight undergo a lumbar puncture to determine disease stage. In our model, we consider the cost of a lumbar puncture in these patients (see Table S10) but the outcome of the lumbar puncture (stage 1 or stage 2) is determined by the transmission model.
  - The WHO recommendations stipulate the following criteria: if there are under 5 leukocytes (or white blood cells, WBC) per microliter of cerebrospinal fluid (CSF) and no trypanosoma in the CSF, then the patient undergoes pentamidine treatment on an inpatient basis. If there are more than 5 leukocytes (or white blood cells, WBC) per microliter then the patient undergoes NECT treatment on an inpatient basis. However, the stage of disease is a critical output of the transmission model (described in section S1.2.1) and therefore, the outcome of the lumbar puncture (stage 1 or stage 2) is determined by the transmission model. We consider the cost of a lumbar puncture in these patients (see Table S13).
  - Pentamidine treatment would consist of intra-muscular infections for 7 days. We assumed all pentamidine treatment would have to be administered on an in-patient basis [16].
  - NECT (Nifurtimox-eflornithine combination therapy) is administered on an inpatient basis for 10 days. Nifurtimox is administered orally for 10 days, while eflornithine is administered intravenously for 7 days [16].
- **Step 2, Group B. Patients eligible for fexinidazole treatment.** We assumed that patients would be treated on an inpatient basis if they were over 6 years old and between 20 kg and 35 kg in weight, otherwise, they would be treated on an outpatient basis as directly observed therapy. We assumed that treatment would consist of 1800 mg of fexinidazole for four days and 1200 mg of fexinidazole for six additional days.

**Uncertainty** Uncertainty in the treatment and cost parameters is the variance of estimates in the literature (see sections S32 and S36 for a list of all treatment effect and cost parameters respectively).

| Patient Characteristic | Parameterization | Summary |
|---|---|---|
| Under 6 years old | Beta(152.5, 2427.9) | 0.06 (0.05, 0.07) |
| Under 35 kg of weight | Beta(8.3, 359.6) | 0.02 (<0.01, 0.04) |
| Late stage-2 disease | Beta(76.9, 44.9) | 0.63 (0.54, 0.72) |

Table S1: Parameters for treatment eligibility

| Eligibility | Rationale | Summary |
|---|---|---|
| **Stage 1** | | |
| Pentamidine | Under 6 years old (1) | 0.06 (0.05, 0.07) |
| Fexinidazole-inpatient | Over 6 years old but under 35 kg of weight | 0.02 (<0.01, 0.04) |
| Fexinidazole-outpatient | Over 6 years old and over 35 kg of weight | 0.92 (0.90, 0.93) |
| **Stage 2** | | |
| NECT | Under 6 years old or late-stage disease | 0.65 (0.57, 0.73) |
| Fexinidazole-inpatient | Over 6 years old but under 35 kg of weight and early stage-2 disease | <0.01 (<0.01, 0.01) |
| Fexinidazole-outpatient | Over 6 years old, over 35 kg of weight, and early stage-2 disease | 0.34 (0.26, 0.42) |

[1] For simplicity, all patients over 6 years old were assumed to be over 20 kg in weight.

Table S2: Eligibility for treatment

| Treatment | Outcomes | Estimate |
|---|---|---|
| **Stage 1** | | |
| Pentamidine | Cured | 0.05 (0.05, 0.06) |
| | Cured with SAEs | <0.01 (<0.01, <0.01) |
| | Rescue treatment | <0.01 (<0.01, <0.01) |
| | Death | <0.01 (<0.01, <0.01) |
| Fexinidazole - inpatient | Cured | 0.02 (<0.01, 0.04) |
| | Cured with SAEs | <0.01 (<0.01, <0.01) |
| | Rescue treatment | <0.01 (<0.01, <0.01) |
| | Death | <0.01 (<0.01, <0.01) |
| Fexinidazole - outpatient | Cured | 0.89 (0.87, 0.91) |
| | Cured with SAEs | 0.01 (<0.01, 0.02) |
| | Rescue treatment | 0.02 (<0.01, 0.03) |
| | Death | <0.01 (<0.01, <0.01) |
| **All treatments** | **Cured** | **0.97 (0.95, 0.98)** |
| | **Cured with SAEs** | **0.01 (<0.01, 0.03)** |
| | **Rescue treatment** | **0.02 (0.01, 0.03)** |
| | **Death** | **<0.01 (<0.01, <0.01)** |
| **Stage 2** | | |
| NECT | Cured | 0.56 (0.49, 0.64) |
| | Cured with SAEs | 0.06 (0.04, 0.08) |
| | Rescue treatment | 0.03 (0.01, 0.04) |
| | Death | <0.01 (<0.01, <0.01) |
| Fexinidazole - inpatient | Cured | <0.01 (<0.01, 0.01) |
| | Cured with SAEs | <0.01 (<0.01, <0.01) |
| | Rescue treatment | <0.01 (<0.01, <0.01) |
| | Death | <0.01 (<0.01, <0.01) |
| Fexinidazole - outpatient | Cured | 0.33 (0.25, 0.41) |
| | Cured with SAEs | <0.01 (<0.01, <0.01) |
| | Rescue treatment | <0.01 (<0.01, 0.01) |
| | Death | <0.01 (<0.01, <0.01) |
| **All treatments** | **Cured** | **0.90 (0.88, 0.92)** |
| | **Cured with SAEs** | **0.07 (0.05, 0.09)** |
| | **Rescue treatment** | **0.03 (0.02, 0.05)** |
| | **Death** | **<0.01 (<0.01, <0.01)** |

Table S3: Treatments and outcomes distributions for stage 1 and 2 patients, calculated according to the probability tree in 1. SAE: severe adverse events.

#### S1.3 Health outcomes denominated as disability-adjusted life-years (DALYs)

As per recommendations of the Bill and Melinda Gates Foundation's reference case and WHO's guidelines for the conduct of cost-effectiveness analyses, we defined the utility of gHAT interventions in terms of disability-adjusted life-years (DALYs) [17–19]. DALYs were discounted at a rate of 3% per year [17, 19]. We follow established conventions to calculate DALYs and evaluate the estimates in present-day terms (after applying discounting) [17, 19, 20].

##### S1.3.1 Brief overview of disability-adjusted life-years

DALYs are defined as the sum of the years of life lost to disability (illness) (YLD) and the years of life lost by fatal cases (YLL) [18, 19]. QALYs, a metric related to DALYs averted, is not typically used in analysis of diseases in the developing settings.

$$DALY_{\text{strategy}} = \sum_i YLL_i + YLD_i$$

Three economic notions are integrated into these terms: time-discounting (with a yearly discount rate $r$), relative utility (via disability weights, which in the context of HAT must be conditional on the stage of disease), and and the average life-expectancy of the population in the analysis.

##### S1.3.2 Years of life lost (YLL)

The years of life lost are the years that the individual $i$ would have been expected to live less the age that the individual died in. We do not use the life-table method of accounting (which would take into account that the life-expectancy is actually conditional on the current age of the patient.) Our method is simple yet, we believe, sufficiently accurate because the average age of death of gHAT is as an adult, when life expectancy does not fluctuate appreciably. We do not adjust for the differences in life-expectancy by gender because this is not common practice, and the difference in DALYs averted would be even smaller after time-discounting is taken into account.

$$YLL_i^{\text{undiscounted}} := \text{life expectancy} - \text{age at death}_i$$

To calculate discounted values, we must calculate the discounted years of life lost in terms of the year in which the death occurs, and then adjust the years lost to present day terms (by discounting a second time):

$$YLL_i^{\text{discounted}} := \frac{1}{(1+r)^{(\text{year of death}_i - \text{present year})}} \sum_{k=1}^{YLL_i^{\text{undiscounted}}} \frac{1}{(1+r)^k}$$

For deaths that occur in the future, that means that we must discount future years of life lost to the individual at the time of death, and then we must apply the discounting rate a second time to value their death in present-day terms. For example, suppose the discounting rate is 3%, the individual loses 20 years of life at death in 2025, and the present day is 2014. In that case, that individual loses $20/(1+0.03)^{20} = 11.07$ in 2025, but then we value the years lost in present-day terms (in 2014) by applying a discount rate again: $11.07/(1+0.03)^{11} = 8$. So an individual whose life is spared from death in 2025 and lives from 2025–2045, only accrues 8 years of "DALYs averted" from the perspective of the decision-maker considering prevention options in 2014. If, on the contrary, another intervention saves a person's life in 2014 and the person is estimated to live 20 more years, that person accrues 11.07 years of "DALYs averted" from the perspective of the decision-maker considering prevention options in 2014. In this way, our calculation of DALYs averted shows a preferences for interventions that save lives in younger individuals and sooner rather than later.

We assumed that the mean age of death from gHAT is 26.6 years, according to the limited existing data on the age of infected patients (see Supplement S6.5.17. We extracted life expectancy estimates from the World Bank's World Development Index [21]. We do not expect that this is a cause of major uncertainty so we have applied a fixed value. For information on the parameterisation of life-expectancy, see Section S6.5.31.

##### S1.3.3 Years of life lost to disability (YLD)

The years of life lost to disability are calculated as follows. It is important to note that disability and duration are conditioned on the stage for HAT.

$$YLD_i^{\text{undiscounted}} := \sum_{\text{stage}=1}^{2} \text{duration}_{\text{stage}} \times \text{disability weight}_{\text{stage}}$$

If we apply discounting, we must calculate the discounted years of life lost due to disability in terms of the year in which the individual experiences the disability:

$$YLD_i^{\text{discounted}} := \sum_{\text{illness years}_i} \frac{YLD_i^{\text{undiscounted}}}{(1+r)^{(\text{illness years}_i - \text{present year})}}$$

A disability weight, a factor between 0 (perfect health) and 1 (equivalent to death), is applied to years of life lost due to illness (but not to death) to reflect the severity of gHAT relative to other diseases and to death. These weights are calculated via studies that calculate weights for a compendium of common symptoms. We use the disability weights calculated and used by the Global Burden of Disease study [22], as detailed in section S6.5.32 and S6.5.33.

#### S1.4 Cost functions

The total costs can be characterised by the following expression. Each of the elements in this function, however, might have a functional form of its own, as described in the tables for active screening, passive surveillance, vector control and treatment.

$$\text{Total costs} = \sum_{i \in \text{all sub-categories}} \left( U_i \times C_i \right)$$

*i* is the cost sub-category. Where *U* is the unit of cost, which varies depending on the activity, such as people screened, teams deployed, fixed health centres outfitted with tests, etc. *C* is the cost per unit. All costs were denominated in 2018 US$.

##### S1.4.1 Cost functions: active screening

The yearly costs of AS were calculated as a function of two groups of expenses: 1) overhead costs, and 2) the number of screening tests and confirmation tests that are used across all teams within the health zone. Because no synergies with other disease programs are believed to exist, we have employed a full costing method.

- Overhead costs: overhead costs are split between capital costs and recurrent costs to run an active screening team. AS teams serve a "coordination"– a subnational designation of the PNLTHA program that manages a set of health zones. Therefore, a health zone where fewer than 60,000 people are targeted for screening does not necessarily incur larger overhead costs for active screening than a health zone where the coverage is closer to a multiple of the yearly capacity of a team.
  - Capital costs consist of vehicles, medical equipment, energy (solar panels) and training (which occurs once every few years).
  - Recurrent costs consist of management and consumables that are spent on the team: fuel, staff time, etc.
- Costs that scale by population served:
  - CATT tests are scaled up according to the number of people that are screened per year, and a slight mark-up is included to account for wastage of CATT tests.
  - Confirmation tests are counted for all of those who are positive according to the CATT test, both the false positives (which are modelled according to the specificity of the test) and the true positives, which are the outputs of the dynamic model.
  - Lumbar punctures are not depicted as part of the AS costs, but are included as part of the treatment costs. Because many patients are eligible for fexinidazole treatment, which does not require lumbar punctures, we include lumbar puncture costs in the treatment portion of the analysis for those patients that are not eligible for fexinidazole. See section S1.4.4.
- For all costs, there is a PNLTHA mark-up to account for the central management at the national program headquarters. Snijders and colleagues [10] estimate that PNLTHA management equals approximately 15% of costs, and we have made the same assumption.

The parameters for AS are described in section S6.5.1 and the cost parameters are described in section S6.6.1.

| **Item** | **Units ($U$)** | **Cost ($C$)** |
|---|---|---|
| Capital (annualized) | AS coverage per year × Population ÷ Capacity of one AS team | AS capital × (1+PNLTHA mark-up) |
| Recurrent expenses | AS coverage per year × Population ÷ Capacity of one AS team | AS recurrent × (1+PNLTHA mark-up) |
| CATT testing (See Note 1) | AS coverage per year × Population × (1+wastage factors for AS CATT administration) | CATT × (1+delivery mark-up) × (1+PNLTHA markup) |
| Confirmation (microscopy) | (1-CATT specificity) × (AS coverage per year × Population) + Cases S1 detected with AS + Cases S2 detected with AS | Microscopy × (1+PNLTHA markup) |

[1] Ideally, CATT tests would be used for active screening and RDT tests would be used for passive screening because of the high wastage of CATT tests in the context of passive screening settings.

Table S4: Active screening: cost function

Briefly, we describe these parameters here, but they are displayed in more detail in tables S31 and S35.

| Variable Name | Parameterization | Summary |
|---|---:|---:|
| Population | Fixed | See Table 1 |
| AS coverage per year | Fixed | See Table 1 |
| Capacity of one AS team per year | Normal(60,000, 10,000) | 59,918 (40,446, 79,547) |
| Wastage factor for CATT administration in AS context | Beta(8, 92) | 0.08 (0.04, 0.14) |
| CATT specificity | Beta(31, 2) | 0.94 (0.84, 0.99) |
| AS capital costs (annualized) | Gamma(81, 115) | 9,269 (7,322, 11,436) |
| AS recurrent costs (annual) | Gamma(63, 631) | 39,935 (30,927, 50,321) |
| Cost of CATT test | Gamma(23, 0.02) | 0.46 (0.29, 0.66) |
| Cost confirmation (microscopy) | Gamma(8.475, 1.27) | 10.73 (4.80, 18.87) |
| Cost of delivery (markup) | Beta(45, 55) | 0.45 (0.35, 0.55) |
| PNLTHA markup | Uniform(0.1, 0.2) | 0.15 (0.10, 0.20) |

Table S5: Components of active screening costs

The cost for each health zone per year, given the number of people screened per health zone, is therefore:

Table S6: Cost breakdown for active screening activities

| Item | Units ($U$) | Cost per unit ($C$) | Cost per category |
|---|---:|---:|---:|
| **Yasa Bonga, mean coverage** | | | |
| Capital (annualized) | 2.18 (1.59, 3.13) | 10,656 (8,376, 13,211) | 23,169 (15,544, 34,958) |
| Recurrent expenses | 2.18 (1.59, 3.13) | 45,908 (35,231, 58,185) | 99,818 (65,722, 151,683) |
| CATT testing | 136,588 (130,945, 144,295) | 0.53 (0.33, 0.76) | 71,839 (44,798, 104,678) |
| Confirmation (microscopy) | 7,636 (1,018, 19,961) | 12.33 (5.51, 21.78) | 94,093 (10,485, 286,739) |
| **Total** | | | **288,919 (181,210, 487,461)** |
| **Yasa Bonga, additional coverage in max AS** | | | |
| Capital (annualized) | 1.30 (0.95, 1.87) | 10,656 (8,376, 13,211) | 13,820 (9,272, 20,852) |
| Recurrent expenses | 1.30 (0.95, 1.87) | 45,908 (35,231, 58,185) | 59,541 (39,203, 90,478) |
| CATT testing | 81,474 (78,108, 86,070) | 0.53 (0.33, 0.76) | 42,852 (26,722, 62,439) |
| Confirmation (microscopy) | 4,555 (608, 11,907) | 12.33 (5.51, 21.78) | 56,125 (6,254, 171,037) |
| **Total** | | | **172,337 (108,090, 290,766)** |
| **Mosango, mean coverage** | | | |
| Capital (annualized) | 0.73 (0.53, 1.05) | 10,656 (8,376, 13,211) | 7,789 (5,226, 11,752) |
| Recurrent expenses | 0.73 (0.53, 1.05) | 45,908 (35,231, 58,185) | 33,558 (22,095, 50,995) |
| CATT testing | 45,920 (44,023, 48,511) | 0.53 (0.33, 0.76) | 24,152 (15,061, 35,192) |
| Confirmation (microscopy) | 2,567 (342, 6,711) | 12.33 (5.51, 21.78) | 31,633 (3,525, 96,399) |
| **Total** | | | **97,132 (60,922, 163,881)** |
| **Mosango, additional coverage in max AS** | | | |
| Capital (annualized) | 0.56 (0.41, 0.80) | 10,656 (8,376, 13,211) | 5,956 (3,996, 8,987) |
| Recurrent expenses | 0.56 (0.41, 0.80) | 45,908 (35,231, 58,185) | 25,662 (16,896, 38,996) |

Table S6: Cost breakdown for active screening activities *(continued)*

| Item | Units ($U$) | Cost per unit ($C$) | Cost per category |
|---|---|---|---|
| CATT testing | 35,115 (33,664, 37,096) | 0.53 (0.33, 0.76) | 18,469 (11,517, 26,911) |
| Confirmation (microscopy) | 1,963 (262, 5,132) | 12.33 (5.51, 21.78) | 24,190 (2,696, 73,717) |
| **Total** | | | **74,277 (46,587, 125,320)** |
| **Kwamouth, mean coverage** | | | |
| Capital (annualized) | 1.08 (0.79, 1.55) | 10,656 (8,376, 13,211) | 11,519 (7,728, 17,380) |
| Recurrent expenses | 1.08 (0.79, 1.55) | 45,908 (35,231, 58,185) | 49,628 (32,676, 75,415) |
| CATT testing | 67,910 (65,104, 71,741) | 0.53 (0.33, 0.76) | 35,718 (22,273, 52,044) |
| Confirmation (microscopy) | 3,797 (506, 9,924) | 12.33 (5.51, 21.78) | 46,782 (5,213, 142,563) |
| **Total** | | | **143,647 (90,096, 242,359)** |
| **Kwamouth, additional coverage in max AS** | | | |
| Capital (annualized) | 0.47 (0.35, 0.68) | 10,656 (8,376, 13,211) | 5,040 (3,381, 7,604) |
| Recurrent expenses | 0.47 (0.35, 0.68) | 45,908 (35,231, 58,185) | 21,712 (14,296, 32,994) |
| CATT testing | 29,711 (28,483, 31,387) | 0.53 (0.33, 0.76) | 15,626 (9,745, 22,769) |
| Confirmation (microscopy) | 1,661 (222, 4,342) | 12.33 (5.51, 21.78) | 20,467 (2,281, 62,371) |
| **Total** | | | **62,845 (39,417, 106,032)** |
| **Boma Bungu, mean coverage** | | | |
| Capital (annualized) | 0.11 (0.08, 0.15) | 10,656 (8,376, 13,211) | 1,134 (761, 1,710) |
| Recurrent expenses | 0.11 (0.08, 0.15) | 45,908 (35,231, 58,185) | 4,884 (3,216, 7,422) |
| CATT testing | 6,683 (6,407, 7,060) | 0.53 (0.33, 0.76) | 3,515 (2,192, 5,122) |
| Confirmation (microscopy) | 374 (50, 977) | 12.33 (5.51, 21.78) | 4,604 (513, 14,030) |
| **Total** | | | **14,136 (8,866, 23,851)** |
| **Boma Bungu, additional coverage in max AS** | | | |
| Capital (annualized) | 0.32 (0.24, 0.46) | 10,656 (8,376, 13,211) | 3,432 (2,303, 5,179) |
| Recurrent expenses | 0.32 (0.24, 0.46) | 45,908 (35,231, 58,185) | 14,788 (9,736, 22,471) |
| CATT testing | 20,235 (19,399, 21,377) | 0.53 (0.33, 0.76) | 10,643 (6,637, 15,507) |
| Confirmation (microscopy) | 1,131 (151, 2,957) | 12.33 (5.51, 21.78) | 13,939 (1,553, 42,479) |
| **Total** | | | **42,802 (26,845, 72,215)** |
| **Budjala, mean coverage** | | | |
| Capital (annualized) | <0.01 (<0.01, 0.01) | 10,656 (8,376, 13,211) | 100 (67, 151) |
| Recurrent expenses | <0.01 (<0.01, 0.01) | 45,908 (35,231, 58,185) | 432 (284, 656) |
| CATT testing | 591 (566, 624) | 0.53 (0.33, 0.76) | 311 (194, 453) |
| Confirmation (microscopy) | 33 (4, 86) | 12.33 (5.51, 21.78) | 407 (45, 1,240) |
| **Total** | | | **1,249 (784, 2,108)** |
| **Budjala, additional coverage in max AS** | | | |
| Capital (annualized) | 0.82 (0.60, 1.17) | 10,656 (8,376, 13,211) | 8,698 (5,835, 13,123) |

Table S6: Cost breakdown for active screening activities *(continued)*

| Item | Units ($U$) | Cost per unit ($C$) | Cost per category |
|---|---|---|---|
| Recurrent expenses | 0.82 (0.60, 1.17) | 45,908 (35,231, 58,185) | 37,472 (24,672, 56,943) |
| CATT testing | 51,276 (49,157, 54,169) | 0.53 (0.33, 0.76) | 26,969 (16,818, 39,296) |
| Confirmation (microscopy) | 2,867 (382, 7,494) | 12.33 (5.51, 21.78) | 35,323 (3,936, 107,643) |
| **Total** | | | **108,461 (68,027, 182,995)** |

##### S1.4.2 Cost functions: passive surveillance (screening at fixed health posts)

The yearly costs of PS were calculated as a function of two groups of expenses: 1) overhead costs, and 2) the number of consultations and screening and confirmation tests that are done in a health zone.

- Overhead costs: overhead costs are split between capital costs and recurrent costs to equip a health center to perform serological confirmation for HAT.
  - Capital costs consist of medical equipment, energy (solar panels) and training (which occurs periodically every few years). These costs are scaled by the number of facilities that can perform serological confirmation.
  - Recurrent costs consist of management: health district and provincial management and supervision. Only one management 'unit' was assumed per health zone, irrespective of the number of health centers that are capable of screening.
- Costs that scale by population served:
  - RDT tests are scaled up according to the number of people that are screened per year, and a slight mark-up is included to account for wastage of tests.
  - Confirmation tests are counted for all of those who are positive according to the RDT test: both the false positives (which are modelled as a factor equal to the specificity of the test) and the true positives, which are the outputs of the dynamic model.
  - Lumbar punctures are not depicted as part of the passive surveillance diagnosis costs, but are included as part of the treatment costs. Because many patients are eligible for fexinidazole treatment, which does not require lumbar punctures, we include lumbar puncture costs in the treatment portion of the analysis for those patients that are not eligible for fexinidazole. See section S1.4.4
- For all costs, there is a PNLTHA mark-up to account for the central management at the national program headquarters. Snijders and colleagues ([10]) estimate that PNLTHA management equals approximately 15% of costs, and we have made the same assumption.

| Item | Units ($U$) | Cost ($C$) |
|---|---|---|
| Capital (annualized) | Number of facilities capable of PS within the health zone | Capital costs (clinic) × (1+PNLTHA mark-up) |
| District management | Per district | District management cost × (1+PNLTHA mark-up) |
| Consultation | PS coverage per year per clinic × Clinics in the health zone × Population | Consultation cost × (1+PNLTHA markup) |
| RDT testing (See Note 1) | PS coverage per year per clinic × Clinics in the health zone × Population | RDT × (1+delivery mark-up) × (1+PNLTHA markup) |
| Confirmation (microscopy) | (1-RDT specificity) × (PS coverage per year per clinic × Clinics in the health zone × Population) + Cases S1 detected with PS + Cases S2 detected with PS | Microscopy × (1+PNLTHA markup) |

[1] Ideally, CATT tests would be used for active screening and RDT tests would be used for passive screening because of the high wastage of CATT tests in the context of passive screening settings.

Table S7: Passive screening: cost function

Briefly, we describe these parameters here, but they are displayed in more detail in tables S31 and S35.

| Variable Name | Parameterization | Summary |
|---|---|---|
| Population | Fixed | See Table 1 |
| PS facilities | Fixed | See Table 1 |
| PS coverage per facility | Beta(36, 7927) | 0.007 (0.004, 0.010) |
| Wastage factor for RDT administration in PS context | Beta(1, 99) | 0.01 (<0.01, 0.04) |
| RDT specificity | Beta(226, 31) | 0.88 (0.84, 0.92) |
| Capital costs (annualized) | Gamma(8.475, 210) | 1,768 (793, 3,112) |
| Recurrent costs (yearly) | Gamma(8.475, 986) | 8,334 (3,723, 14,869) |
| Cost of consultation | Gamma(1.37, 3.33) | 2.34 (1.41, 3.28) |
| Cost of RDT | Gamma(8.475, 0.19) | 1.61 (0.72, 2.88) |
| Cost confirmation (microscopy) | Gamma(8.475, 1.27) | 10.73 (4.80, 18.87) |
| Cost of delivery (markup) | Beta(45, 55) | 0.45 (0.35, 0.55) |
| PNLTHA markup | Uniform(0.1, 0.2) | 0.15 (0.10, 0.20) |

Table S8: Components of passive screening costs

The cost for each health zone per year, given the number of people screened per health zone and the number of health centres available for PS, is shown in table S9.

| Item | Units ($U$) | Cost per unit ($C$) | Cost per category |
|---|---|---|---|
| **Yasa Bonga** | | | |
| Capital (annualized) | 4 | 2,033 (909, 3,590) | 8,131 (3,635, 14,359) |
| District management | 1 | 9,580 (4,261, 17,063) | 9,580 (4,261, 17,063) |
| Consultation | 5,893 (3,210, 9,310) | 2.69 (1.62, 3.80) | 15,850 (6,904, 29,000) |
| RDTs | 5,893 (3,210, 9,310) | 2.68 (1.21, 4.82) | 15,824 (5,712, 33,225) |
| Confirmation (microscopy) | 711 (355, 1,225) | 12.33 (5.51, 21.78) | 8,768 (2,954, 19,035) |
| **Total** | | | **58,153 (35,668, 90,214)** |
| **Mosango** | | | |
| Capital (annualized) | 1 | 2,033 (909, 3,590) | 2,033 (909, 3,590) |
| District management | 1 | 9,580 (4,261, 17,063) | 9,580 (4,261, 17,063) |
| Consultation | 830 (452, 1,312) | 2.69 (1.62, 3.80) | 2,233 (973, 4,086) |
| RDTs | 830 (452, 1,312) | 2.68 (1.21, 4.82) | 2,230 (805, 4,682) |
| Confirmation (microscopy) | 100 (50, 173) | 12.33 (5.51, 21.78) | 1,235 (416, 2,682) |
| **Total** | | | **17,311 (10,855, 25,836)** |
| **Kwamouth** | | | |
| Capital (annualized) | 5 | 2,033 (909, 3,590) | 10,164 (4,544, 17,949) |
| District management | 1 | 9,580 (4,261, 17,063) | 9,580 (4,261, 17,063) |
| Consultation | 4,349 (2,369, 6,871) | 2.69 (1.62, 3.80) | 11,698 (5,095, 21,402) |
| RDTs | 4,349 (2,369, 6,871) | 2.68 (1.21, 4.82) | 11,678 (4,216, 24,521) |
| Confirmation (microscopy) | 525 (262, 904) | 12.33 (5.51, 21.78) | 6,471 (2,180, 14,048) |
| **Total** | | | **49,590 (31,425, 74,572)** |
| **Boma Bungu** | | | |
| Capital (annualized) | 2 | 2,033 (909, 3,590) | 4,066 (1,818, 7,180) |
| District management | 1 | 9,580 (4,261, 17,063) | 9,580 (4,261, 17,063) |
| Consultation | 1,141 (622, 1,803) | 2.69 (1.62, 3.80) | 3,070 (1,337, 5,617) |
| RDTs | 1,141 (622, 1,803) | 2.68 (1.21, 4.82) | 3,065 (1,106, 6,435) |
| Confirmation (microscopy) | 138 (69, 237) | 12.33 (5.51, 21.78) | 1,698 (572, 3,687) |
| **Total** | | | **21,478 (13,939, 31,301)** |
| **Budjala** | | | |
| Capital (annualized) | 2 | 2,033 (909, 3,590) | 4,066 (1,818, 7,180) |
| District management | 1 | 9,580 (4,261, 17,063) | 9,580 (4,261, 17,063) |
| Consultation | 1,772 (965, 2,799) | 2.69 (1.62, 3.80) | 4,765 (2,076, 8,718) |
| RDTs | 1,772 (965, 2,799) | 2.68 (1.21, 4.82) | 4,757 (1,717, 9,988) |
| Confirmation (microscopy) | 214 (107, 368) | 12.33 (5.51, 21.78) | 2,636 (888, 5,722) |
| **Total** | | | **25,803 (16,759, 37,823)** |

Table S9: Cost breakdown for passive screening activities

##### S1.4.3 Cost functions: vector control

The costs of VC were calculated as a function of two features: 1) the extent of the rivers whereVC is deployed, and 2) the number of targets per kilometer of river where the targets were deployed. Full cost methods were used for the available cost data and that method was thus employed here. Because no synergies with other disease programs are believed to exist, we believe that a full costing method is appropriate.

The costs of entomological surveys (tsetse monitoring), sensitisation of the population (information campaigns), and district management were assumed to scale in relation to the extent of the health zone that where VC is deployed. The materials and labor-time related to target deployment was then considered to scale in relation to the amount of targets deployed.

There are thus far three recent instances of vector control described in the scientific literature: Arua, Uganda ([13, 23]); Mandoul, Chad ([4]); and Yasa Bonga, DRC ([15]). In Arua, 1551 targets were deployed at 20 targets per kilometer, for a total of 77.5 kilometers of riverbank. In Mandoul, 2708 targets were deployed at 60 targets per kilometer, for a total of 45 kilometers of riverbank. In Yasa Bonga, 22,622 targets were deployed over a year throughout two deployments at 55 targets per kilometer (personal communication with colleagues from the Liverpool School of Tropical Medicine (LSTM), June 2020). The standard operating procedures for VC from LSTM say that 40 targets should be deployed per linear kilometer, although the particular landscape of the river ought to be taken into account ([14]).

The extent of the riverbank that is necessary to cover for adequate coverage is determined by the case reports of the previous five years. Because both the extent of riverbank and the density of targets is unknown unless a thorough analysis of the health zone is performed, we have chosen to show extensize sensitivity analyses of the cost-effectiveness of different strategies:

- In Yasa Bonga, we only performed analyses of the costs of 210 km of river bank and 55 targets per kilometer.
- In Kwamouth, the plan is to deploy targets along 432 kilometers of riverbank at approximately 20 targets per kilometer. Although operations have begun, the full deployment has not been performed yet. To speak to concerns of uncertainty, we performed the analyses with 40 targets per kilometer, to understand the resource implications if more ambitious VC operations are necessary to reach a certain goal for tsetse density.

The full extent of our sensitivity analyses are described in section S1.7.

The parameters for VC are described in section S6.5.35 and the VC cost parameters are described in section S6.6.18.

| Item | Units ($U$) | Cost ($C$) |
|---|---:|---:|
| Entomological surveys, sensitization and management | Kilometers of river covered | Cost for entomological surveys, sensitisation and district management per kilometer × (1+PNLTHA markup) |
| Target deployment | Kilometers of river covered × Targets per kilometer × Number of deployments per year | Cost for target deployment per target × (1+PNLTHA markup) |

Table S10: Vector Control: cost function

Briefly, we describe these parameters here, but they are displayed in more detail in tables S34 and S37.

Per year, the simulated costs according to the above formulation and parameters results in the following estimates.

| Variable Name | Parameterization | Summary |
|---|---|---|
| Linear km | Fixed (sensitivity analyses) | 100 or 210 (see note 1) |
| Units per km | Fixed (sensitivity analyses) | 20 or 40 (see note 2) |
| Deployments per year | Fixed | 2 |
| Cost for entomological surveys, sensitisation and district management per kilometer | Gamma(8.475, 14.17) | 120.60 (53.70, 213.63) |
| Cost per target deployment per target | Gamma(8.475, 0.54) | 4.55 (2.02, 8.04) |
| PNLTHA markup | Uniform(0.1, 0.2) | 0.15 (0.10, 0.20) |

[1] Due to Kwamouth's large size and two transmission hotspots, VC is deployed in 432 km of river.
[2] In Yasa Bonga, the current units per km are 60, above the recommendations.

Table S11: Components of vector control costs

| Item | Units ($U$) | Cost ($C$) |
|---|---|---|
| **Yasa Bonga** | | |
| Entomological surveys, sensitization and management | 210 km | 29,119 (13,023, 51,750) |
| Target deployment | 25200 targets | 131,835 (58,625, 233,769) |
| **Total** | | **160,954 (85,049, 265,011)** |
| **Kwamouth (planned): 432 km, 20 targets per kilometer** | | |
| Entomological surveys, sensitization and management | 432 km | 59,902 (26,789, 106,456) |
| Target deployment | 17280 targets | 90,401 (40,200, 160,299) |
| **Total** | | **150,303 (87,033, 232,730)** |
| **Kwamouth (sensitivity analysis): 432 km, 40 targets per kilometer** | | |
| Entomological surveys, sensitization and management | 432 km | 59,902 (26,789, 106,456) |
| Target deployment | 34560 targets | 180,802 (80,400, 320,597) |
| **Total** | | **240,704 (131,663, 386,342)** |
| **Other health zones (default analysis): 100 km, 40 targets per kilometer** | | |
| Entomological surveys, sensitization and management | 100 km | 13,866 (6,201, 24,643) |
| Target deployment | 8000 targets | 41,852 (18,611, 74,212) |
| **Total** | | **55,719 (30,478, 89,431)** |
| **Other health zones (sensitivity analysis 1): 100 km, 20 targets per kilometer** | | |
| Entomological surveys, sensitization and management | 100 km | 13,866 (6,201, 24,643) |
| Target deployment | 4000 targets | 20,926 (9,306, 37,106) |
| **Total** | | **34,792 (20,147, 53,873)** |
| **Other health zones (sensitivity analysis 2): 210 km, 20 targets per kilometer** | | |
| Entomological surveys, sensitization and management | 210 km | 29,119 (13,023, 51,750) |
| Target deployment | 8400 targets | 43,945 (19,542, 77,923) |
| **Total** | | **73,064 (42,308, 113,133)** |
| **Other health zones (sensitivity analysis 3): 210 km, 40 targets per kilometer** | | |
| Entomological surveys, sensitization and management | 210 km | 29,119 (13,023, 51,750) |
| Target deployment | 16800 targets | 87,890 (39,083, 155,846) |
| **Total** | | **117,009 (64,003, 187,805)** |

Table S12: Cost breakdown for vector control activities

##### S1.4.4 Cost functions: treatment

| Item | Units ($U$) | Cost ($C$) |
|---|---|---|
| Doctor's consult | All patients | Outpatient consult × (1+PNLTHA markup) |
| Staging cases (supplies and time); patients ineligible for fexinidazole treatment (see Notes 1-2). | Patients in both stages of disease ineligible for fexinidazole treatment. | Lumbar puncture cost × (1+PNLTHA markup) |
| Pentamidine (see Notes 1-3). | Cases of stage 1 disease detected with AS or PS × proportion of patients ineligible for fexinidazole. | Pentamidine × (1+delivery mark-up) × (1+PNLTHA markup) |
| Inpatient care for stage 1 with pentamidine (see Notes 1-3). | Cases of stage 1 disease detected with AS or PS × proportion of patients ineligible for fexinidazole × length of treatment for pentamidine. | Inpatient cost per day × (1+PNLTHA markup) |
| NECT (see Notes 1-3). | Cases of stage 2 disease detected with AS or PS × proportion of patients ineligible for fexinidazole. | NECT × (1+delivery mark-up) × (1+PNLTHA markup) |
| Inpatient care for stage 2 with NECT (see Notes 1-3). | (Cases stage 2 detected with AS or PS) × proportion of patients ineligible for fexinidazole × length of treatment for NECT. | Inpatient cost per day × (1+PNLTHA markup) |
| Fexinidazole (see Notes 1-2). | Patients in both stages of disease eligible for fexinidazole treatment. | Fexinidazole × (1+delivery mark-up) × (1+PNLTHA markup) |
| Inpatient care for either stage 1 or 2 with fexinidazole (see Notes 1-3) | Patients eligible for fexinidazole on an inpatient basis × length of treatment for fexinidazole. | Inpatient cost per day × (1+PNLTHA markup) |
| Outpatient care for either stage 1 or 2 with fexinidazole (see Notes 1-4). | Patients eligible for outpatient treatment. | Outpatient consult × (1+PNLTHA markup) |
| Treatment for severe adverse events | Patients under each treatment who experience severe adverse events | (Outpatient consult + Inpatient cost per day × length of treatment for severe adverse events) × (1+PNLTHA markup) |

[1] See WHO treatment recommendations for eligibility for fexinidazole. While some patients are eligible for fexinidazole (over 6 years of age and below the severe threshold of disease) the recommendations stipulate inpatient care for some patients due to low weight.
[2] The proportion of patients eligible for fexinidazole was determined as follows: (1-proportion of patients under 6 years of age) × (1-proportion of patients with signs of late stage 2 disease).
[3] The proportion of patients who had fexinidazole treatment on an inpatient basis was determined by multiplying the equation in note 2 with the proportion of patients who were over 35 kg of weight.
[4] Fexinidazole is currently recommended on an outpatient basis as a directly-observed therapy, so we have imputed a cost for the daily administration by a village health worker. For simplicity, we have given this the same value as a regular outpatient visit since it constitutes a small portion of all costs.

Table S13: Treatment: cost function

We show here the components of the costs per case treated depending on the stage and the treatment. The parameters for the above table are available in section S6.6.11 and eligibility distributions are described in table S2.

| Variable.Name | Parameterization | Summary |
|---|---|---|
| Lumbar puncture and laboratory exam - cost | Gamma(2.42, 3.66) | 8.83 (1.41, 22.81) |
| PNLTHA mark-up | Uniform(0.1, 0.2) | 0.15 (0.10, 0.20) |
| Duration of hospital stay for NECT treatment in days | Fixed | 10 |
| Duration of hospital stay fexinidazole for stage 1 or 2 disease in days | Fixed | 10 |
| Duration of severe adverse events in days | Gamma (1.219, 2.377) | 2.90 (0.13, 9.67) |
| Probability of serious adverse events - pentamidine | Beta(1,499) | 0.0026 (0.0002, 0.0083) |
| Probability of serious adverse events - NECT | Beta(11.6,226.4) | 0.10 (0.07, 0.13) |
| Probability of serious adverse events - fexinidazole | Beta(3,261) | 0.01 (<0.01, 0.03) |
| Outpatient consultation - cost | Gamma(1.37,3.33) | 2.34 (1.41, 3.28) |
| Hospital day - cost | Gamma(5.81,0.24) | 1.39 (0.49, 2.72) |
| Course of pentamidine - cost | Fixed | 54 |
| Course of NECT - cost | Fixed | 460 |
| Course of fexinidazole - cost | Fixed | 50 |
| Delivery mark-up | Beta(45,55) | 0.45 (0.35, 0.55) |

Table S14: Parameters for treatment costs

| | Pentamidine | NECT | Fexinidazole - inpatient | Fexinidazole - outpatient |
|---|---|---|---|---|
| Staging | 10.15 (1.61, 26.22) | 10.15 (1.61, 26.22) | 0 | 0 |
| Doctor's consult | 21.52 (12.99, 30.36) | 2.34 (1.41, 3.28) | 2.34 (1.41, 3.28) | 23.40 (14.14, 32.80) |
| Inpatient care | 0 | 13.90 (4.92, 27.19) | 13.90 (4.92, 27.19) | 0 |
| Medicine | 78.31 (73.17, 83.56) | 667.09 (623.26, 711.81) | 72.51 (67.75, 77.37) | 72.51 (67.75, 77.37) |
| Treatment for SAE | 0.02 (<0.01, 0.06) | 0.61 (0.19, 1.74) | 0.07 (<0.01, 0.25) | 0.07 (<0.01, 0.25) |
| **Total** | **100 (89, 117)** | **691 (645, 739)** | **87 (77, 101)** | **94 (83, 105)** |

Table S15: Cost per person for different gHAT treatments. PNLTHA markup is included. Because these are costs averaged over all patients and SAEs are rare, the average cost per patient for SAE is low.

#### S1.5 Cost-effectiveness analysis

Cost-effectiveness analyses assess the technical efficiency of several strategies in a head-to-head comparison. Such analyses denominate the resource inputs in terms of dollars and the health benefits in terms of DALYs. The amount of resource inputs that ought to be allocated for a given health benefit (the "willingness-to-pay" (WTP)) is not pre-designated in the analysis; rather, the optimal strategies are shown for a range of resource inputs (WTP).

While the purpose of such efforts is to inform the use of budgets, an analysis that optimises budgets across disease priorities is often intractable (see the WHO CHOICE program [19, 24]), but the use of certain agreed upon conventions helps make analyses comparable across disease programs. To that end, this analysis abides by the *Consolidated Health Economic Evaluation Reporting Standards (CHEERS)* guidelines. For more information, see the CHEERS checklist in section S5.

The basic metric of cost-effectiveness is the incremental cost-effectiveness ratio (ICER, see Box 1 Glossary), which is the cost of averting a DALY of disease compared to the next-best strategy [25, 26]. In recent years, and as reflected in the CHEERS checklist, there has been a tendency to integrate different forms of uncertainty in decision analyses. To address parameter uncertainty, the "net monetary benefits (NMB) framework" was developed to integrate the outputs of probabilistic modelling into a metric that integrates the benefits of ICERs [26, 27].

It should be noted, that the net benefits framework yields two metrics: the probability that a strategy is optimal (in terms of cost-effectiveness), and whether the strategy yields the highest expected net benefit. While the strategy with the highest probability of being optimal is often the strategy with the highest mean NMB, it is not always the case. This situations arises when key parameters are simulated from asymmetric distributions to characterize their uncertainty. This paradox evokes a discussion in the literature as to whether decision-makers are risk-averse or utility-maximizing, and it is beyond the scope of this discussion to engage in that debate [28], but it is mentioned here as some results may reflect this paradox (see Figures S5 and S6).

***Decision model used.*** The decision model used is in the form of a decision tree, although it contains both time-dependent disease transmission nodes and single-time point disease treatment nodes (Figure S2). The incidence of cases is modelled via an SEIRS model (briefly described in Section S1.2.1 but developed in [2]). The components could be conceptualised as shown in Figure 4. Although untreated infection lasts for 2–3 years, the majority of disease burden is borne by fatal cases and the majority of the costs arises from detection, so a detailed Markov model of disease progression with respect to time was not necessary.

***Costs included.*** It should be noted that cost-effectiveness analyses aim to quantify all economic costs in the perspective of the agent (the health system in our analysis) therefore, materials that are not directly purchased for the purpose of the named strategies but that are kept from other medical use are also quantified. A separate analysis by Sutherland *et al* [29] examined the financial as well as the economic impacts of gHAT strategies.

Because some of the strategies proposed in this paper have not been put into place, a model-based estimate of the costs has been employed in order to project the size of those costs, detailed in section S1.4.

***Time horizon and discounting.*** Since the purpose of the analysis is to assess the resource implications of EOT, which may require large investments in the short-term in return for long-term benefits, we have chosen the 20-year horizon as a default in the analysis. A ten-year horizon essentially discounts every year at 3% for ten years and at 100% thereafter, implying that long-term benefits are of no consequence to the decision-maker, but we do not believe this is the case in the context of gHAT elimination. We have provided select metrics in 10-year and 30-year horizons here in the supplement and in the companion website (see section S3.4). We have not chosen to provide time-horizons of longer than 30 years out of concern for our capacity to project robust costs and benefits this far into the future.

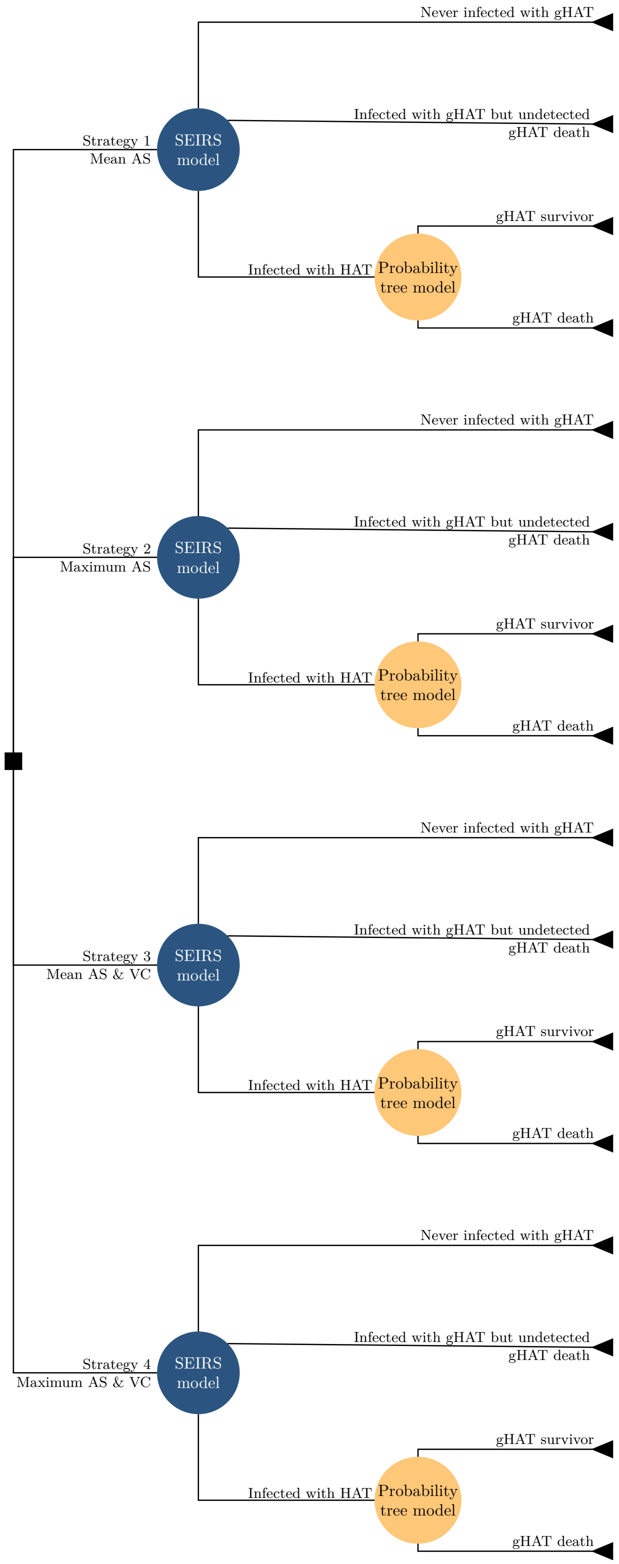


Figure S2: Decision tree model of the current analysis. The relationship between the dynamic (SIR) model and the probability tree model of disease treatment outcomes is shown here. The SIR model is described briefly in section S1.2.1, and the treatment model is shown in figure 1 and described in section S1.2.3.

#### S1.6 Incremental cost-effectiveness ratios, dominance, and weak dominance

We give here a step-by-step calculation of incremental cost-effectiveness ratios (ICERs) and how their calculation could lead to the conclusion that a strategy is under strong or weak dominance.

Below is the calculation for Kwamouth, assuming an investment horizon of ten years (2020–2030). Preliminary

| Strategy | Cost Difference | DALYs averted | Preliminary ICER | Final ICER |
|---|---|---|---|---|
| **2020-2030** | | | | |
| Mean AS | 0 | 0 | Min Cost | Min Cost |
| Max AS | 574,743 | 440 | 1,307 | Weakly Dominated |
| Mean AS & VC | 816,312 | 2,049 | 150 | 398 |
| Max AS & VC | 1,276,239 | 2,156 | 4,263 | 4,263 |

Table S16: Computing incremental cost-effectiveness ratios (ICERs) for Kwamouth.

ICERs display the difference in costs and effects between a given strategy and the next strategy of lesser value (in Kwamouth, it would be the intervention in the previous line, but Budjala the progression to more expensive interventions is more complicated). Due to diminishing returns, ICERs increase with more ambitious interventions. Some ICERs (shown in red) show that the strategy delivers a worse-value-for-money the next best intervention (in the following line). The strategies with red ICERs are 'weakly dominated' and should not be adopted. The final ICERs show the ICERs once weakly dominated strategies have been removed. The ICERs highlighted in yellow are ICERs that have changed with the removal of weakly dominated strategies.

| Strategy | Cost Difference | DALYs averted | Preliminary ICER | Final ICER |
|---|---|---|---|---|
| **2020-2040** | | | | |
| Mean AS | 0 | 0 | Min Cost | Min Cost |
| Max AS | 335,786 | 47 | 91,025 | Weakly Dominated |
| Mean AS & VC | 131,747 | 45 | 2,922 | 2,922 |
| Max AS & VC | 423,280 | 62 | 6,075 | 17,515 |

Table S17: Computing incremental cost-effectiveness ratios (ICERs) for Budjala

#### S1.7 Sensitivity Analysis

**Vector control.** While the default level of tsetse population reduction used in this paper (80%) appears realistic and possibly conservative [4, 13, 15, 30], the operations necessary to achieve a certain level of effectiveness for VC are unclear. Therefore, we performed three way sensitivity analysis of VC effectiveness, vector extent, and vector target density:

- VC is assumed to decrease the population of tsetse in the first year of deployment by 80% in the baseline analysis, but we examined outcomes and optimal choices assuming a decline of 60% and 90% in tsetse population. In Yasa Bonga, where a 90% decrease in the population of tsetse has been observed, we do not run this sensitivity analysis.
- The extent of the riverbank treated: for Mosango, Boma Bungu, and Budjala, where no VC activities are currently planned, we examined the cost-effectiveness of interventions if 210 km of riverbank had to be treated instead of the default assumption of 100 km.
- The density of targets deployed per kilometer of riverbank treated. We assumed that 20 targets per kilometer would be applied in Kwamouth (which is what is planned) and that 40 targets per kilometer would be necessary in Mosango, Boma Bungu, and Budjala. In sensitivity analyses, we tested the robustness of our decision analysis if the density of targets would have be doubled in Kwamouth or halved in Mosango, Boma Bungu, and Budjala.

We point out in the supplementary results (section S3.5) any policy-related changes to decision-making that were notable during sensitivity analyses, but complete calculations of costs, effects, and cost-effectiveness with each configuration of assumptions are available in the companion website for the paper: https://hatmepp.warwick.ac.uk/CEA/v1.

**Decision analysis conventions.** Two further elements of the decision analysis were re-examined in supplemental analyses. First, while an investment horizon of 2020–2040 was chosen for the principle analysis; shorter (2020–2030) and longer (2020–2050) investment horizons are reported in the supplementary results (section S3.4). In order to examine any differences in optimal choice. Second, while the convention in the cost-effectiveness literature is to discount both costs and health benefits by 3% per year, we have also reported the results when no discounting is taken into account. Results of these analyses are available in the companion website for the paper: https://hatmepp.warwick.ac.uk/CEA/v1.

### S2 Glossary of technical terms

**Box 1: Glossary**

**Epidemiology Terms**

**Intervention** Interventions are separate activities to address a health need (e.g. active screening (AS) or vector control (VC)).

**Strategy** A strategy is a combination of interventions, carried out with a specific coverage, and in parallel. In this paper, we simulate strategies with and without an improvement in AS and with and without VC (e.g. strategy 1 is passive surveillance (PS) and mean AS, and strategy 4 is PS, maximum AS and VC).

**Elimination of transmission (EOT)** Globally this is the 2030 goal for gHAT; here we also consider local EOT for health zones. The feasibility of EOT is expressed as a probability equal to the proportion of our simulations in which new infections is zero before a given year (usually 2030).

**Disability-adjusted life-year (DALY)** In order to present the burden of disease in one common metric across diseases, DALYs are calculated in cost-effectiveness analyses. This is the sum of the years lived with disability due to the disease and the years of life lost by fatal cases. See section S1.3.

**Health Economics Terms**

**Parameter uncertainty** Uncertainty in the level of transmission or in the costs of interventions and treatment due to unknown underlying parameters (see supplementary section S6 for an explanation of our parameterization of the health outcomes and cost model).

**Willingness-to-pay (WTP) or cost-effectiveness threshold** The amount of money that payers would pay to avert one DALY arising from the disease in the analysis (gHAT). No specific threshold is recommended, but a recent analysis shows that the WTP in DRC is between 5 - 230 USD per DALY averted [31–33].

**Incremental cost-effectiveness ratio** A ratio of marginal cost for a marginal benefit, calculated as follows:

$$\text{ICER} = \frac{\Delta\text{Costs}}{\Delta\text{DALYs}} = \frac{\text{Costs}_{\text{strategy}} - \text{Costs}_{\text{next best}}}{\text{Effects}_{\text{strategy}} - \text{Effects}_{\text{next best}}}$$

For an example on how interventions are ranked and ICERs are calculated, see Section S1.6.

**Cost-effective strategy** The strategy where the ICER is less than the WTP (or cost-effectiveness threshold). We say that the cost-effective strategy is “conditional" on the WTP.

**Dominated strategy** A strategy that costs more than the minimum cost intervention while reducing the burden by a smaller degree. This strategy ought not be implemented.

**Weakly dominated strategy (or strategies under extended dominance)** A strategy in which the ICER is higher that the next more expensive strategy. See section S1.6 for a discussion on this matter with respect to the strategies presented in this analysis.

**Net monetary benefit** The net benefits (NMB) framework is derived from ICERs, but also takes uncertainty into account.

$$\text{NMB}|\text{WTP} : \text{WTP} \times \Delta\text{DALYs} - \Delta\text{Costs}$$

The optimal strategy at a given WTP is the strategy with the highest mean NMB at that value of WTP.

**Optimal strategy** Analogous to the cost-effective strategy when no uncertainty is assumed, this is the strategy that is recommended by the NMB framework.

### S3 Supplementary results

S3.1 Supplemental outcome summaries . . . S30
S3.2 Supplemental cost summaries . . . S30
S3.2.1 Strategy costs per person, per case, and per area, 2020-2040 . . . S34
S3.3 Supplementary cost-effectiveness acceptability curves . . . S39
S3.4 Sensitivity analysis: time-horizon and time discounting . . . S41
S3.5 Sensitivity analysis: vector control . . . S45

#### S3.1 Supplemental outcome summaries

| | Cases detected (95% PI) | Deaths (95% PI) | DALYs (95% PI) | Total costs ($ millions) (95% PI) | Yearly cost ($) per capita (95% PI) |
|---|---|---|---|---|---|
| **Yasa Bonga** | | | | | |
| Mean AS & VC | 5 (0, 23) | 2 (0, 7) | 62 (1, 238) | 2.52 (1.14, 4.64) | 1.03 (0.47, 1.90) |
| Max AS & VC | 4 (0, 23) | 2 (0, 7) | 62 (1, 240) | 3.26 (1.32, 6.15) | 1.33 (0.54, 2.52) |
| **Mosango** | | | | | |
| Mean AS | 22 (1, 74) | 11 (1, 36) | 393 (32, 1,224) | 1.00 (0.46, 1.76) | 0.73 (0.34, 1.28) |
| Max AS | 21 (0, 91) | 8 (0, 25) | 270 (2, 888) | 1.44 (0.59, 2.72) | 1.04 (0.43, 1.98) |
| Mean AS & VC | 9 (0, 41) | 5 (0, 15) | 168 (2, 510) | 0.98 (0.47, 1.66) | 0.71 (0.34, 1.21) |
| Max AS & VC | 10 (0, 54) | 4 (0, 12) | 131 (1, 421) | 1.29 (0.58, 2.27) | 0.93 (0.42, 1.65) |
| **Kwamouth** | | | | | |
| Mean AS | 379 (129, 790) | 159 (36, 430) | 5,546 (1,333, 14,860) | 2.27 (1.60, 3.45) | 1.58 (1.11, 2.39) |
| Max AS | 379 (122, 809) | 136 (32, 363) | 4,773 (1,166, 12,527) | 2.95 (2.05, 4.59) | 2.05 (1.42, 3.18) |
| Mean AS & VC | 116 (41, 234) | 54 (17, 115) | 1,880 (627, 4,011) | 3.17 (2.06, 4.69) | 2.20 (1.43, 3.25) |
| Max AS & VC | 120 (38, 269) | 49 (16, 104) | 1,708 (561, 3,648) | 3.70 (2.33, 5.59) | 2.57 (1.62, 3.88) |
| **Boma Bungu** | | | | | |
| Mean AS | 1 (0, 10) | 0 (0, 4) | 17 (0, 149) | 0.27 (0.18, 0.40) | 0.29 (0.19, 0.42) |
| Max AS | 1 (0, 10) | 0 (0, 3) | 13 (0, 109) | 0.38 (0.22, 0.66) | 0.40 (0.23, 0.70) |
| Mean AS & VC | 1 (0, 7) | 0 (0, 3) | 13 (0, 107) | 0.41 (0.23, 0.69) | 0.43 (0.25, 0.73) |
| Max AS & VC | 1 (0, 8) | 0 (0, 3) | 11 (0, 97) | 0.51 (0.27, 0.92) | 0.54 (0.29, 0.98) |
| **Budjala** | | | | | |
| Mean AS | 4 (0, 21) | 5 (0, 16) | 159 (0, 564) | 0.29 (0.19, 0.42) | 0.20 (0.13, 0.29) |
| Max AS | 4 (0, 24) | 2 (0, 8) | 80 (0, 277) | 0.66 (0.24, 1.25) | 0.45 (0.16, 0.85) |
| Mean AS & VC | 2 (0, 12) | 2 (0, 8) | 83 (0, 274) | 0.43 (0.22, 0.74) | 0.29 (0.15, 0.50) |
| Max AS & VC | 3 (0, 19) | 2 (0, 6) | 56 (0, 208) | 0.75 (0.24, 1.39) | 0.51 (0.17, 0.94) |

Table S18: Summary of effects and costs 2020-2030. Two differences should be noted between these estimates and those used for decision analysis shown in table S25. First, these estimates are not discounted. Second due to asymmetric distributions, a naive difference in mean costs would not equal the mean differences in costs across simulations – the metric we used in decision analysis. Undetected cases are reflected in deaths, as very few deaths (<1 percent) originate from treated cases. Estimates shown are means and 95% prediction intervals (PI) of the cases, deaths, DALYs, and costs across iterations of the dynamic transmission model.

#### S3.2 Supplemental cost summaries

| | Cases detected (95% PI) | Deaths (95% PI) | DALYs (95% PI) | Total costs ($ millions) (95% PI) | Yearly cost ($) per capita (95% PI) |
|---|---|---|---|---|---|
| **Yasa Bonga** | | | | | |
| Mean AS & VC | 5 (0, 23) | 2 (0, 7) | 62 (1, 240) | 3.69 (2.10, 5.99) | 0.54 (0.31, 0.87) |
| Max AS & VC | 4 (0, 23) | 2 (0, 7) | 62 (1, 242) | 4.42 (2.31, 7.49) | 0.64 (0.34, 1.09) |
| **Mosango** | | | | | |
| Mean AS | 23 (1, 79) | 12 (1, 42) | 428 (32, 1,431) | 1.44 (0.76, 2.54) | 0.37 (0.20, 0.66) |
| Max AS | 22 (0, 92) | 8 (0, 28) | 282 (2, 987) | 1.86 (0.91, 3.42) | 0.48 (0.24, 0.88) |
| Mean AS & VC | 9 (0, 41) | 5 (0, 15) | 169 (2, 510) | 1.32 (0.77, 2.06) | 0.34 (0.20, 0.53) |
| Max AS & VC | 10 (0, 54) | 4 (0, 12) | 131 (1, 421) | 1.63 (0.90, 2.65) | 0.42 (0.23, 0.68) |
| **Kwamouth** | | | | | |
| Mean AS | 511 (144, 1,230) | 228 (41, 709) | 7,946 (1,514, 24,538) | 5.68 (3.58, 8.92) | 1.40 (0.88, 2.20) |
| Max AS | 490 (137, 1,158) | 188 (36, 566) | 6,575 (1,310, 19,499) | 7.20 (4.35, 11.78) | 1.77 (1.07, 2.90) |
| Mean AS & VC | 116 (41, 235) | 54 (18, 116) | 1,890 (628, 4,026) | 4.28 (2.88, 6.49) | 1.05 (0.71, 1.60) |
| Max AS & VC | 120 (38, 270) | 49 (16, 105) | 1,718 (562, 3,660) | 4.83 (3.16, 7.57) | 1.19 (0.78, 1.86) |
| **Boma Bungu** | | | | | |
| Mean AS | 1 (0, 10) | 0 (0, 4) | 17 (0, 149) | 0.70 (0.46, 1.02) | 0.26 (0.17, 0.38) |
| Max AS | 1 (0, 10) | 0 (0, 3) | 13 (0, 109) | 0.81 (0.52, 1.20) | 0.30 (0.20, 0.45) |
| Mean AS & VC | 1 (0, 7) | 0 (0, 3) | 13 (0, 107) | 0.84 (0.54, 1.23) | 0.32 (0.20, 0.46) |
| Max AS & VC | 1 (0, 8) | 0 (0, 3) | 11 (0, 97) | 0.94 (0.58, 1.42) | 0.35 (0.22, 0.53) |
| **Budjala** | | | | | |
| Mean AS | 4 (0, 22) | 5 (0, 18) | 163 (0, 601) | 0.81 (0.53, 1.18) | 0.20 (0.13, 0.28) |
| Max AS | 4 (0, 24) | 2 (0, 8) | 80 (0, 277) | 1.17 (0.66, 1.86) | 0.28 (0.16, 0.45) |
| Mean AS & VC | 2 (0, 12) | 2 (0, 8) | 83 (0, 274) | 0.95 (0.59, 1.41) | 0.23 (0.14, 0.34) |
| Max AS & VC | 3 (0, 19) | 2 (0, 6) | 56 (0, 208) | 1.26 (0.67, 1.99) | 0.31 (0.16, 0.48) |

Table S19: Summary of effects and costs 2020-2050. Two differences should be noted between these estimates and those used for decision analysis shown in table S26. First, these estimates are not discounted. Second due to asymmetric distributions, a naive difference in mean costs would not equal the mean differences in costs across simulations – the metric we used in decision analysis. Undetected cases are reflected in deaths, as very few deaths (<1 percent) originate from treated cases. Estimates shown are means and 95% prediction intervals (PI) of the cases, deaths, DALYs, and costs across iterations of the dynamic transmission model.

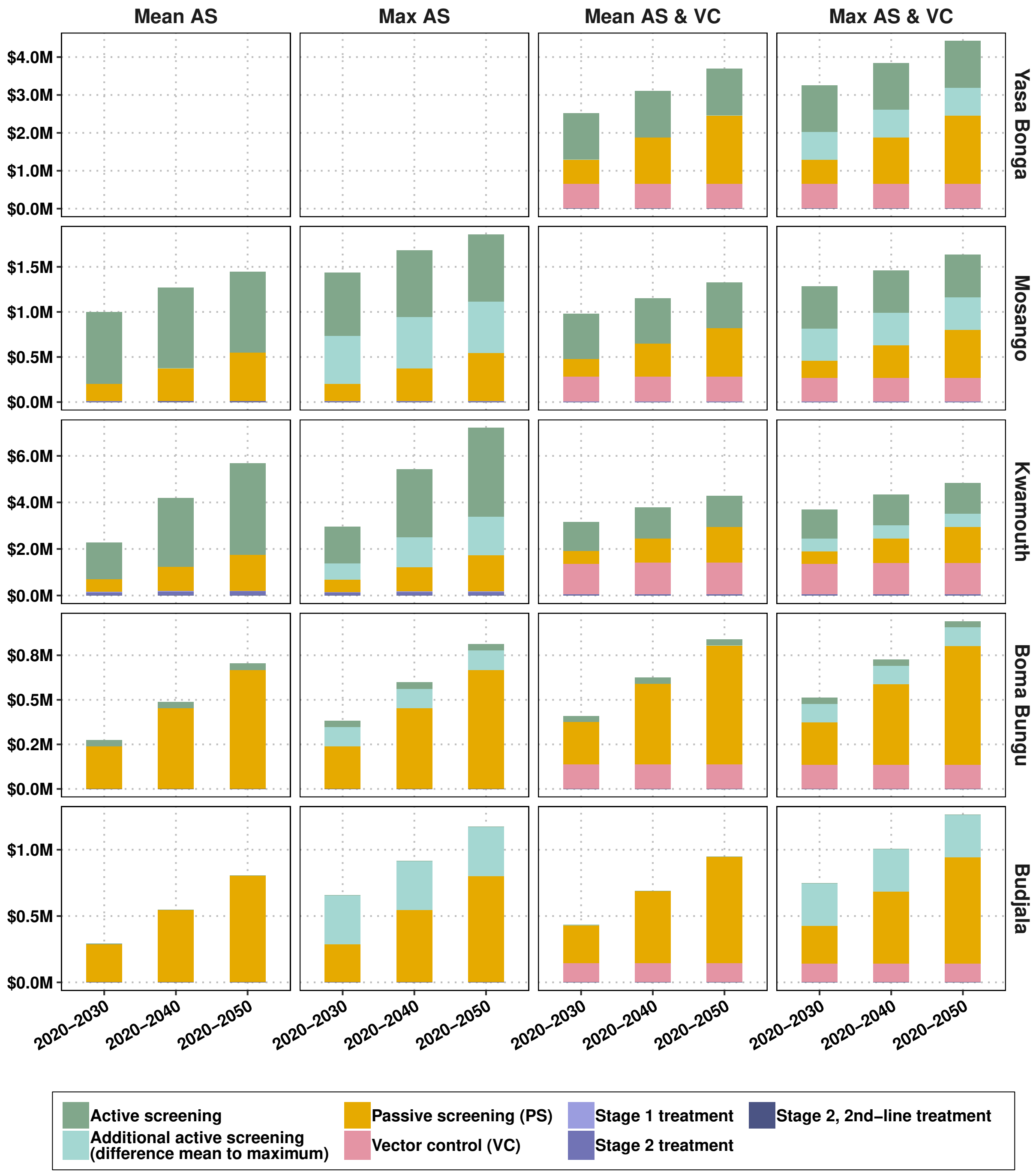


Figure S3: Components of cumulative costs across three time horizons, by strategy and location. Costs are not discounted.

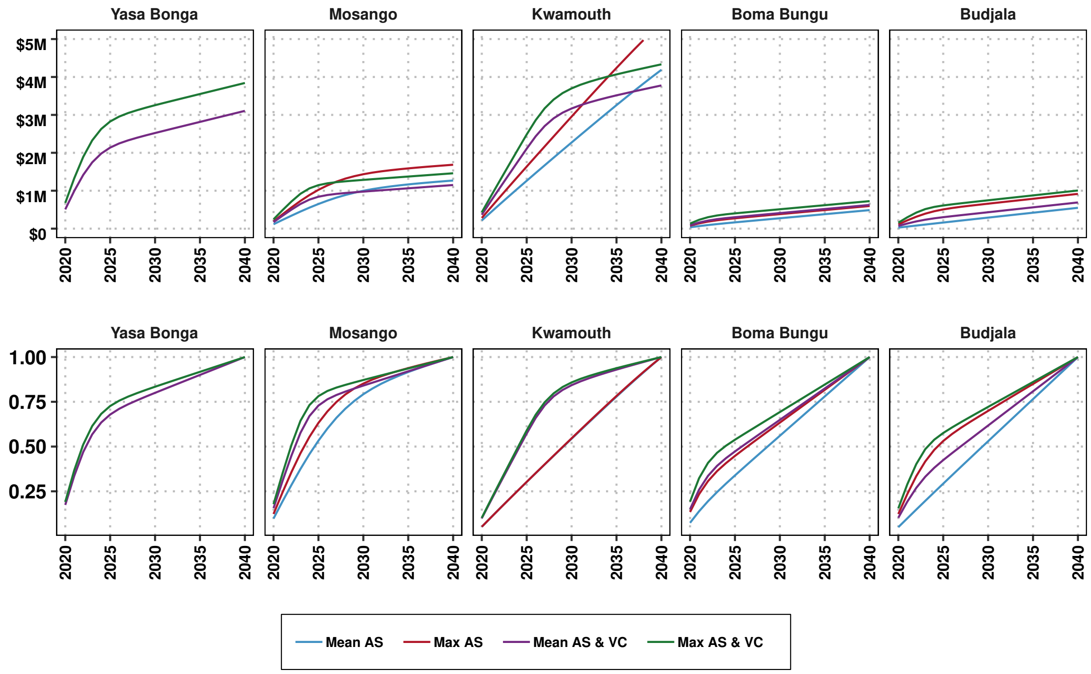


Figure S4: Cumulative costs for each strategy through time, by health zone (top row) and the percent of the total cost spent by each year (bottom). Costs are not discounted.

##### S3.2.1 Strategy costs per person, per case, and per area, 2020-2040

| | Mean AS & VC | Max AS & VC |
|---|---|---|
| **Active screening** | | |
| Total cases found | 0 (0, 0) | 0 (0, 0) |
| People screened | 538,327 (126,007, 1,008,056) | 860,932 (201,251, 1,610,008) |
| Stage 1 cases found | 0 (0, 0) | 0 (0, 0) |
| Stage 2 cases found | 0 (0, 0) | 0 (0, 0) |
| Total costs | 1,233,844 (325,683, 2,744,540) | 1,971,127 (499,872, 4,364,954) |
| Cost per person screened | 2.29 (1.44, 3.87) | 2.29 (1.44, 3.87) |
| Cost per capita | 5.56 (1.47, 12.37) | 8.88 (2.25, 19.67) |
| Cost per case detected | 1,232,131 (319,224, 2,741,385) | 1,970,133 (493,072, 4,364,954) |
| Pr. zero detections via AS | >0.99 | >0.99 |
| **Passive screening** | | |
| Total cases found | 5 (0, 23) | 4 (0, 23) |
| People screened | 123,751 (67,406, 195,510) | 123,751 (67,406, 195,510) |
| Cases stage 1 found | 0 (0, 1) | 0 (0, 1) |
| Cases stage 2 found | 4 (0, 23) | 4 (0, 22) |
| Total costs | 1,221,210 (749,019, 1,894,498) | 1,221,210 (749,019, 1,894,498) |
| Cost per person screened | 9.91 (7.08, 14.06) | 9.91 (7.08, 14.06) |
| Cost per capita | 5.50 (3.38, 8.54) | 5.50 (3.38, 8.54) |
| Cost per case detected | 713,580 (48,480, 1,747,968) | 720,392 (48,120, 1,765,962) |
| Pr. zero detections via PS | 0.34 | 0.34 |
| **Vector control** | | |
| Total costs | 648,662 (151,591, 1,503,502) | 646,984 (147,933, 1,479,354) |
| Cost per area | 249 (58, 577) | 248 (57, 568) |
| Cost per capita | 2.92 (0.68, 6.78) | 2.92 (0.67, 6.67) |

Table S20: Detailed summary of active screening, passive screening, and vector control in Yasa Bonga for the period of 2020-2040. Costs are not discounted.

| | Mean AS | Max AS | Mean AS & VC | Max AS & VC |
|---|---|---|---|---|
| **Active screening** | | | | |
| Total cases found | 10 (0, 53) | 13 (0, 76) | 4 (0, 32) | 6 (0, 48) |
| People screened | 390,825 (127,128, 720,392) | 572,093 (224,280, 1,046,640) | 219,694 (84,752, 339,008) | 362,878 (149,520, 598,080) |
| Stage 1 cases found | 7 (0, 35) | 9 (0, 52) | 3 (0, 20) | 4 (0, 32) |
| Stage 2 cases found | 4 (0, 20) | 4 (0, 26) | 2 (0, 13) | 2 (0, 19) |
| Total costs | 895,966 (279,880, 1,919,511) | 1,313,464 (400,973, 2,871,685) | 504,360 (185,375, 1,018,169) | 831,432 (293,111, 1,668,199) |
| Cost per person screened | 2.29 (1.44, 3.87) | 2.29 (1.44, 3.87) | 2.29 (1.44, 3.87) | 2.29 (1.44, 3.87) |
| Cost per capita | 7.16 (2.24, 15.35) | 10.50 (3.21, 22.96) | 4.03 (1.48, 8.14) | 6.65 (2.34, 13.34) |
| Cost per case detected | 350,428 (18,207, 1,233,222) | 539,729 (17,791, 1,867,479) | 333,907 (15,141, 885,973) | 524,994 (17,058, 1,418,912) |
| Pr. zero detections via AS | 0.30 | 0.33 | 0.52 | 0.52 |
| **Passive screening** | | | | |
| Total cases found | 13 (0, 42) | 9 (0, 31) | 5 (0, 20) | 4 (0, 17) |
| People screened | 17,437 (9,498, 27,548) | 17,437 (9,498, 27,548) | 17,437 (9,498, 27,548) | 17,437 (9,498, 27,548) |
| Cases stage 1 found | 2 (0, 10) | 2 (0, 8) | 1 (0, 4) | 1 (0, 3) |
| Cases stage 2 found | 10 (0, 34) | 7 (0, 25) | 4 (0, 18) | 3 (0, 15) |
| Total costs | 363,532 (227,948, 542,553) | 363,532 (227,948, 542,553) | 363,532 (227,948, 542,553) | 363,532 (227,948, 542,553) |
| Cost per person screened | 20.77 (12.79, 37.23) | 20.77 (12.79, 37.23) | 20.77 (12.79, 37.23) | 20.77 (12.79, 37.23) |
| Cost per capita | 2.91 (1.82, 4.34) | 2.91 (1.82, 4.34) | 2.91 (1.82, 4.34) | 2.91 (1.82, 4.34) |
| Cost per case detected | 77,306 (7,879, 406,682) | 116,479 (10,355, 456,496) | 171,730 (16,009, 486,859) | 202,667 (19,613, 504,582) |
| Pr. zero detections via PS | 0.04 | 0.09 | 0.18 | 0.26 |
| **Vector control** | | | | |
| Total costs | 0 (0, 0) | 0 (0, 0) | 278,549 (79,794, 559,701) | 261,416 (76,490, 526,134) |
| Cost per area | NA | NA | 104 (30, 209) | 98 (29, 197) |
| Cost per capita | NA | NA | 2.23 (0.64, 4.47) | 2.09 (0.61, 4.21) |

Table S21: Detailed summary of active screening, passive screening, and vector control in Mosango for the period of 2020-2040. Costs are not discounted.

| | Mean AS | Max AS | Mean AS & VC | Max AS & VC |
|---|---|---|---|---|
| **Active screening** | | | | |
| Total cases found | 189 (30, 479) | 222 (33, 580) | 46 (2, 143) | 56 (2, 188) |
| People screened | 1,300,535 (1,072,377, 1,324,701) | 1,852,338 (1,450,880, 1,904,280) | 584,830 (378,486, 946,215) | 834,809 (544,080, 1,360,200) |
| Stage 1 cases found | 127 (17, 354) | 155 (20, 440) | 28 (1, 93) | 36 (1, 126) |
| Stage 2 cases found | 62 (11, 150) | 66 (11, 162) | 18 (0, 61) | 20 (0, 70) |
| Total costs | 2,961,960 (1,824,236, 5,040,768) | 4,217,655 (2,544,127, 7,191,397) | 1,331,356 (681,211, 2,631,073) | 1,899,505 (961,791, 3,752,968) |
| Cost per person screened | 2.28 (1.43, 3.84) | 2.28 (1.43, 3.84) | 2.28 (1.43, 3.84) | 2.28 (1.43, 3.84) |
| Cost per capita | 22.61 (13.92, 38.47) | 32.19 (19.42, 54.89) | 10.16 (5.20, 20.08) | 14.50 (7.34, 28.64) |
| Cost per case detected | 25,802 (5,412, 93,864) | 32,805 (6,515, 120,955) | 88,491 (7,481, 611,920) | 131,763 (8,189, 1,083,753) |
| Pr. zero detections via AS | <0.01 | <0.01 | <0.01 | 0.01 |
| **Passive screening** | | | | |
| Total cases found | 287 (96, 635) | 242 (80, 530) | 70 (27, 131) | 64 (23, 123) |
| People screened | 91,330 (49,747, 144,288) | 91,330 (49,747, 144,288) | 91,330 (49,747, 144,288) | 91,330 (49,747, 144,288) |
| Cases stage 1 found | 87 (26, 195) | 74 (23, 166) | 14 (4, 30) | 13 (3, 27) |
| Cases stage 2 found | 200 (66, 457) | 168 (55, 373) | 56 (21, 110) | 51 (18, 105) |
| Total costs | 1,041,399 (659,928, 1,566,012) | 1,041,399 (659,928, 1,566,012) | 1,041,399 (659,928, 1,566,012) | 1,041,399 (659,928, 1,566,012) |
| Cost per person screened | 11.46 (8.04, 16.92) | 11.46 (8.04, 16.92) | 11.46 (8.04, 16.92) | 11.46 (8.04, 16.92) |
| Cost per capita | 7.95 (5.04, 11.95) | 7.95 (5.04, 11.95) | 7.95 (5.04, 11.95) | 7.95 (5.04, 11.95) |
| Cost per case detected | 4,613 (1,428, 11,616) | 5,435 (1,745, 13,853) | 17,588 (6,714, 40,881) | 19,457 (7,236, 46,573) |
| Pr. zero detections via PS | <0.01 | <0.01 | <0.01 | <0.01 |
| **Vector control** | | | | |
| Total costs | 0 (0, 0) | 0 (0, 0) | 1,353,920 (660,706, 2,545,956) | 1,343,034 (648,485, 2,538,634) |
| Cost per area | NA | NA | 93 (45, 175) | 92 (44, 174) |
| Cost per capita | NA | NA | 10.33 (5.04, 19.43) | 10.25 (4.95, 19.38) |

Table S22: Detailed summary of active screening, passive screening, and vector control in Kwamouth for the period of 2020-2040. Costs are not discounted.

| | Mean AS | Max AS | Mean AS & VC | Max AS & VC |
|---|---|---|---|---|
| **Active screening** | | | | |
| Total cases found | 0 (0, 1) | 0 (0, 4) | 0 (0, 1) | 0 (0, 3) |
| People screened | 16,160 (6,142, 42,994) | 63,263 (24,744, 148,464) | 15,441 (6,142, 36,852) | 60,727 (24,744, 148,464) |
| Stage 1 cases found | 0 (0, 1) | 0 (0, 2) | 0 (0, 0) | 0 (0, 1) |
| Stage 2 cases found | 0 (0, 1) | 0 (0, 2) | 0 (0, 0) | 0 (0, 1) |
| Total costs | 37,176 (9,415, 106,860) | 145,680 (37,888, 406,611) | 35,536 (9,415, 94,364) | 139,533 (37,924, 362,492) |
| Cost per person screened | 2.30 (1.44, 3.88) | 2.30 (1.44, 3.88) | 2.30 (1.44, 3.88) | 2.30 (1.44, 3.88) |
| Cost per capita | 0.43 (0.11, 1.24) | 1.69 (0.44, 4.73) | 0.41 (0.11, 1.10) | 1.62 (0.44, 4.22) |
| Cost per case detected | 36,082 (9,395, 101,975) | 135,510 (34,445, 371,329) | 34,800 (9,336, 91,462) | 132,012 (34,858, 345,606) |
| Pr. zero detections via AS | 0.96 | 0.94 | 0.97 | 0.94 |
| **Passive screening** | | | | |
| Total cases found | 1 (0, 9) | 1 (0, 7) | 1 (0, 7) | 1 (0, 6) |
| People screened | 23,968 (13,055, 37,866) | 23,968 (13,055, 37,866) | 23,968 (13,055, 37,866) | 23,968 (13,055, 37,866) |
| Cases stage 1 found | 0 (0, 4) | 0 (0, 3) | 0 (0, 3) | 0 (0, 2) |
| Cases stage 2 found | 1 (0, 5) | 0 (0, 5) | 0 (0, 5) | 0 (0, 4) |
| Total costs | 451,039 (292,720, 657,315) | 451,039 (292,720, 657,315) | 451,039 (292,720, 657,315) | 451,039 (292,720, 657,315) |
| Cost per person screened | 18.85 (12.09, 32.24) | 18.85 (12.09, 32.24) | 18.85 (12.09, 32.24) | 18.85 (12.09, 32.24) |
| Cost per capita | 5.25 (3.41, 7.65) | 5.25 (3.41, 7.65) | 5.25 (3.41, 7.65) | 5.25 (3.41, 7.65) |
| Cost per case detected | 397,682 (48,218, 644,710) | 405,111 (59,589, 649,568) | 404,892 (63,109, 649,492) | 413,726 (74,397, 652,577) |
| Pr. zero detections via PS | 0.75 | 0.78 | 0.77 | 0.80 |
| **Vector control** | | | | |
| Total costs | 0 (0, 0) | 0 (0, 0) | 137,698 (34,163, 358,971) | 134,888 (34,129, 347,038) |
| Cost per area | NA | NA | 48 (12, 125) | 47 (12, 121) |
| Cost per capita | NA | NA | 1.60 (0.40, 4.18) | 1.57 (0.40, 4.04) |

Table S23: Detailed summary of active screening, passive screening, and vector control in Boma Bungu for the period of 2020-2040. Costs are not discounted.

| | Mean AS | Max AS | Mean AS & VC | Max AS & VC |
|---|---|---|---|---|
| **Active screening** | | | | |
| Total cases found | 0 (0, 1) | 2 (0, 18) | 0 (0, 0) | 1 (0, 15) |
| People screened | 2,331 (0, 5,973) | 165,660 (0, 339,696) | 1,677 (0, 3,801) | 142,352 (0, 291,168) |
| Stage 1 cases found | 0 (0, 0) | 1 (0, 12) | 0 (0, 0) | 1 (0, 9) |
| Stage 2 cases found | 0 (0, 0) | 1 (0, 6) | 0 (0, 0) | 0 (0, 6) |
| Total costs | 5,343 (0, 15,172) | 373,576 (0, 943,892) | 3,851 (0, 9,585) | 321,393 (0, 779,730) |
| Cost per person screened | 2.30 (1.44, 3.88) | 2.26 (1.42, 3.81) | 2.30 (1.45, 3.88) | 2.26 (1.42, 3.81) |
| Cost per capita | 0.04 (<0.01, 0.11) | 2.80 (<0.01, 7.07) | 0.03 (<0.01, 0.07) | 2.41 (<0.01, 5.84) |
| Cost per case detected | 5,326 (0, 15,152) | 297,971 (0, 829,900) | 3,848 (0, 9,571) | 270,249 (0, 708,646) |
| Pr. zero detections via AS | 0.97 | 0.79 | 0.98 | 0.82 |
| **Passive screening** | | | | |
| Total cases found | 4 (0, 22) | 2 (0, 12) | 2 (0, 12) | 1 (0, 10) |
| People screened | 37,202 (20,264, 58,774) | 37,202 (20,264, 58,774) | 37,202 (20,264, 58,774) | 37,202 (20,264, 58,774) |
| Cases stage 1 found | 2 (0, 9) | 1 (0, 5) | 1 (0, 5) | 0 (0, 4) |
| Cases stage 2 found | 3 (0, 15) | 1 (0, 8) | 1 (0, 8) | 1 (0, 7) |
| Total costs | 541,863 (351,941, 794,279) | 541,863 (351,941, 794,279) | 541,863 (351,941, 794,279) | 541,863 (351,941, 794,279) |
| Cost per person screened | 14.63 (9.87, 23.37) | 14.63 (9.87, 23.37) | 14.63 (9.87, 23.37) | 14.63 (9.87, 23.37) |
| Cost per capita | 4.06 (2.64, 5.95) | 4.06 (2.64, 5.95) | 4.06 (2.64, 5.95) | 4.06 (2.64, 5.95) |
| Cost per case detected | 314,247 (23,299, 743,891) | 399,535 (40,726, 765,310) | 404,435 (41,593, 765,381) | 440,375 (50,750, 775,746) |
| Pr. zero detections via PS | 0.33 | 0.48 | 0.49 | 0.59 |
| **Vector control** | | | | |
| Total costs | 0 (0, 0) | 0 (0, 0) | 143,999 (0, 414,413) | 141,440 (0, 387,365) |
| Cost per area | NA | NA | 33 (0, 94) | 32 (0, 88) |
| Cost per capita | NA | NA | 1.08 (<0.01, 3.11) | 1.06 (<0.01, 2.90) |

Table S24: Detailed summary of active screening, passive screening, and vector control in Budjala for the period of 2020-2040. Costs are not discounted.

507 ## S3.3 Supplementary cost-effectiveness acceptability curves

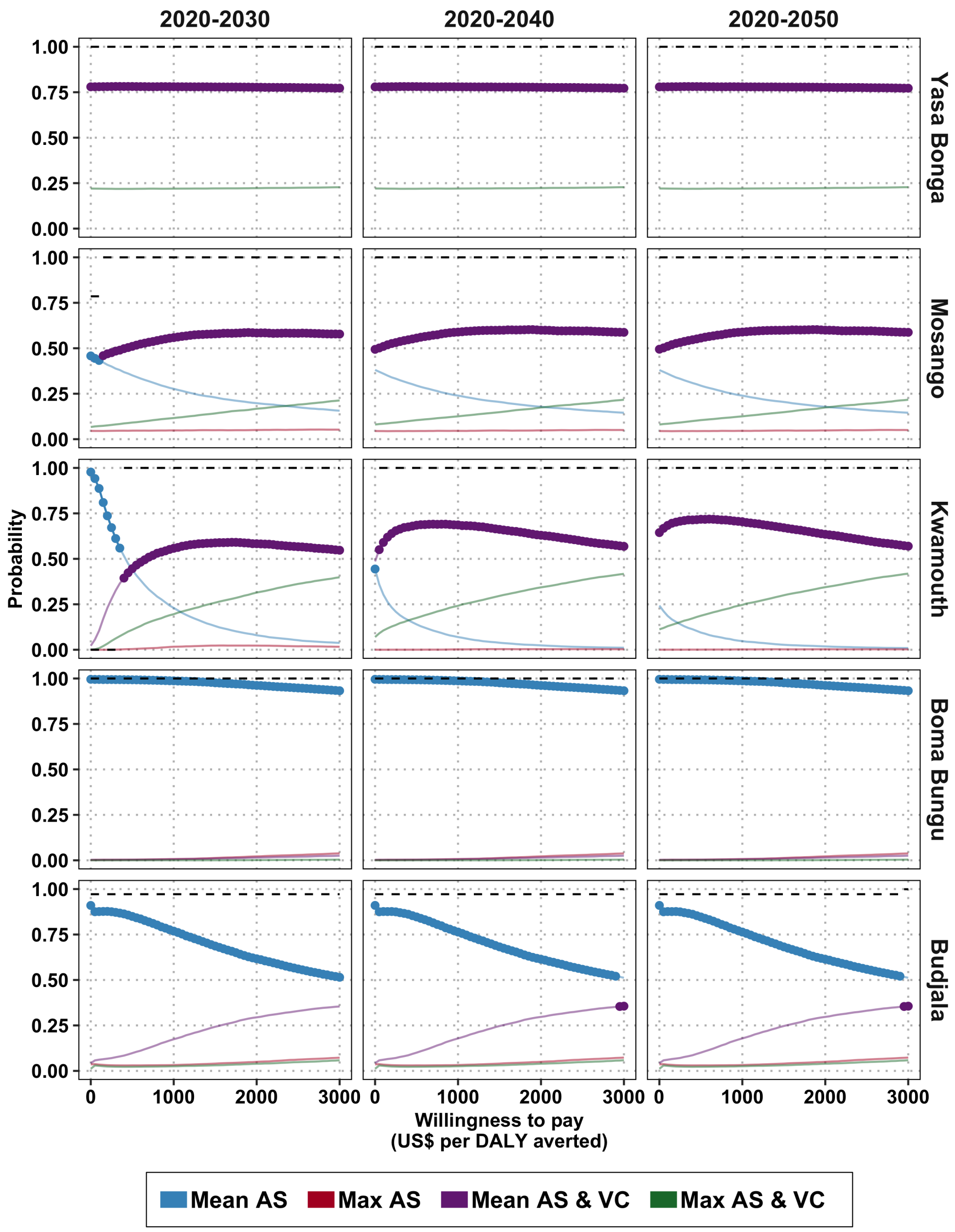


Figure S5: Cost-effectiveness acceptability curves across different horizons assuming both costs and health effects are discounted. The probability that each intervention is optimal (cost-effective) is expressed by the solid lines in colors. The preferred intervention (in terms of mean net monetary benefit) is expressed by the dotted lines. The black dashed line shows probability that the preferred intervention reaches elimination of transmission (EOT) by 2030. For example, in Kwamouth, if one considers a 10-year horizon for investments, the strategy 'Mean AS' is cost effective at willingness-to-pay values below $500 per DALY averted, as shown by the dark blue dotted line, but that strategy has almost zero probability of reaching EOT by 2030, as shown by the black dashed line. If a decision-maker is willing to pay more than $500 per DALY averted, then the cost-effective strategy is 'Mean AS & VC', as displayed by the dark purple dotted line, and that strategy has an almost 100% probability of reaching EoT by 2030.

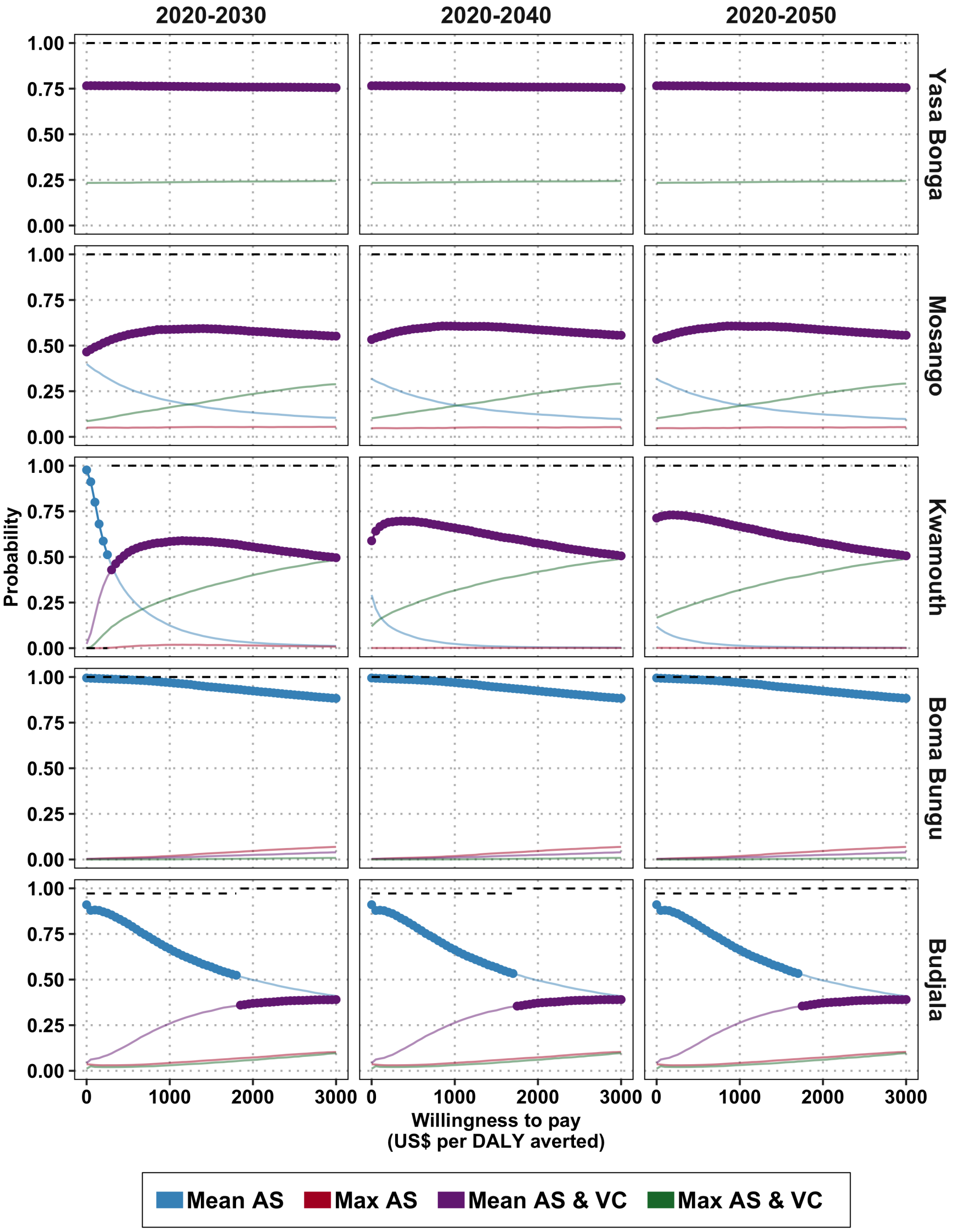


Figure S6: Cost-effectiveness acceptability curves with no cost or health benefit discounts. The probability of cost-effectiveness is expressed by the solid lines. The preferred intervention (in terms of mean net monetary benefit) is expressed by the dotted dark line. The black dashed line shows probability that the preferred intervention reaches elimination of transmission (EoT) by 2030. For example, in Kwamouth, if one considers a 10-year horizon for investments, the strategy 'Mean AS' is cost effective at willingness-to-pay values below $500 per DALY averted, as shown by the dark blue dotted line, but that strategy has almost zero probability of reaching EoT by 2030, as shown by the black dashed line. If a decision-maker is willing to pay more than $500 per DALY averted, then the cost-effective strategy is 'Mean AS & VC', as displayed by the dark purple dotted line, and that strategy has an almost 100% probability of reaching EoT by 2030.

#### S3.4 Sensitivity analysis: time-horizon and time discounting

| | Cost-effectiveness analysis without uncertainty | | | Net benefit (uncertainty) analysis: Prob. that a strategy is optimal, (conditional on willingness-to-pay) | | | | |
|---|---|---|---|---|---|---|---|---|
| | Cost difference vs comparator | DALYs averted vs comparator | ICER | $0 USD per DALY averted | $250 per DALY averted | $500 per DALY averted | $1,000 per DALY averted | Prob. EOT by 2030 |
| **Yasa Bonga** | | | | | | | | |
| Mean AS & VC | 0 | 0 | Min Cost | 0.78 | 0.78 | 0.78 | 0.78 | >0.99 |
| Max AS & VC | 670,123 | 0 | 2,156,689 | 0.22 | 0.22 | 0.22 | 0.22 | >0.99 |
| **Mosango** | | | | | | | | |
| Mean AS | 0 | 0 | Min Cost | 0.46 | 0.4 | 0.35 | 0.28 | 0.79 |
| Max AS | 390,631 | 71 | Dominated | 0.05 | 0.05 | 0.05 | 0.05 | 0.92 |
| Mean AS & VC | 15,695 | 127 | 123 | 0.43 | 0.48 | 0.51 | 0.56 | >0.99 |
| Max AS & VC | 301,004 | 150 | 12,196 | 0.07 | 0.08 | 0.09 | 0.12 | >0.99 |
| **Kwamouth** | | | | | | | | |
| Mean AS | 0 | 0 | Min Cost | 0.98 | 0.67 | 0.44 | 0.23 | <0.01 |
| Max AS | 574,743 | 440 | Weakly Dominated | 0 | 0 | 0 | 0.02 | <0.01 |
| Mean AS & VC | 816,312 | 2,049 | 398 | 0.02 | 0.28 | 0.45 | 0.56 | >0.99 |
| Max AS & VC | 1,276,239 | 2,156 | 4,263 | 0 | 0.05 | 0.11 | 0.2 | >0.99 |
| **Boma Bungu** | | | | | | | | |
| Mean AS | 0 | 0 | Min Cost | 1 | 0.99 | 0.99 | 0.99 | >0.99 |
| Max AS | 101,573 | 3 | 40,380 | 0 | 0 | 0 | 0.01 | >0.99 |
| Mean AS & VC | 127,904 | 2 | Dominated | 0 | 0 | 0 | 0.01 | >0.99 |
| Max AS & VC | 223,244 | 4 | 93,437 | 0 | 0 | 0 | 0 | >0.99 |
| **Budjala** | | | | | | | | |
| Mean AS | 0 | 0 | Min Cost | 0.91 | 0.88 | 0.85 | 0.77 | 0.97 |
| Max AS | 335,514 | 45 | Weakly Dominated | 0.04 | 0.03 | 0.03 | 0.03 | >0.99 |
| Mean AS & VC | 131,875 | 43 | 3,049 | 0.04 | 0.07 | 0.1 | 0.17 | >0.99 |
| Max AS & VC | 423,315 | 60 | 17,581 | 0.01 | 0.02 | 0.02 | 0.03 | >0.99 |

Table S25: Summary of cost-effectiveness, assuming a time horizon of 2020-2030. Cost differences and DALYs averted are relative to the comparator–first strategy listed for each location. Mean DALYs averted and mean cost differences are shown; these estimates are discounted at 3 percent per year in accordance with guidelines. The uncertainty analysis (columns 5-8) shows the probability that a strategy is cost-effective. Strategies highlighted in pink are optimal strategies: the strategies for which the mean net monetary benefit (NMB) is highest. ICER: incremental cost-effectiveness ratio, DALY: disability adjusted life-years. For an extended discussion of these terms, see supplement section S1.5.

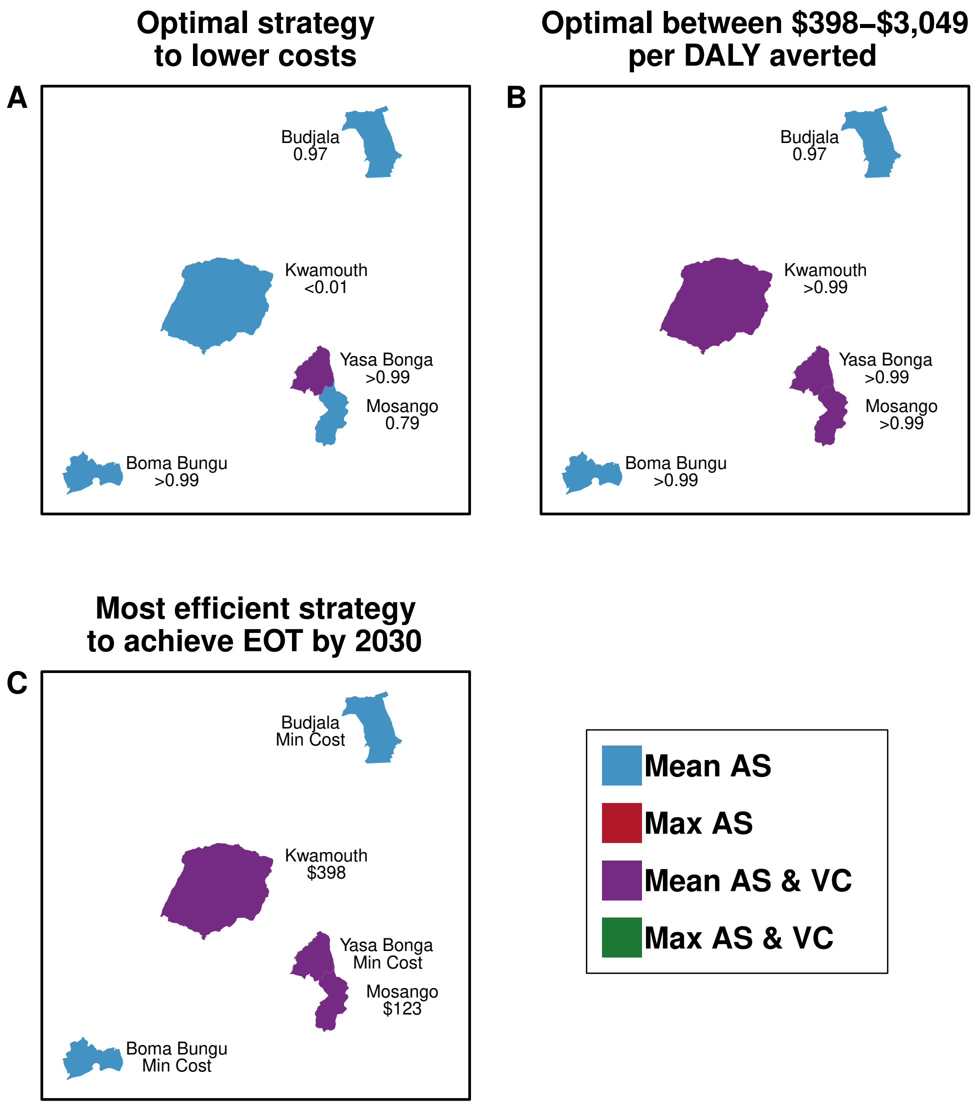


Figure S7: Maps of preferred strategies according to economic or budgetary goals for 2020–2030. Maps A & B show the optimal strategies depending on WTP. The text indicates the probability that the optimal strategy will lead to EOT by 2030. Map C shows the optimal strategy that has at >90% probability of EOT by 2030 and shows the ICER of the indicated strategy (Mean AS for all locations except Yasa Bonga, where it is Mean AS & VC). Maps are not drawn to scale.

| | Cost-effectiveness analysis without uncertainty | | | Net benefit (uncertainty) analysis: Prob. that a strategy is optimal, (conditional on willingness-to-pay) | | | | |
|---|---|---|---|---|---|---|---|---|
| | Cost difference vs comparator | DALYs averted vs comparator | ICER | $0 USD per DALY averted | $250 per DALY averted | $500 per DALY averted | $1,000 per DALY averted | Prob. EOT by 2030 |
| **Yasa Bonga** | | | | | | | | |
| Mean AS & VC | 0 | 0 | Min Cost | 0.78 | 0.78 | 0.78 | 0.78 | >0.99 |
| Max AS & VC | 671,462 | 0 | 2,209,891 | 0.22 | 0.22 | 0.22 | 0.22 | >0.99 |
| **Mosango** | | | | | | | | |
| Mean AS | 0 | 0 | Dominated | 0.38 | 0.33 | 0.29 | 0.24 | 0.79 |
| Max AS | 376,544 | 80 | Dominated | 0.04 | 0.04 | 0.04 | 0.05 | 0.92 |
| Mean AS & VC | -49,378 | 142 | Min Cost | 0.49 | 0.53 | 0.56 | 0.59 | >0.99 |
| Max AS & VC | 236,263 | 166 | 12,216 | 0.08 | 0.09 | 0.1 | 0.13 | >0.99 |
| **Kwamouth** | | | | | | | | |
| Mean AS | 0 | 0 | Dominated | 0.24 | 0.14 | 0.09 | 0.05 | <0.01 |
| Max AS | 1,052,454 | 667 | Dominated | 0 | 0 | 0 | 0 | <0.01 |
| Mean AS & VC | -451,240 | 2,976 | Min Cost | 0.64 | 0.71 | 0.72 | 0.7 | >0.99 |
| Max AS & VC | 26,747 | 3,084 | 4,426 | 0.11 | 0.15 | 0.19 | 0.25 | >0.99 |
| **Boma Bungu** | | | | | | | | |
| Mean AS | 0 | 0 | Min Cost | 1 | 0.99 | 0.99 | 0.99 | >0.99 |
| Max AS | 101,606 | 3 | 40,288 | 0 | 0 | 0 | 0.01 | >0.99 |
| Mean AS & VC | 127,894 | 2 | Dominated | 0 | 0 | 0 | 0.01 | >0.99 |
| Max AS & VC | 223,232 | 4 | 93,059 | 0 | 0 | 0 | 0 | >0.99 |
| **Budjala** | | | | | | | | |
| Mean AS | 0 | 0 | Min Cost | 0.91 | 0.87 | 0.85 | 0.76 | 0.97 |
| Max AS | 335,786 | 47 | Weakly Dominated | 0.04 | 0.03 | 0.03 | 0.03 | >0.99 |
| Mean AS & VC | 131,746 | 45 | 2,921 | 0.04 | 0.07 | 0.1 | 0.18 | >0.99 |
| Max AS & VC | 423,279 | 62 | 17,515 | 0.01 | 0.02 | 0.02 | 0.03 | >0.99 |

Table S26: Summary of cost-effectiveness, assuming a time horizon of 2020-2050. Cost differences and DALYs averted are relative to the comparator–first strategy listed for each location. Mean DALYs averted and mean cost differences are shown; these estimates are discounted at 3 percent per year in accordance with guidelines. The uncertainty analysis (columns 5-8) shows the probability that a strategy is cost-effective. Strategies highlighted in pink are optimal strategies: the strategies for which the mean net monetary benefit (NMB) is highest. ICER: incremental cost-effectiveness ratio, DALY: disability adjusted life-years. For an extended discussion of these terms, see supplement section S1.5.

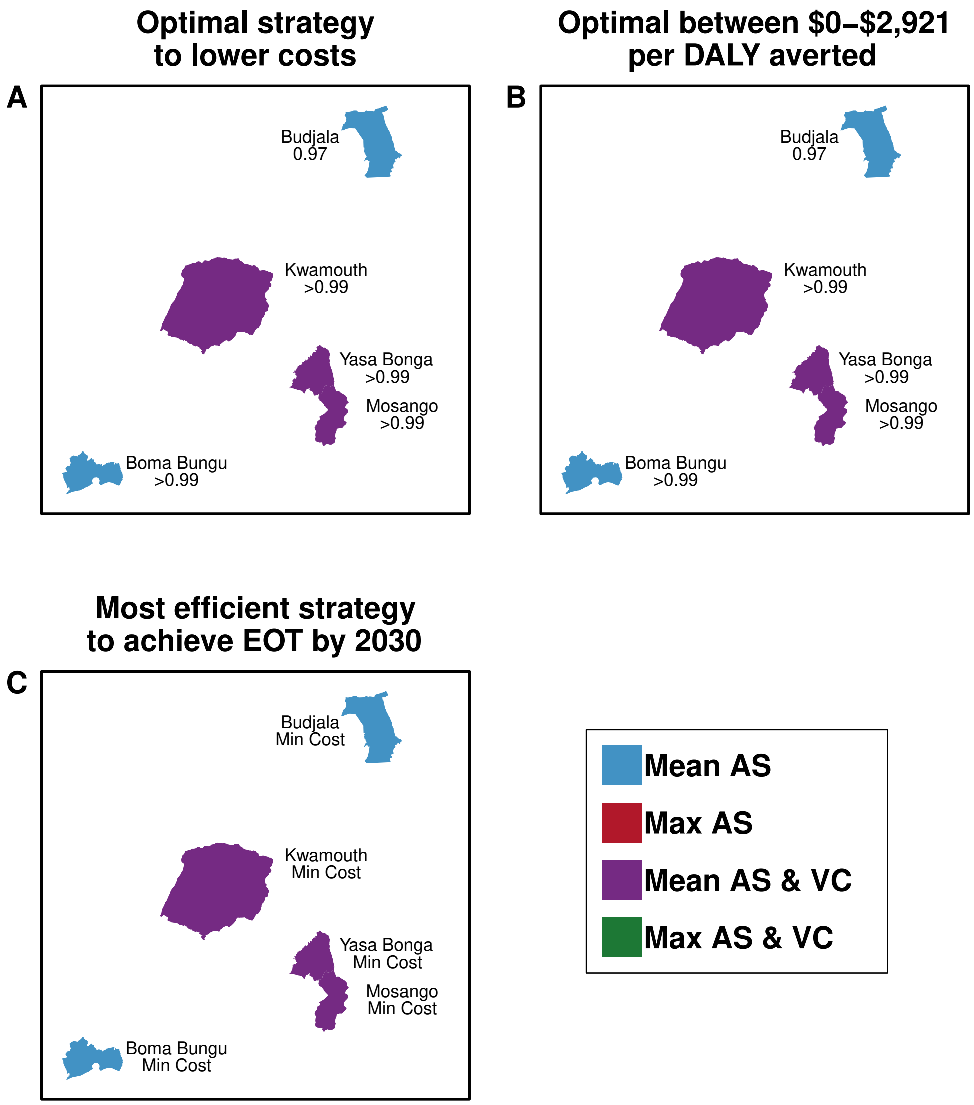


Figure S8: Maps of preferred strategies according to economic or budgetary goals for 2020–2050. Maps A & B show the optimal strategies depending on WTP. The text indicates the probability that the optimal strategy will lead to EOT by 2030. Map C shows the optimal strategy that has at >90% probability of EOT by 2030 and shows the ICER of the indicated strategy (Mean AS for all locations except Yasa Bonga, where it is Mean AS & VC). Maps are not drawn to scale.

#### S3.5 Sensitivity analysis: vector control

| Strategy | VC effectiveness: 60% annual decline of tsetse populations ICER | Pr EOT by 2030 | VC effectiveness: 80% annual decline of tsetse populations ICER | Pr EOT by 2030 | VC effectiveness: 90% annual decline of tsetse populations ICER | Pr EOT by 2030 |
|---|---|---|---|---|---|---|
| **20 targets per kilometer** | | | | | | |
| Mean AS | Min Cost | <0.01 | Min Cost | <0.01 | Dominated | <0.01 |
| Max AS | Dominated | <0.01 | Dominated | <0.01 | Dominated | <0.01 |
| Mean AS & VC | 59 | >0.99 | 4 | >0.99 | Min Cost | >0.99 |
| Max AS & VC | 3,937 | >0.99 | 4,421 | >0.99 | 4,606 | >0.99 |
| **40 targets per kilometer** | | | | | | |
| Mean AS | Min Cost | <0.01 | Min Cost | <0.01 | Min Cost | <0.01 |
| Max AS | Dominated | <0.01 | Dominated | <0.01 | Dominated | <0.01 |
| Mean AS & VC | 343 | >0.99 | 258 | >0.99 | 236 | >0.99 |
| Max AS & VC | 3,873 | >0.99 | 4,373 | >0.99 | 4,548 | >0.99 |

Table S27: Incremental cost-effectiveness ratios (ICERs) under distinct assumptions for vector control

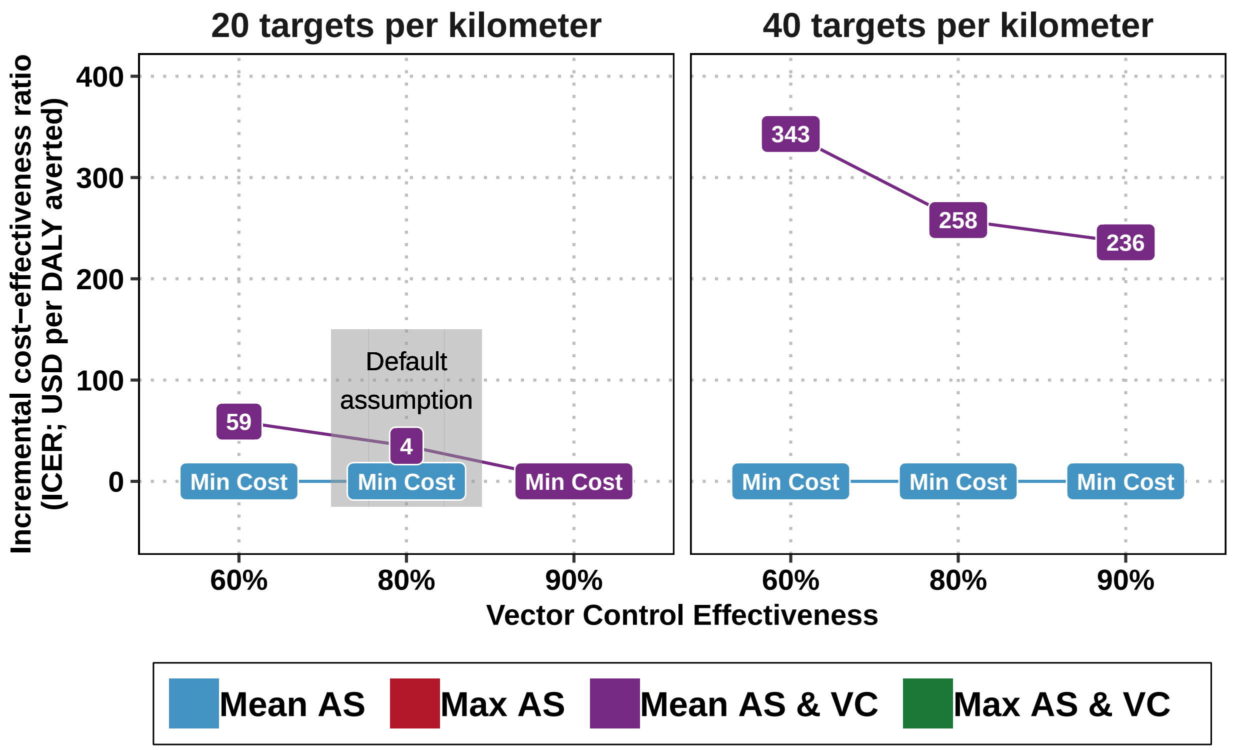


Figure S9: Sensitivity analyses of incremental cost-effectiveness ratios (ICERs) with alternative assumptions of the effectiveness of vector control and the intensity of targets deployed (targets per kilometer) in Kwamouth. In Kwamouth, it is estimated that vector control will be need to be deployed along 390 km of the river Kwa and 42 km of river in Kwamouth East (for a total of 432 km of river). Costs and effects are discounted, and the investment horizon is 2020-2040. Dominated or weakly dominated strategies are not depicted (see section S1.6 for a discussion on weak and strong dominance).

| Strategy | VC effectiveness: 60% tsetse annual decline | | VC effectiveness: 80% tsetse annual decline | | VC effectiveness: 90% tsetse annual decline | |
|---|---|---|---|---|---|---|
| | ICER | Pr EOT by 2030 | ICER | Pr EOT by 2030 | ICER | Pr EOT by 2030 |
| **100 kilometers; 20 targets per kilometer** | | | | | | |
| Mean AS | Dominated | 0.79 | Dominated | 0.79 | Dominated | 0.79 |
| Max AS | Dominated | 0.92 | Dominated | 0.92 | Dominated | 0.92 |
| Mean AS & VC | Min Cost | >0.99 | Min Cost | >0.99 | Min Cost | >0.99 |
| Max AS & VC | 11,245 | >0.99 | 12,444 | >0.99 | 14,224 | >0.99 |
| **210 kilometers; 20 targets per kilometer** | | | | | | |
| Mean AS | Min Cost | 0.79 | Min Cost | 0.79 | Min Cost | 0.79 |
| Max AS | Dominated | 0.92 | Dominated | 0.92 | Dominated | 0.92 |
| Mean AS & VC | 480 | >0.99 | 218 | >0.99 | 142 | >0.99 |
| Max AS & VC | 10,815 | >0.99 | 12,017 | >0.99 | 13,720 | >0.99 |
| **100 kilometers; 40 targets per kilometer** | | | | | | |
| Mean AS | Dominated | 0.79 | Dominated | 0.79 | Dominated | 0.79 |
| Max AS | Dominated | 0.92 | Dominated | 0.92 | Dominated | 0.92 |
| Mean AS & VC | Min Cost | >0.99 | Min Cost | >0.99 | Min Cost | >0.99 |
| Max AS & VC | 11,009 | >0.99 | 12,215 | >0.99 | 13,940 | >0.99 |
| **210 kilometers; 40 targets per kilometer** | | | | | | |
| Mean AS | Min Cost | 0.79 | Min Cost | 0.79 | Min Cost | 0.79 |
| Max AS | Dominated | 0.92 | Dominated | 0.92 | Dominated | 0.92 |
| Mean AS & VC | 2,051 | >0.99 | 1,626 | >0.99 | 1,488 | >0.99 |
| Max AS & VC | 10,320 | >0.99 | 11,535 | >0.99 | 13,124 | >0.99 |

Table S28: Incremental cost-effectiveness ratios (ICERs) under different assumptions for vector control in Mosango. Costs and effects were discounted, and the investment horizon is 2020-2040.

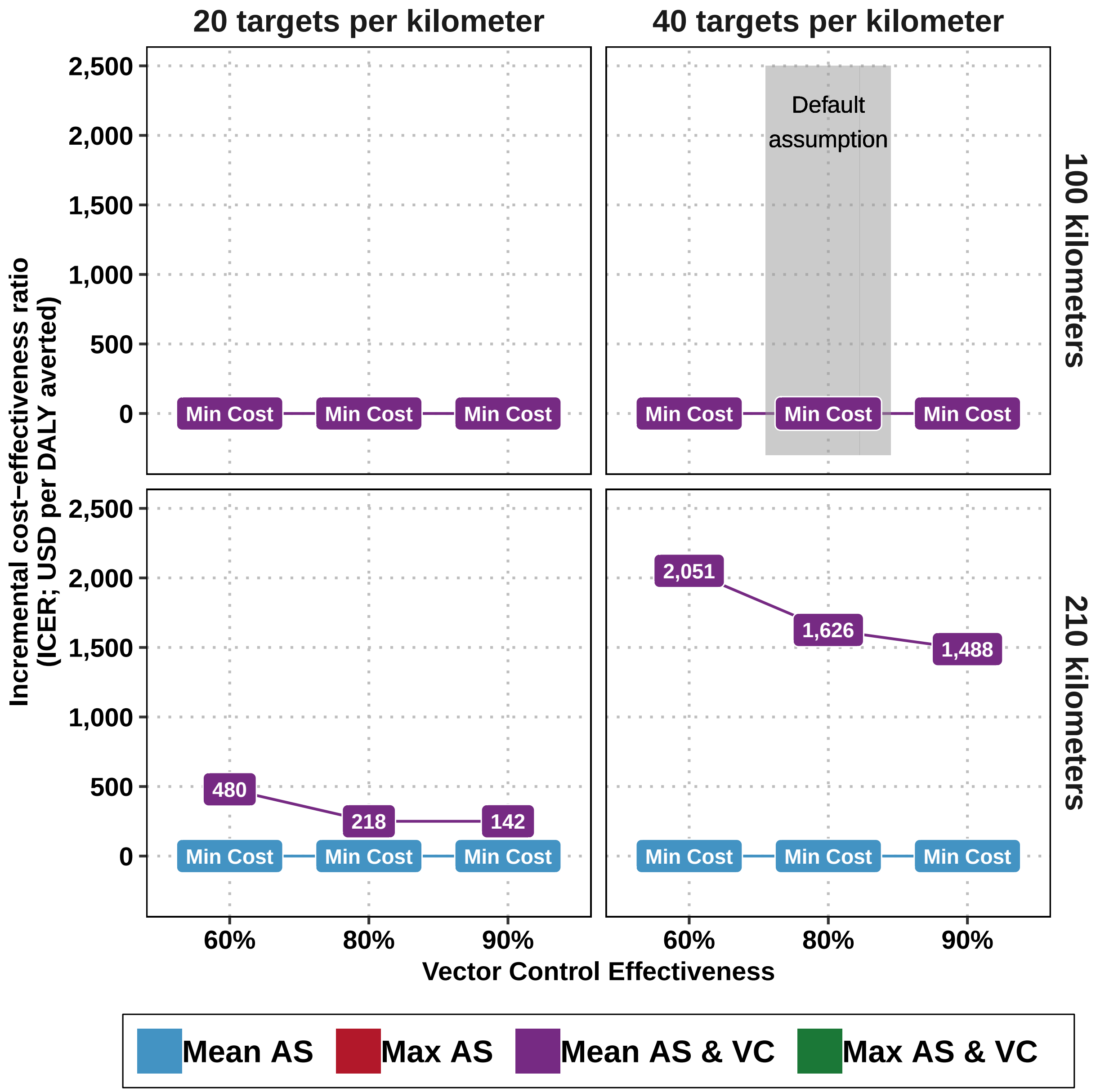


Figure S10: Sensitivity analyses of incremental cost-effectiveness ratios (ICERs) with alternative assumptions of the effectiveness of vector control, the extent to which the health zone must have vector control deployed, and the intensity of targets deployed (targets per kilometer) in Mosango. Costs and effects are discounted, and the investment horizon is 2020-2040. Dominated or weakly dominated strategies are not included in this figure.

### S4 PRIME-NTD criteria

| **Principle and what has been done to satisfy the principle?** | **Where in the manuscript is this described?** |
|---|---|
| **1. Stakeholder engagement**<br>Strategy components were determined along with the country director of PNLTHA, Erick Miaka (co-author). Implementation of simulations of AS and PS costs were aided by Rian Snijders, who has helped run field operations in former Bandundu, and implementation of costs of VC were aided by Rian Snijders and collaborators at the Liverpool School of Tropical Medicine, who have run field operations in DRC since 2015. | Authorship list and acknowledgements |
| **2. Complete model documentation**<br>Full model (including the fitting code) and documentation are available through OpenScienceFramework (OSF). The epidemiological model is fully described in the fitting study of Crump *et al* [1] and cost model in this SI. | Description in SI section S1.2 and access the code via `5HZCEAOSF`[1] and `ProjOSF`[2]. |
| **3. Complete description of data used**<br>Information about the data used for fitting is described in Crump *et al* [1]. The data used for clinical outcomes and costs were estimates from the literature. No data from intervention operations was used. Assumptions and estimates were parameterized according to conventions in the economic evaluation literature [34] | For assumptions around intervention, treatment effects and costs, see supplemental sections S3.2.1, S1.3.1, S2. |
| **4. Communicating uncertainty**<br>*Structural uncertainty*:<br>Uncertainty arising from the choice of vector control operation inputs, discounting, time horizon are shown by re-running the entire analysis with alternative assumptions. | Main text results and discussion. A three-way sensitivity analysis for VC operation is in section S3.5, and time horizons and discounting two-way sensitivity analysis is in section S3.4. |
| *Parameter uncertainty*:<br>The epidemiological parameters are the posterior distributions of a model fitted to time-series data, and full details are available in another publication [1]. For the parameters to model health outcomes and costs, assumptions and estimates were parameterized according to conventions in the economic evaluation literature [34], taking care to sample from large distributions for aspects for which we knew very little. | Epidemiological parameters are available on OSF `FittingOSF` [3]. Health outcome and cost-effectiveness parameters: see SI sections S3.2.1, S1.3.1, S2. Sensitivity analysis for VC in Section S3.5 above and in our `GUI`[4]. |
| *Prediction uncertainty*:<br>Observational uncertainty in epidemiological model predictions. Tables 2 and 3 present both means and 95% prediction intervals. The `GUI`[4] includes box and whisker plots to show uncertainty in cases, deaths and DALYs. We include the probability of meeting EOT by 2030 as well as the expected year of EOT. For cost-effectiveness results, we present the optimal decisions (the probability of each of being cost-effective) at different willingness-to-pay thresholds rather than solely providing ICERs using the net benefits framework (see table 4). | Tables 2-4 of the manuscript and the `GUI`[4]. |
| **5. Testable model outcomes**<br>Epidemiological model outputs are routinely reported metrics of the disease course: detected active and passive case detections. Therefore, these predictions can be compared to future data as long as the data is put into context alongside measures of active screening coverage and the number of fixed health posts equipped for passive surveillance. Some components of costs predictions can be validated against expenditures, but it must be noted that these are economic costs, and so resource use for which there is no explicit invoice is taken into account as well. | Epidemiological projections for the period 2020-2040 are shown in table 3, and for the period ending in 2030 in table S18 and for the period ending in 2050 in table S19. Epidemiological outcomes for each year until 2050 can be viewed in the `GUI`[4]. All the ingredients to the economic costs were shown in detail in the supplement section S3.2.1. |

[1] `5HZCEAOSF` with full address: `https://osf.io/xbwte/`.
[2] `ProjOSF` with full address: `https://osf.io/jza27/`.
[3] `FittingOSF` with full address: `https://osf.io/ck3tr/`.
[4] `GUI` with full address: `https://hatmepp.warwick.ac.uk/5HZCEA/v2/`

Table S29: PRIME-NTD criteria fulfilment. We summarise how the NTD Modelling Consortium's "5 key principles of good modelling practice" have been met in the present study.

### S5 CHEERS checklist

| Section/item | Item No | Recommendation | Reported on page no, line no |
|---|---|---|---|
| Title | 1 | Identify the study as an economic evaluation or use more specific terms such as “cost-effectiveness analysis”, and describe the interventions compared. | Cover page, no line number |
| Abstract | 2 | Provide a structured summary of objectives, perspective, setting, methods (including study design and inputs), results (including base case and uncertainty analyses), and conclusions. | Abstract |
| Background and Objectives | 3 | Provide an explicit statement of the broader context for the study. Present the study question and its relevance for health policy or practice decisions. | Introduction section, last paragraph in particular. |
| Target population and subgroups | 4 | Describe characteristics of the base case population and subgroups analysed, including why they were chosen. | First paragraph of the results. Ordinarily we would put this in the methods, but the occurrence of the results before the methods made the results a more useful place to put this portion. |
| Settings and location | 5 | State relevant aspects of the system(s) in which the decision(s) need(s) to be made. | Second subsection of the methods, "Strategies". Illustrated in figure S1. |
| Study perspective | 6 | Describe the perspective of the study and relate this to the costs being evaluated. | Fifth subsection of the methods, "Costs". |
| Comparators | 7 | Describe the interventions or strategies being compared and state why they were chosen. | Second subsection of the methods, "Strategies", and figure 1. Supplemental section: S1.2.2. |
| Time horizon | 8 | State the time horizon(s) over which costs and consequences are being evaluated and say why appropriate. | First paragraph of the ’Cost-effectiveness’ subsection of the results. Ordinarily we would put this in the methods, but the occurrence of the results before the methods made the results a more useful place to put this portion. |
| Discount rate | 9 | Report the choice of discount rate(s) used for costs and outcomes and say why appropriate. | First paragraph of the ’Cost-effectiveness’ subsection of the results. Ordinarily we would put this in the methods, but the occurrence of the results before the methods made the results a more useful place to put this portion. 3% is the recommended rate by WHO-CHOICE and the Gates Reference Case. |
| Choice of health outcomes | 10 | Describe what outcomes were used as the measure(s) of benefit in the evaluation and their relevance for the type of analysis performed. | Fourth subsection of the methods, "Cases, deaths, and treatment outcome model". Supplemental sections S1.3, S1.2.3 contain detailed explanations of how DALYs are calculated and how the natural history of HAT was considered. |

*(continued)*

| Section/item | Item No | Recommendation | Reported on page no, line no |
|---|---|---|---|
| Measurement of effectiveness | 11a | *Single study-based estimates:* Describe fully the design features of the single effectiveness study and why the single study was a sufficient source of clinical effectiveness data. | NA |
| | 11b | *Synthesis-based estimates:* Describe fully the methods used for identification of included studies and synthesis of clinical effectiveness data. | Effectiveness of AS strategies and PS strategies: model-based, of VC: sensitivity analysis, of screening tests and treatment: the literature. Described in detail in sections S2, S3, and S2. |
| Measurement and valuation of preference based outcomes | 12 | If applicable, describe the population and methods used to elicit preferences for outcomes. | NA |
| Estimating resources and costs | 13a | *Single study-based economic evaluation:* Describe approaches used to estimate resource use associated with the alternative interventions. Describe primary or secondary research methods for valuing each resource item in terms of its unit cost. Describe any adjustments made to approximate to opportunity costs. | NA |
| | 13b | *Model-based economic evaluation:* Describe approaches and data sources used to estimate resource use associated with model health states. Describe primary or secondary research methods for valuing each resource item in terms of its unit cost. Describe any adjustments made to approximate to opportunity costs. | The construction of program costs are detailed in section S1.4, in pages S10-S21. As can be seen, in a publication of this scope, detailing the cost inputs in the main body would be unfeasible. However, the resulting expected costs are in table 3 of the main body of the paper. |
| Currency, price date, and conversion | 14 | Report the dates of the estimated resource quantities and unit costs. Describe methods for adjusting estimated unit costs to the year of reported costs if necessary. Describe methods for converting costs into a common currency base and the exchange rate. | Our general approach for this is in the section "Principles for parameterization" in page S57, section S6.2. The specific choices for each parameter are in SI sectionS6.6, pages S53-S86. |
| Choice of model | 15 | Describe and give reasons for the specific type of decision- analytical model used. Providing a figure to show model structure is strongly recommended. | The decision analytic model is discussed in section S1.5 and illustrated in Figure S2, but the components models feeding into the decision analytic model are described as follows: the dynamic transmission (SEIRS) model is described briefly in section S1.2.1, and the treatment model is shown in figure 4 and described in Section S1.2.3. |
| Assumptions | 16 | Describe all structural or other assumptions underpinning the decision-analytical model. | Sections 2 and 3 of the method section and all of the supplemental methods. |

*(continued)*

| Section/item | Item No | Recommendation | Reported on page no, line no |
|---|---|---|---|
| Analytical methods | 17 | Describe all analytical methods supporting the evaluation. This could include methods for dealing with skewed, missing, or censored data; extrapolation methods; methods for pooling data; approaches to validate or make adjustments (such as half cycle corrections) to a model; and methods for handling population heterogeneity and uncertainty. | Section S1.5 and the parameter glossary, Section S6. |
| Study parameters | 18 | Report the values, ranges, references, and, if used, probability distributions for all parameters. Report reasons or sources for distributions used to represent uncertainty where appropriate. Providing a table to show the input values is strongly recommended. | Section S6. |
| Incremental costs and outcomes | 19 | For each intervention, report mean values for the main categories of estimated costs and outcomes of interest, as well as mean differences between the comparator groups. If applicable, report incremental cost-effectiveness ratios. | Table 3 in the main body, and tables S18 and S19 for alternative time horizons. |
| Characterising uncertainty | 20a | Single study-based economic evaluation: Describe the effects of sampling uncertainty for the estimated incremental cost and incremental effectiveness parameters, together with the impact of methodological assumptions (such as discount rate, study perspective). | NA |
| | 20b | Model-based economic evaluation: Describe the effects on the results of uncertainty for all input parameters, and uncertainty related to the structure of the model and assumptions. | Table 4 in the main body, and tables S25 and S26 for alternative time horizons. In addition, cost-effectiveness acceptability curves are depicted in supplementary figures S5 and S6. The interpretation is in Section 2.4.2 of the results (entitled "Cost-effectiveness in the presence of probabilitic uncertainty") and in the sixth paragraph of the discussion. |
| Characterising heterogeneity | 21 | If applicable, report differences in costs, outcomes, or cost-effectiveness that can be explained by variations between subgroups of patients with different baseline characteristics or other observed variability in effects that are not reducible by more information. | Results section. The analysis is framed as 5 case studies to show the impacts of heterogeneity. |

*(continued)*

| Section/item | Item No | Recommendation | Reported on page no, line no |
|---|---|---|---|
| Study findings, limitations, generalisability, and current knowledge | 22 | Summarise key study findings and describe how they support the conclusions reached. Discuss limitations and the generalisability of the findings and how the findings fit with current knowledge. | Paragraphs 1-4 and 6-8 of the discussion. |
| Source of funding | 23 | Describe how the study was funded and the role of the funder in the identification, design, conduct, and reporting of the analysis. Describe other non-monetary sources of support. | Last section of the methods, Section 4.7, and Acknowledgements section. |
| Conflicts of interest | 24 | Describe any potential for conflict of interest of study contributors in accordance with journal policy. In the absence of a journal policy, we recommend authors comply with International Committee of Medical Journal Editors recommendations. | Conflict of interest statement. |

### S6 Parameter Glossary

S6.1 Organization of parameters . . . S55
S6.2 Principles for parameterization . . . S55
S6.3 Summary of health outcome parameters . . . S56
S6.4 Summary of cost parameters . . . S61
S6.5 Notes on effect parameters . . . S65
S6.5.1 Surveillance parameters . . . S65
S6.5.2 Population . . . S65
S6.5.3 Passive surveillance: coverage of the population per facility . . . S65
S6.5.4 Passive surveillance: number of facilities . . . S66
S6.5.5 Active screening: coverage . . . S66
S6.5.6 Active screening: coverage, enhanced . . . S66
S6.5.7 Active screening: maximum capacity by each traditional team . . . S66
S6.5.8 CATT algorithm: diagnostic specificity . . . S67
S6.5.9 RDT algorithm: diagnostic sensitivity . . . S67
S6.5.10 RDT algorithm: diagnostic specificity . . . S67
S6.5.11 CATT algorithm: wastage in the context of active screening . . . S67
S6.5.12 RDT algorithm: wastage in the context of passive surveillance . . . S67
S6.5.13 Treatment parameters . . . S68
S6.5.14 Percent cases under 6 as a proportion of all cases . . . S68
S6.5.15 Percent cases under 35 kg as a proportion of all cases over the age of 6 . . . S68
S6.5.16 Percent cases of severe stage 2 (S2) as a proportion of cases in stage 2 (S2) . . . S70
S6.5.17 Age of death from infection (for years of life lost per fatal case) . . . S71
S6.5.18 Length of hospital stay: NECT treatment . . . S72
S6.5.19 Length of hospital stay: fexinidazole treatment for stage 1 or 2 disease . . . S73
S6.5.20 Probability of relapse (treatment failure): pentamidine for S1 disease . . . S73
S6.5.21 Probability of relapse (treatment failure): NECT for S2 disease . . . S73
S6.5.22 Probability of relapse (treatment failure): fexinidazole for S1 disease . . . S74
S6.5.23 Probability of relapse (treatment failure): fexinidazole for late S2 disease . . . S75
S6.5.24 Serious adverse events (SAE): pentamidine treatment . . . S75
S6.5.25 Serious adverse events (SAE): NECT treatment . . . S76
S6.5.26 Serious adverse events (SAE): fexinidazole treatment . . . S76
S6.5.27 Days lost to disability: due to stage 1 (S1) disease . . . S77
S6.5.28 Days lost to disability: due to stage 2 (S2) disease . . . S77
S6.5.29 Days lost to disability: due to a serious adverse event (SAE) . . . S77
S6.5.30 Life-years lost (DALY) parameters . . . S77
S6.5.31 Life expectancy . . . S77
S6.5.32 Disability weights: stage 1 (S1) disease . . . S78
S6.5.33 Disability weights: stage 2 (S2) disease . . . S78
S6.5.34 Disability weights: serious adverse events (SAE) . . . S78
S6.5.35 Vector control parameters . . . S79
S6.5.36 Vector control: linear kilometers targets . . . S79
S6.5.37 Vector control: units per kilometer . . . S79
S6.5.38 Vector control: replacement rate of each unit . . . S79
S6.6 Notes on cost parameters . . . S79
S6.6.1 Surveillance cost parameters . . . S79
S6.6.2 Active screening: capital costs of a traditional team . . . S79
S6.6.3 Active screening: fixed management costs of a traditional team . . . S80
S6.6.4 CATT algorithm: cost per test used . . . S80
S6.6.5 Lumbar puncture and laboratory exam: cost per person that needs to be staged . . . S81
S6.6.6 Microscopy (Blood sample, LNA, mAECT): cost per person that needs to be confirmed . . . S81
S6.6.7 RDT algorithm: costs per test used . . . S81
S6.6.8 Variable management costs (PNLTHA mark-up) . . . S82

S6.6.9 Passive surveillance: capital costs of a facility . . . . . . S82
S6.6.10 Passive surveillance: fixed recurrent management costs . . . . . . S83
S6.6.11 Treatment cost parameters . . . . . . S83
S6.6.12 Hospital stay: cost per day . . . . . . S83
S6.6.13 Outpatient consultation: cost . . . . . . S83
S6.6.14 Course of pentamidine: cost . . . . . . S84
S6.6.15 Course of nifurtimox eflornithine combination therapy (NECT): cost . . . . . . S84
S6.6.16 Course of fexinidazole: cost . . . . . . S84
S6.6.17 Drug delivery mark-up . . . . . . S85
S6.6.18 Vector cost parameters . . . . . . S85
S6.6.19 Vector control: operational cost per kilometer of riverbank covered . . . . . . S85
S6.6.20 Vector control: deployment cost per target . . . . . . S85

#### S6.1 Organization of parameters

The code makes use of five r lists: surv_cost, surv_effect, treat_effect, treat_cost, vector_effect, vector_cost. Within those lists sit a set of variables described in the tables later in this document.

- surv_effect: these are the parameters related to the consumption (units) and effectiveness of screening and passive surveillance.
- surv_cost: these are the costs of active screening and passive surveillance. The units are either general demographic units (cases or population) or parameters found in surv_effect.
- treat_effect: these are the parameters related to the consumption (units) and effectiveness of treatment.
- treat_cost: these are the costs of treatment. The units are either general demographic units (cases or population) or parameters found in treat_effect.
- vector_effect: these are the parameters related to the consumption (units) and effectiveness of treatment.
- vector_cost: these are the costs of treatment. The units are either general demographic units (cases or population) or parameters found in treat_effect.

Additionally, a list named epi_output holds the output from the dynamic model: stage 1 and stage 2 cases detected by passive and active surveillance.

#### S6.2 Principles for parameterization

These are the rules I have for myself to parameterize my models. The rationale behind some of the items in my tables are easier to understand with these rules.

- **Transferability of costs across time** Costs have been updated to 2018 USD values. All costs are converted to local currency units in the year of the study, inflated to 2018 values using the consumer price index (CPI) of the country, and then converted to USD using the exchange rate in 2018. It should be noted that the 2003 WHO Guide to Cost-effectiveness recommends that the GDP inflator be used (see 3.2.6 Transferability of costs across time, page 43) but we found that the data on this measure (from the World Bank) were sometimes sparse so we relied on the consumer price index instead [1].

- **Transferability of costs across settings** To ‘borrow’ data from other countries, we follow the 2003 WHO Guide to Cost-effectiveness recommendations in section 3.2.7 Transferability of costs across settings) [1]. For non-traded items (nurse and doctor time) we convert USD or LCU prices into PPP (international dollars) values in the year of the cost study and then turn the value in international dollars to local currency (still in the year of the study) of the country where a cost estimate is needed. Then, we use the CPI to inflate costs to 2018 levels and then use the exchange rate with USD to get 2018 USD values.

- **Combining multiple sources of information** Values from different publications are combined using meta-analytic methods, as described, in sections S6.5 and S6.6.

- **Choice of probability distributions** Costs and ratios were modeled via gamma distributions and proportions were modeled with beta distributions. These distributions were parameterized using the method of moments (see Briggs 2006, Chapter 4) [2].

- **Missing information on uncertainty: Gamma distributions.**

1. Option A: Whenever uncertainty was missing for a cost or ratio, we assigned the parameter a gamma distribution that would yield credible intervals between half and double the estimate available in the literature.
2. Option B: If at least 2 studies listed a cost, then we take the range of the costs to parameterize a gamma distribution for which the the 95 percent confidence interval (the 2.5th and 97.5th percentile) matches the range of estimates from the literature. For these occasions, we use a method to parameterize gamma distributions using quantiles rather than using the mean and standard error of a sample (method of moments) [3].

- **Missing information on uncertainty: Beta distributions.**

1. Option A: Usually modeled assuming that 100 trials were observed with the **proportion_estimate x 100** as the alpha parameter and **(1-proportion_estimate) x 100** as beta parameter.

2. Option B: If at least 2 studies listed a propability or a proportion, then we take the range of the costs to parameterize a beta distribution by assuming the range matches the 95 percent confidence interval (the 2.5th and 97.5th percentile). We use a method to parameterize Beta distributions using quantiles rather than the mean and standard error of a sample (method of moments) [4].

#### S6.3 Summary of health outcome parameters

Below are all the parameters that model health outcomes, as well as a summary of their characteristics. An extended discussion of our choices is featured in the sections annotated in the table.

| Variable description | Variable name | Statistical Distribution | Descriptive Summary | Notes |
|---|---|---|---|---|
| Population | surv_effect[["pop"]] | Fixed value | Varies by health zone | See section S6.5.2 |
| Passive surveillance: coverage of the population per facility | surv_effect[["ps_coverage"]] | Beta(14, 2094) | 0.007 (0.004, 0.010) | See section S6.5.3 |
| Passive surveillance: number of facilities | surv_effect[["ps_facilities"]] | Fixed value | Varies by health zone | See section S6.5.4 |
| Active screening: coverage | surv_effect[["as_traditional"]] | Fixed value | Varies by health zone | See section S6.5.5 |
| Active screening: coverage, enhanced | surv_effect[["as_traditional_max"]] | Fixed value | Varies by health zone | See section S6.5.6 |
| Active screening: maximum capacity by each traditional team | surv_effect[["as_traditional_capacity"]] | Normal(60000, 10000) | 60,055 (40,448, 79,471) | See section S6.5.7 |
| CATT algorithm: diagnostic specificity | surv_effect[["dx_spec_catt"]] | Beta(31, 2) | 0.94 (0.84, 0.99) | See section S6.5.8 |
| RDT algorithm: diagnostic sensitivity | surv_effect[["dx_sens_rdt"]] | Beta(230, 1) | 1.00 (0.98, 1.00) | See section S6.5.9 |
| RDT algorithm: diagnostic specificity | surv_effect[["dx_spec_rdt"]] | Beta(226, 31) | 0.88 (0.84, 0.92) | See section S6.5.10 |
| CATT algorithm: wastage in the context of active screening | surv_effect[["dx_wastage_catt_as"]] | Beta(8, 92) | 0.08 (0.03, 0.14) | See section S6.5.11 |
| RDT algorithm: wastage in the context of passive surveillance | surv_effect[["dx_wastage_rdt_ps"]] | Beta(1, 99) | 0.01 (<0.01, 0.04) | See section S6.5.12 |

Table S31: Surveillance parameters

| Variable description | Variable name | Statistical Distribution | Descriptive Summary | Notes |
|---|---|---|---|---|
| Percent cases under 6 as a proportion of all cases | treat_effect[["patient_char"]] [["pc_s1s2_under6]] | Beta(152.53, 2427.9) | 0.06 (0.05, 0.07) | See section S6.5.14 |
| Percent cases under 35 kg as a proportion of all cases over the age of 6 | treat_effect[["patient_char"]] [["pc_s1s2_under35kg]] | Beta(8.3, 359.6) | 0.02 (<0.01, 0.04) | See section S6.5.15 |
| Percent cases of severe stage 2 (S2) as a proportion of cases in stage 2 (S2) | treat_effect[["patient_char"]] [["pc_s2_late]] | Beta(76.93, 44.87) | 0.63 (0.54, 0.72) | See section S6.5.16 |
| Age of death from infection (for years of life lost per fatal case) | treat_effect[["yld_yll"]] [["aoi_s1s2_years]] | Gamma(148, 0.18) | 26.63 (22.41, 31.08) | See section S6.5.17 |
| Length of hospital stay: NECT treatment | treat_effect[["length_hosp_stay"]] [["lhs_s2_nect_days]] | Fixed value | 10 | See section S6.5.18 |
| Length of hospital stay: fexinidazole treatment for stage 1 or 2 disease | treat_effect[["length_hosp_stay"]] [["lhs_s1s2_fexinidazole_days]] | Fixed value | 10 | See section S6.5.19 |
| Probability of relapse (treatment failure): pentamidine for S1 disease | treat_effect[["treatment_failure"]] [["tf_prob_s1_pentamidine]] | Beta(50.3, 665.48) | 0.07 (0.05, 0.09) | See section S6.5.20 |
| Probability of relapse (treatment failure): NECT for S2 disease | treat_effect[["treatment_failure"]] [["tf_prob_s2_nect]] | Beta(15.87, 378.55) | 0.05 (0.02, 0.08) | See section S6.5.21 |
| Probability of relapse (treatment failure): fexinidazole for S1 disease | treat_effect[["treatment_failure"]] [["tf_prob_s1s2_fexinidazole]] | Beta(9.49, 496.54) | 0.02 (<0.01, 0.03) | See section S6.5.22 |
| Probability of relapse (treatment failure): fexinidazole for late S2 disease | treat_effect[["treatment_failure"]] [["tf_prob_s2late_fexinidazole]] | Beta(43.56, 300.11) | 0.13 (0.09, 0.16) | See section S6.5.23 |
| Serious adverse events (SAE): pentamidine treatment | treat_effect[["sae"]] [["sae_prob_s1_pentamidine]] | Beta(1.43, 551.42) | 0.002 (<0.001, 0.008) | See section S6.5.24 |
| Serious adverse events (SAE): NECT treatment | treat_effect[["sae"]] [["sae_prob_s2_nect]] | Beta(40.88, 367.8) | 0.05 (0.03, 0.08) | See section S6.5.25 |
| Serious adverse events (SAE): fexinidazole treatment | treat_effect[["sae"]] [["sae_prob_s1s2_fexinidazole]] | Beta(3, 261) | 0.01 (<0.01, 0.03) | See section S6.5.26 |
| Days lost to disability: due to stage 1 (S1) disease | treat_effect[["yld_yll"]] [["yld_s1_days]] | Gamma(21.89, 26.07) | 569.21 (355.50, 831.29) | See section S6.5.27 |
| Days lost to disability: due to stage 2 (S2) disease | treat_effect[["yld_yll"]] [["yld_s2_days]] | Gamma(22.18, 12.38) | 275.57 (172.46, 401.20) | See section S6.5.28 |
| Days lost to disability: due to a serious adverse event (SAE) | treat_effect[["yld_yll"]] [["sae_yld_s1s2_days]] | Gamma(1.22, 2.38) | 2.96 (0.14, 9.99) | See section S6.5.29 |

Table S32: Treatment parameters

| Variable description | Variable name | Statistical Distribution | Descriptive Summary | Notes |
|---|---:|---:|---:|---:|
| Life expectancy | disability[["le_years"]] | Fixed value | 60.02 | See section S6.5.31 |
| Disability weights: stage 1 (S1) disease | disability[["dw_s1"]] | Beta(22.96, 147.21) | 0.14 (0.09, 0.19) | See section S6.5.32 |
| Disability weights: stage 2 (S2) disease | disability[["dw_s2"]] | Beta(18.37, 15.63) | 0.54 (0.37, 0.70) | See section S6.5.33 |
| Disability weights: serious adverse events (SAE) | disability[["sae_dw_s1s2"]] | Uniform(0.04, 0.11) | 0.08 (0.04, 0.11) | See section S6.5.34 |

Table S33: Life-years lost (DALY) parameters

| Variable description | Variable name | Statistical Distribution | Descriptive Summary | Notes |
|---|---|---|---|---|
| Vector control: linear kilometers targets | vector_effect[["vc_km"]] | Fixed value | Varies by health zone | See section S6.5.36 |
| Vector control: units per kilometer | vector_effect[["vc_units"]] | Fixed value | Varies by health zone | See section S6.5.37 |
| Vector control: replacement rate of each unit | vector_effect[["vc_deployments_yr"]] | Fixed value | 2 | See section S6.5.38 |

Table S34: Vector control parameters

#### S6.4 Summary of cost parameters

Below are all the cost parameters and a summary of their characteristics in three tables for surveillance, treatment, and vector control costs. An extended discussion of our choices is featured in the sections annotated in the tables.

| Variable description | Variable name | Statistical Distribution | Descriptive Summary | Notes |
|---|---|---|---|---|
| Active screening: capital costs of a traditional team | surv_cost[["as_capital"]] | Gamma(81.02, 114.54) | 9,276 (7,378, 11,375) | See section S6.6.2 |
| Active screening: fixed management costs of a traditional team | surv_effect[["as_recurrent"]] | Gamma(63.31, 630.94) | 39,955 (30,845, 50,435) | See section S6.6.3 |
| CATT algorithm: cost per test used | surv_cost[["dx_catt"]] | Gamma(25.19, 0.02) | 0.52 (0.34, 0.75) | See section S6.6.4 |
| Lumbar puncture and laboratory exam: cost per person that needs to be staged | surv_cost[["dx_lp_lab"]] | Gamma(2.42, 3.66) | 8.90 (1.45, 23.20) | See section S6.6.5 |
| Microscopy (Bloodsample, LNA, mAECT): cost per person that needs to be confirmed | surv_cost[["dx_microscopy1"]] | Gamma(8.47, 1.27) | 10.68 (4.70, 18.84) | See section S6.6.6 |
| RDT algorithm: costs per test used | surv_cost[["dx_rdt"]] | Gamma(8.47, 0.19) | 1.60 (0.71, 2.83) | See section S6.6.7 |
| Variable management costs (PNLTHA mark-up) | surv_cost[["pnltha_mu"]] | Uniform(0.1, 0.2) | 0.15 (0.10, 0.20) | See section S6.6.8 |
| Passive surveillance: capital costs of a facility | surv_effect[["ps_capital"]] | Gamma(8.47, 209.8) | 1,777 (778, 3,157) | See section S6.6.9 |
| Passive surveillance: fixed recurrent management costs | surv_effect[["ps_recurrent"]] | Gamma(8.47, 985.55) | 8,368 (3,743, 14,965) | See section S6.6.10 |

Table S35: Surveillance cost parameters

| Variable description | Variable name | Statistical Distribution | Descriptive Summary | Notes |
|---|---|---|---|---|
| Hospital stay: cost per day | treat_cost[["ip_day"]] | Gamma(5.81, 0.24) | 1.39 (0.50, 2.71) | See section S6.6.12 |
| Outpatient consultation: cost | treat_cost[["op"]] | Uniform(1.37, 3.33) | 2.34 (1.42, 3.28) | See section S6.6.13 |
| Course of pentamidine: cost | treat_cost[["rx_pentamidine"]] | Fixed value | 54 | See section S6.6.14 |
| Course of nifurtimox eflornithine combination therapy (NECT): cost | treat_cost[["rx_nect"]] | Fixed value | 460 | See section S6.6.15 |
| Course of fexinidazole: cost | treat_cost[["rx_fexinidazole"]] | Fixed value | 50 | See section S6.6.16 |
| Drug delivery mark-up | treat_cost[["rx_delivery"]] | Beta(45, 55) | 0.45 (0.35, 0.55) | See section S6.6.17 |

Table S36: Treatment cost parameters

| Variable description | Variable name | Statistical Distribution | Descriptive Summary | Notes |
|---|---|---|---|---|
| Vector control: operational cost per kilometer of riverbank covered | vector_cost[["vc_operation"]] | Gamma(8.47, 14.17) | 120.28 (53.33, 212.26) | See section S6.6.19 |
| Vector control: deployment cost per target | vector_cost[["vc_deploy"]] | Gamma(8.47, 0.54) | 4.57 (2.02, 8.26) | See section S6.6.20 |

Table S37: Vector cost parameters

#### S6.5 Notes on effect parameters

##### S6.5.1 Surveillance parameters

##### S6.5.2 Population

- Name in the code: `surv_effect[["pop"]]`
- Source: [5]
- Country of estimate: DRC
- Statistical distribution and parameters: Fixed value
- Summary statistics (mean and 95% CI or fixed value): Varies by health zone

**Notes**

Population of each health zone. See table 1.

##### S6.5.3 Passive surveillance: coverage of the population per facility

- Name in the code: `surv_effect[["ps_coverage"]]`
- Source: Personal communication with Rian Snijders, ITM Antwerp
- Country of estimate: DRC
- Statistical distribution and parameters: Beta(14, 2094)
- Summary statistics (mean and 95% CI or fixed value): 0.007 (0.004, 0.010)

**Notes**

In personal communications with Rian Snijders (ITM-Antwerp) we determined that each facility would screen about 3-10 people per thousand in a health zone, so health zones with more facilities screen more people than health zones with fewer facilities, and health zones that are more densely populated screen more persons per facility than health zones that are more sparsely populated. Using the method of moments, we assigned a beta distribution with mean equal to 0.0065 (rounded to 0.007 in the summary) and with standard error equal to 0.00175: Beta(14, 2094). The sample therefore has mean and confidence interval: 0.0065 (0.0036, 0.0103).

In the health zones in this study, this would yield the following results:

| Health zone | Population | Number of sites | People screening per site per year (mean) | People screened per health zone per year (mean) |
|---|---|---|---|---|
| Yasa Bonga | 221,917 | 4 | 1,442 | 5,770 |
| Mosango | 125,076 | 1 | 813 | 813 |
| Kwamouth | 131,022 | 5 | 852 | 4,258 |
| Boma Bungu | 85,960 | 2 | 559 | 1,117 |
| Budjala | 133,425 | 2 | 867 | 1,735 |

As a form of crude validation, we note that the Report of the third WHO stakeholder's meeting on gHAT elimination, replicated below, indicated 851 patients screened per center in 2017. We do not know the source population in the areas where the population was screened, but it provides a sense of the number of individuals screened per site per year.

| Year | Number of functioning sites | Total people screened | People screening per site |
|---|---|---|---|
| 2013 | 317 | 275,369 | 869 |
| 2014 | 317 | 271,436 | 856 |
| 2015 | 242 | 277,538 | 1147 |
| 2016 | 198 | 410,035 | 2071 |
| 2017 | 514 | 437,402 | 851 |

###### S6.5.4 Passive surveillance: number of facilities

- Name in the code: `surv_effect[["ps_facilities"]]`
- Source: [6]; see table 1
- Country of estimate: DRC
- Statistical distribution and parameters: Fixed value
- Summary statistics (mean and 95% CI or fixed value): Varies by health zone

**Notes**

These values were retrieved from Simarro et. al. (see the supplement) [6]. These are facilities that can perform serological tests (CATT or RDT), microbiological confirmation, and/or treatment.

###### S6.5.5 Active screening: coverage

- Name in the code: `surv_effect[["as_traditional"]]`
- Source: HAT Atlas data, see table 1
- Country of estimate: DRC
- Statistical distribution and parameters: Fixed value
- Summary statistics (mean and 95% CI or fixed value): Varies by health zone

**Notes**

The percent of the population that is screened by mobile teams in their villages each year. The estimates are available in table 1. These were determined by the average percent of the population in each health zone that was screened over the years 2014-2018.

###### S6.5.6 Active screening: coverage, enhanced

- Name in the code: `surv_effect[["as_traditional_max"]]`
- Source: HAT Atlas data, see table 1
- Country of estimate: DRC
- Statistical distribution and parameters: Fixed value
- Summary statistics (mean and 95% CI or fixed value): Varies by health zone

**Notes**

The percent of the population that is screened by mobile teams in their villages each year assuming that active screening is bolstered to “maximum levels”. The estimates are available in table 1. These were determined by the maximum percent of the population in each health zone that was screened over the years 2000-2018.

###### S6.5.7 Active screening: maximum capacity by each traditional team

- Name in the code: `surv_effect[["as_traditional_capacity"]]`
- Source: [7–9]
- Country of estimate: DRC
- Statistical distribution and parameters: Normal(60000, 10000)
- Summary statistics (mean and 95% CI or fixed value): 60,055 (40,448, 79,471)

**Notes**

The capacity of an active surveillance team in DRC has a mean of 60,000 and a lower bound of 40,000 [7] and an upper bound of 80,000 with a work year of 220 days [9]. Teams are managed by the coordination and serve a span of multiple health zones and dozens of villages.

Capacity can vary by location, so to parameterize the model, we chose a normal distribution with upper and lower bounds of 40-80 thousand people, therefore the parameters are Normal(60,000, 10,000).

##### S6.5.8 CATT algorithm: diagnostic specificity

- Name in the code: `surv_effect[["dx_spec_catt"]]`
- Source: [10]
- Country of estimate: Various
- Statistical distribution and parameters: Beta(31, 2)
- Summary statistics (mean and 95% CI or fixed value): 0.94 (0.84, 0.99)

**Notes**

Based on a systematic review by Mitashi and colleagues [10], who reported ranges of 83.5-99.3%. The beta distribution that corresponds with that range as 95% confidence intervals is Beta(33, 2).

##### S6.5.9 RDT algorithm: diagnostic sensitivity

- Name in the code: `surv_effect[["dx_sens_rdt"]]`
- Source: [11]
- Country of estimate: Guinea and Cote d'Ivoire
- Statistical distribution and parameters: Beta(230, 1)
- Summary statistics (mean and 95% CI or fixed value): 1.00 (0.98, 1.00)

**Notes**

Based on a study in Guinea and Cote d'Ivoire [11], there was 1 sample from a gHAT patient that tested negative out of 231. Therefore, the parameter distribution for specifity is Beta(230, 1).

##### S6.5.10 RDT algorithm: diagnostic specificity

- Name in the code: `surv_effect[["dx_spec_rdt"]]`
- Source: [11]
- Country of estimate: Guinea and Cote d'Ivoire
- Statistical distribution and parameters: Beta(226, 31)
- Summary statistics (mean and 95% CI or fixed value): 0.88 (0.84, 0.92)

**Notes**

Based on a study in Guinea and Cote d'Ivoire [11], there where 31 samples from non-HAT patients that tested positive out of 257. Therefore, the parameter distribution for specifity is Beta(226, 31).

##### S6.5.11 CATT algorithm: wastage in the context of active screening

- Name in the code: `surv_effect[["dx_wastage_catt_as"]]`
- Source: [9]
- Country of estimate: DRC
- Statistical distribution and parameters: Beta(8, 92)
- Summary statistics (mean and 95% CI or fixed value): 0.08 (0.03, 0.14)

**Notes**

CATT tests come in packs of 50, and the list cost is assumed to consider that a pack is used on 50 patients. Once a pack is opened, one test is used as a positive control and one test is used as a negative control, so wastage is at least 4 percent. Shelf life of the test is one week in refrigeration and wastage in active screening activities is relatively low; generally, wastage of CATT tests in the context of active screening occurs at the end of the day when there are tests remaining in an open pack. To be conservative, we doubled the 4-percent lower bound for wastage and assigned the parameter a distribution of Beta(8, 92).

##### S6.5.12 RDT algorithm: wastage in the context of passive surveillance

- Name in the code: `surv_effect[["dx_wastage_rdt_ps"]]`
- Source: [12]
- Country of estimate: DRC

- Statistical distribution and parameters: Beta(1, 99)
- Summary statistics (mean and 95% CI or fixed value): 0.01 (<0.01, 0.04)

**Notes**

We followed the same assumption as Snijders and colleagues that less than 1 percent of RDT tests would not be used [12]. Because there was no sense of uncertainty in this parameter, we assumed a Beta (1,99) distribution.

###### S6.5.13 Treatment parameters

###### S6.5.14 Percent cases under 6 as a proportion of all cases

- Name in the code: `treat_effect[["patient_char"]][["pc_s1s2_under6]]`
- Source: [13, 14]
- Country of estimate: South Sudan
- Statistical distribution and parameters: Beta(152.53, 2427.9)
- Summary statistics (mean and 95% CI or fixed value): 0.06 (0.05, 0.07)

**Notes**

There were only two studies where the number of children under 5 or 6 years of age was stated explicitly [14, 13].

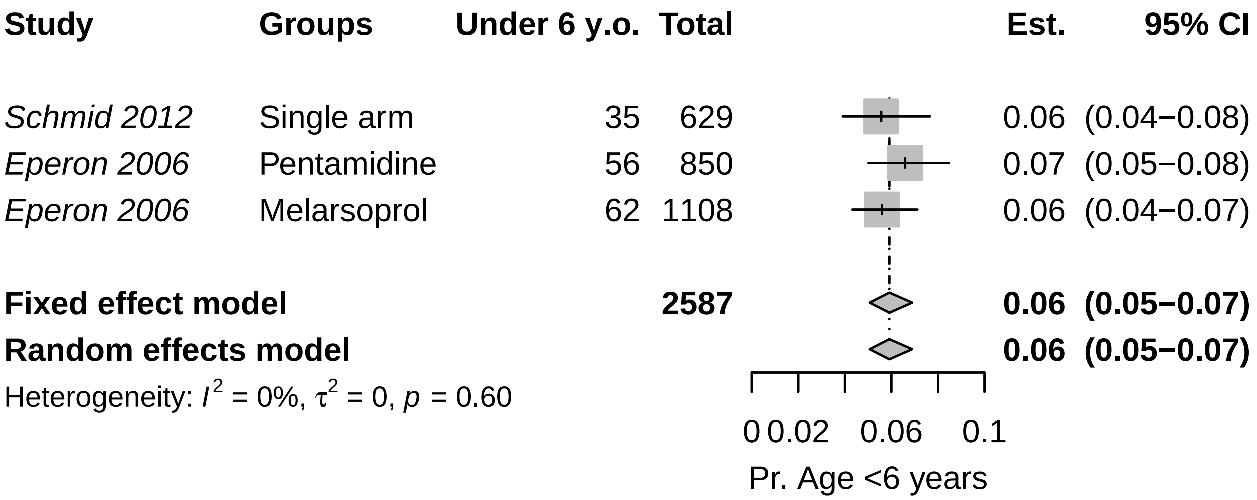


Because the data showed non-significant heterogeneity according to the tau-squared test for heterogeneity, we have chosen to use the fixed-effects combined estimate: 0.059 (0.051, 0.069). The beta parameters of the random-effects estimate Beta(153.53, 2427.90) for the probability of that a patient is under 6 years old.

###### S6.5.15 Percent cases under 35 kg as a proportion of all cases over the age of 6

- Name in the code: `treat_effect[["patient_char"]][["pc_s1s2_under35kg]]`
- Source: [14–23]
- Country of estimate: Various
- Statistical distribution and parameters: Beta(8.3, 359.6)
- Summary statistics (mean and 95% CI or fixed value): 0.02 (<0.01, 0.04)

**Notes**

To determine whether a patient is eligible for fexinidazole treatment, we could not find any studies that would tell us the number of HAT patients who weighed less than 35 kg, but we have estimated the number of people that might weigh less than 35 kg by examining the distribution of weight among patients in the trials in the literature. Furthermore, we have examined how this variable is related to potential selection by age if the study population. We are interested in the proportion of older children and adults that might weigh less than 35 kg, as age under 6 is a contraindication for fexinidazole.

We fit a gamma distribution by the method of moments to the reported mean and standard deviations of each of the studies. For Priotto 2012, no SD was reported, but an inter-quartile range was reported, so we fit a gamma distribution by the method in Cook [3].

We then took the expected number of people under 35 kg, and then performed a single-proportion meta-analysis with the expected number of people in each study under and over the 35 kg-threshold.

| Citation | Group | Age Group | Mean weight | Measure of spread | No. of obser-vations | Gamma distr. alpha par. | Gamma distr. beta par. | Prop. <35kg | Simulated No. <35kg |
|---|---|---|---|---|---|---|---|---|---|
| Priotto 2006 | Melarsoprol and Nifurtimox | All ages | 49.20 | SD = 14.4 | 18 | 11.67 | 4.21 | 0.16 | 3 |
| Priotto 2006 | Melarsoprol and Eflornithine | All ages | 50.00 | SD = 10.3 | 19 | 23.56 | 2.12 | 0.06 | 1 |
| Priotto 2006 | NECT | All ages | 51.40 | SD = 8.4 | 17 | 37.44 | 1.37 | 0.02 | 0 |
| Priotto 2007 | NECT | Over 15 years old | 51.70 | SD = 7.4 | 52 | 48.81 | 1.06 | 0.01 | 0 |
| Priotto 2007 | Eflornithine | Over 15 years old | 53.10 | SD = 7.2 | 51 | 54.39 | 0.98 | 0.00 | 0 |
| Checchi 2007 | NECT | All ages | 44.80 | SD = 15.1 | 31 | 8.80 | 5.09 | 0.28 | 9 |
| Priotto 2009 | NECT | Over 15 years old | 53.00 | SD = 8.7 | 143 | 37.11 | 1.43 | 0.01 | 2 |
| Priotto 2009 | Eflornithine | Over 15 years old | 53.90 | SD = 8.3 | 143 | 42.17 | 1.28 | 0.01 | 1 |
| Ngoyi 2010 | Pentamidine and Melarsoprol | Over 12 years old | 56.00 | SD = 10.0 | 360 | 31.36 | 1.79 | 0.01 | 3 |
| Priotto 2012 | Single arm | All ages | 49.00 | IQR: 40-56 | 2190 | 16.37 | 2.96 | 0.12 | 265 |
| Schmid 2012 | Single arm | All ages | 45.00 | SD = 16.0 | 629 | 7.91 | 5.69 | 0.29 | 182 |
| Burri 2016 | Pentamidine | Over 15 years old and >35 kg | 48.50 | SD = 7.6 | 40 | 40.83 | 1.19 | 0.03 | 1 |
| Pohlig 2016 | Pentamidine | Over 12 years old and >30 kg | 45.70 | SD = 7.8 | 137 | 34.15 | 1.34 | 0.08 | 10 |
| Pohlig 2016 | Pafuramidine | Over 12 years old and >30 kg | 44.70 | SD = 7.9 | 136 | 32.02 | 1.40 | 0.10 | 14 |
| Kansiime 2018 | All | Over 15 years old | 51.69 | SD = 9.7 | 109 | 28.22 | 1.83 | 0.03 | 3 |
| Mesu 2018 | NECT | Over 15 years old | 50.70 | SD = 9.6 | 130 | 27.89 | 1.82 | 0.04 | 5 |
| Mesu 2018 | Fexinidazole | Over 15 years old | 50.50 | SD = 8.2 | 264 | 37.93 | 1.33 | 0.02 | 5 |

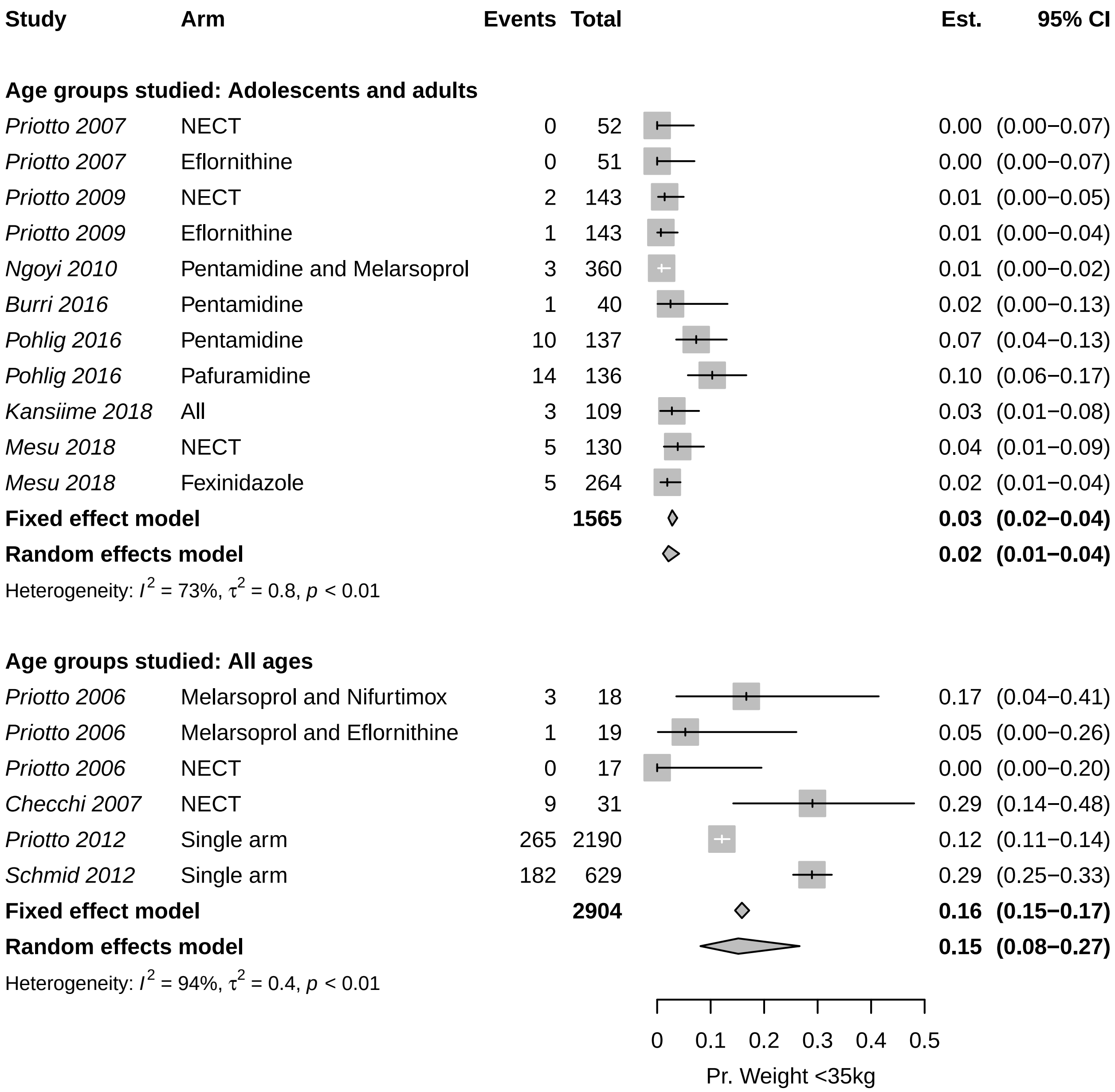


Because the data showed significant heterogeneity according to the tau-squared test for heterogeneity, we have chosen to use the random-effects estimate: 0.02 (0.01-0.04), represented by probability distribution: Beta(8.30, 359.61).

##### S6.5.16 Percent cases of severe stage 2 (S2) as a proportion of cases in stage 2 (S2)

- Name in the code: `treat_effect[["patient_char"]][["pc_s2_late]]`
- Source: [13–19, 24]
- Country of estimate: Various
- Statistical distribution and parameters: Beta(76.93, 44.87)
- Summary statistics (mean and 95% CI or fixed value): 0.63 (0.54, 0.72)

**Notes**

The definition of severe stage 2 gHAT disease by the WHO is when there are more than 100 white blood cells (WBC, leukocytes) per micro-liter in the cerebro-spinal fluid. We have searched the clinical trials for the proportion of stage 2 patients that have high concentrations of leukocytes upon admission to treatment.

| Study | Arm | Events | Total | Est. | 95% CI |
|---|---|---|---|---|---|
| *Priotto 2006* | Melarsoprol and Nifurtimox | 9 | 18 | 0.50 | (0.26−0.74) |
| *Priotto 2006* | Melarsoprol and Eflornithine | 12 | 19 | 0.63 | (0.38−0.84) |
| *Priotto 2006* | NECT | 6 | 17 | 0.35 | (0.14−0.62) |
| *Eperon 2006* | Melarsoprol | 407 | 1108 | 0.37 | (0.34−0.40) |
| *Priotto 2007* | NECT | 31 | 52 | 0.60 | (0.45−0.73) |
| *Priotto 2007* | Eflornithine | 39 | 51 | 0.76 | (0.63−0.87) |
| *Checchi 2007* | NECT | 18 | 31 | 0.58 | (0.39−0.75) |
| *Priotto 2009* | NECT | 107 | 143 | 0.75 | (0.67−0.82) |
| *Priotto 2009* | Eflornithine | 115 | 143 | 0.80 | (0.73−0.87) |
| *Ngoyi 2010* | Pentamidine and Melarsoprol | 209 | 272 | 0.77 | (0.71−0.82) |
| *Priotto 2012* | Single arm | 1373 | 2190 | 0.63 | (0.61−0.65) |
| *Schmid 2012* | Single arm | 397 | 629 | 0.63 | (0.59−0.67) |
| **Fixed effect model** | | | **4673** | **0.58** | **(0.57−0.60)** |
| **Random effects model** | | | | **0.63** | **(0.55−0.71)** |

Heterogeneity: $I^2 = 96\%$, $\tau^2 = 0.3$, $p < 0.01$

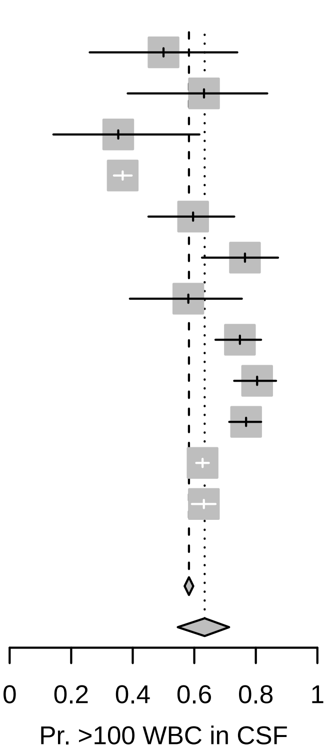


Because the data showed significant heterogeneity according to the tau-squared test for heterogeneity, we have chosen to use the random-effects combined estimate: 0.634 (0.546, 0.713), represented by the probability distribution Beta(76.93, 44.87).

##### S6.5.17 Age of death from infection (for years of life lost per fatal case)

- Name in the code: `treat_effect[["yld_yll"]][["aoi_s1s2_years]]`
- Source: [13–23, 25–27]
- Country of estimate: DRC and South Sudan
- Statistical distribution and parameters: Gamma(148, 0.18)
- Summary statistics (mean and 95% CI or fixed value): 26.63 (22.41, 31.08)

**Notes**

No good registry of age of infection exists, so we have searched through the literature that we have used to parameterize the model for the average age of HAT patients. Among the studies that we have used to inform other parameters, eight studies reported age information in a sample of patients of all ages, and nine studies have reported the age information in a sample of older children or adults (12-15 years and older).

However, the data exists in a state that is difficult to synthesize:

| Citation | Group | Age Group | Summary |
|---|---|---|---|
| Priotto 2006 | Melarsoprol and Nifurtimox | All ages | Mean: 29.1 range: 5-56 |
| Priotto 2006 | Melarsoprol and Eflornithine | All ages | Mean: 28.1 range: 11-61 |
| Priotto 2006 | NECT | All ages | Mean: 29.1 range: 9-62 |
| Balasegaram 2006 | Pentamidine | All ages | 148 under 15 and 504 over 15 |
| Eperon 2006 | Pentamidine | All ages | 56 patients 0-5, 226 patients 6-15, 568 patients 15+ |
| Eperon 2006 | Melarsoprol | All ages | 63 patients 0-5, 249 patients 6-15, 796 patients 15+ |
| Priotto 2007 | NECT | Over 15 years old | Mean: 33.1 range: 15-69 |
| Priotto 2007 | Eflornithine | Over 15 years old | Mean: 36.1 range: 15-70 |
| Checchi 2007 | NECT | All ages | Mean: 23.9 range: 4-45 |
| Priotto 2009 | NECT | Over 15 years old | Mean: 32.8 SD: 12.5 |
| Priotto 2009 | Eflornithine | Over 15 years old | Mean: 34.6 SD: 13.5 |
| Ngoyi 2010 | Pentamidine | Over 12 years old | Mean: 35 SD: 13 |
| Ngoyi 2010 | Pentamidine and Melarsoprol | Over 12 years old | Mean: 34 SD: 12 |
| Priotto 2012 | Single arm | All ages | Median: 24 IQR: 15-35 |
| Schmid 2012 | Single arm | All ages | 35 patients 0-4 yo, 65 patients 5-11 yo, and 529 patients 12 yo or more. |
| Hasker 2012 | All | All ages | Median: 27 IQR: 16-40 |
| Alirol 2013 | Single arm | All ages | Median: 36 IQR: 20-50 |
| Burri 2016 | Pentamidine | Over 15 years old and >35 kg | Median: 31 range: 15-50 |
| Pohlig 2016 | Pentamidine | Over 12 years old and >30 kg | Median: 31 range: 13-75 |
| Pohlig 2016 | Pafuramidine | Over 12 years old and >30 kg | Median: 30 range: 12-64 |
| Kansiime 2018 | NECT | Over 15 years old | Mean: 27.23 SD: 12.07 |
| Kansiime 2018 | Eflornithine | Over 15 years old | Mean: 27.33 SD: 8.59 |
| Mesu 2018 | NECT | Over 15 years old | Mean: 35.2 SD: 13.2 |
| Mesu 2018 | Fexinidazole | Over 15 years old | Mean: 34.5 SD: 12.6 |

Therefore, we have fit a gamma distribution to the means and medians of the studies that included patients of all ages. We have omitted the median from Alirol 2013 as this median seems unusually high – even higher than the mean age of patients in studies were only adults (over the age of 15) were recruited. Our distribution is therefore Gamma(147.93, 0.18) with a mean and 95% confidence interval of 27 (23, 31), which provides a sufficiently large bound of uncertainty in lieu of the information we have.

##### S6.5.18 Length of hospital stay: NECT treatment

- Name in the code: `treat_effect[["length_hosp_stay"]][["lhs_s2_nect_days]]`
- Source: [23]
- Country of estimate: Global recommendations
- Statistical distribution and parameters: Fixed value
- Summary statistics (mean and 95% CI or fixed value): 10

**Notes**

For NECT treatment, patients must stay in inpatient care for a minimum of 7 days for the eflornithine infusions, and whether they stay for a total of 10 days for nifurtimox administration is unclear. For the most recent clinical trial [23], NECT patients were released on days 13-18 after admission, but we have assumed that for the most recent trial the average patient can be released from care after 10 days in the hospital.

##### S6.5.19 Length of hospital stay: fexinidazole treatment for stage 1 or 2 disease

- Name in the code: `treat_effect[["length_hosp_stay"]][["lhs_s1s2_fexinidazole_days]]`
- Source: [23]
- Country of estimate: Global recommendations
- Statistical distribution and parameters: Fixed value
- Summary statistics (mean and 95% CI or fixed value): 10

**Notes**

For the only trial that is published [23], patients were released on days 13-18 after the initiation of treatment, although the treatment only took 10 days, so we have assumed that in routine care that the average patient will be in inpatient care for 10 days.

##### S6.5.20 Probability of relapse (treatment failure): pentamidine for S1 disease

- Name in the code: `treat_effect[["treatment_failure"]][["tf_prob_s1_pentamidine]]`
- Source: [13, 19–21, 25, 28]
- Country of estimate: Various
- Statistical distribution and parameters: Beta(50.3, 665.48)
- Summary statistics (mean and 95% CI or fixed value): 0.07 (0.05, 0.09)

**Notes**

The WHO guidelines for treatment of HAT in 2019 [24] presented existing data on treatment failure of pentamidine treatment. To produce one comprehensive estimate of treatment failure, we performed a meta-analysis on proportions within a single group.

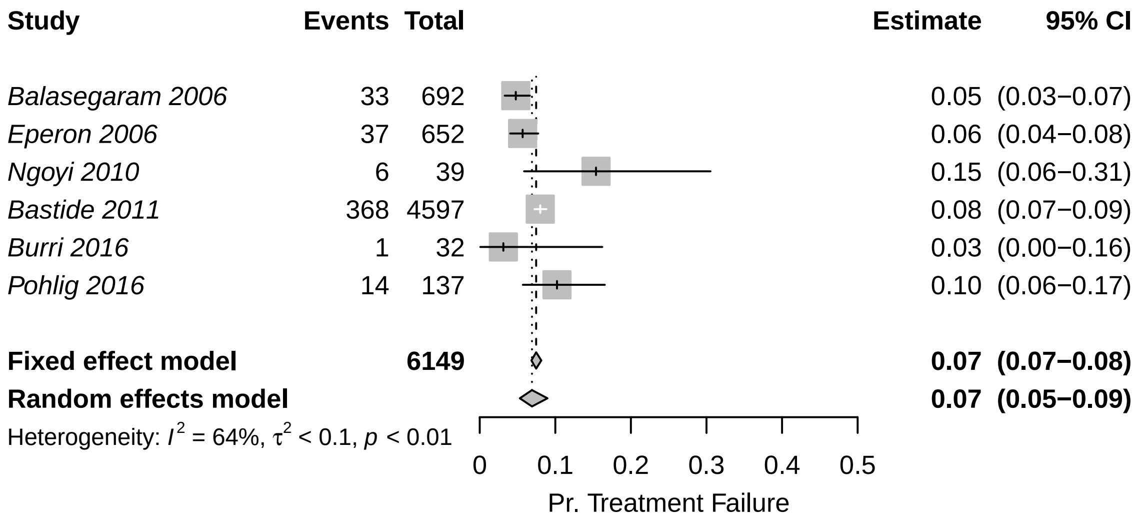


Because the data showed significant heterogeneity according to the tau-squared test for heterogeneity, we have chosen to use the random-effects estimate, to which we assigned a distribution of Beta(50.30, 665.48).

##### S6.5.21 Probability of relapse (treatment failure): NECT for S2 disease

- Name in the code: `treat_effect[["treatment_failure"]][["tf_prob_s2_nect]]`
- Source: [15–18, 22, 23]
- Country of estimate: Various
- Statistical distribution and parameters: Beta(15.87, 378.55)
- Summary statistics (mean and 95% CI or fixed value): 0.05 (0.02, 0.08)

**Notes**

The WHO guidelines for treatment of HAT in 2019 [24] presented existing data on treatment failure of NECT. Kansiime and colleagues [22] also performed a systematic review of studies estimating the outcomes of NECT treatment. To produce one comprehensive estimate of treatment failure, we performed a meta-analysis on proportions within single groups.

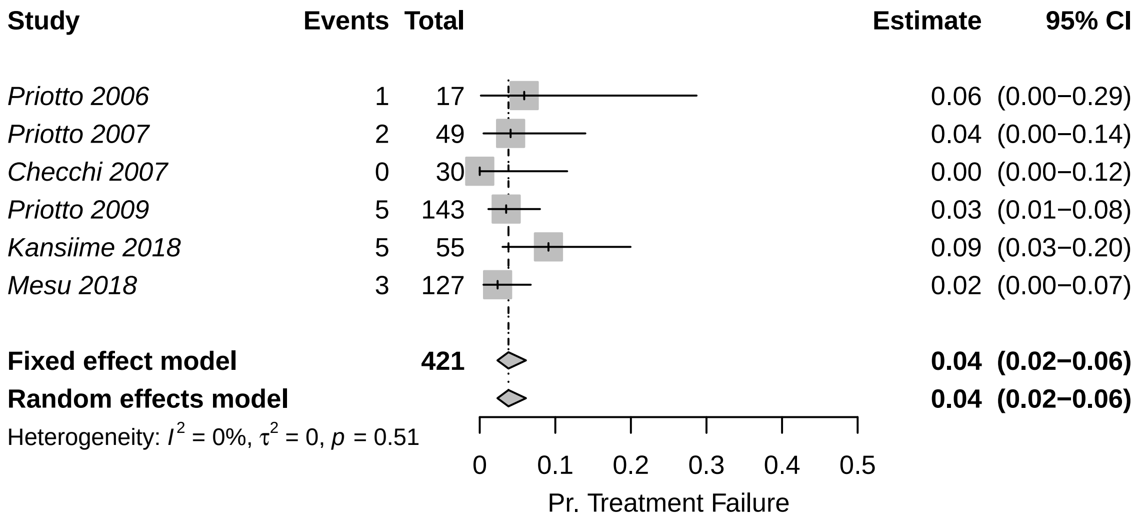


Because the data showed non-significant heterogeneity according to the tau-squared test for heterogeneity, we have chosen to use the fixed-effects estimate, to which we assigned a distribution of Beta(15.87, 378.55)

##### S6.5.22 Probability of relapse (treatment failure): fexinidazole for S1 disease

- Name in the code: `treat_effect[["treatment_failure"]][["tf_prob_s1s2_fexinidazole]]`
- Source: [24]
- Country of estimate: DRC
- Statistical distribution and parameters: Beta(9.49, 496.54)
- Summary statistics (mean and 95% CI or fixed value): 0.02 (<0.01, 0.03)

**Notes**

Mesu and colleagues [23] have published the only study on fexinidazole treatment effectiveness on late-stage 2 cases. Moreover, the accompanying meta-analysis for the WHO treatment guidelines released in 2019 show the outcomes an additional extension study on stage 1, both early and late-stage 2 disease as well we for the data from Mesu et al stratified by the concentration of WBC in the CSF [24].

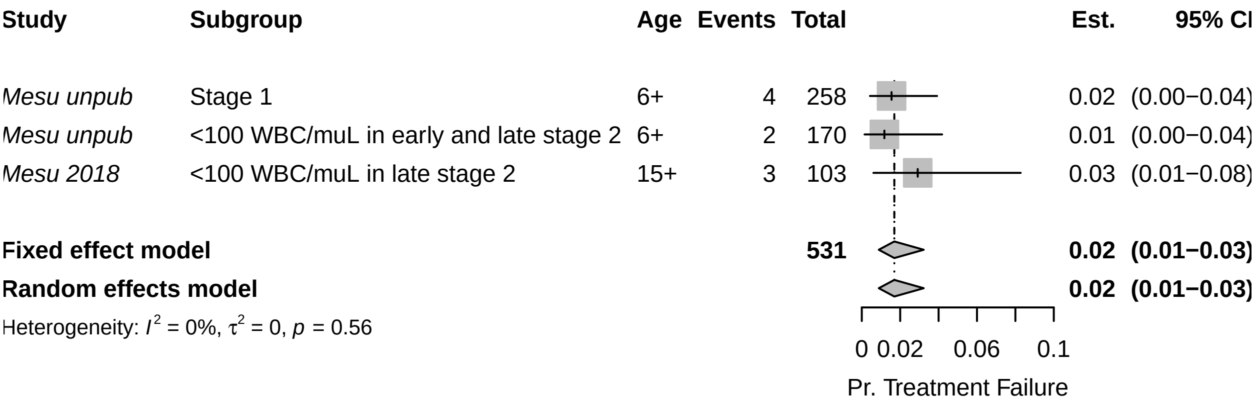


Because the data showed non-significant heterogeneity according to the tau-squared test for heterogeneity, we have chosen to use the fixed-effects combined estimate: 0.017 (0.009, 0.032), for which we assigned a distribution of Beta(9.49, 496.54).

##### S6.5.23 Probability of relapse (treatment failure): fexinidazole for late S2 disease

- Name in the code: `treat_effect[["treatment_failure"]][["tf_prob_s2late_fexinidazole]]`
- Source: [23, 24]
- Country of estimate: DRC and CAR
- Statistical distribution and parameters: Beta(43.56, 300.11)
- Summary statistics (mean and 95% CI or fixed value): 0.13 (0.09, 0.16)

**Notes**

Mesu et al have published the only study on fexinidazole treatment effectiveness on late-stage 2 cases. Moreover, the accompanying meta-analysis for the WHO treatment guidelines released in 2019 show the outcomes an additional extension study on stage 1, both early and late-stage 2 disease ([24]).

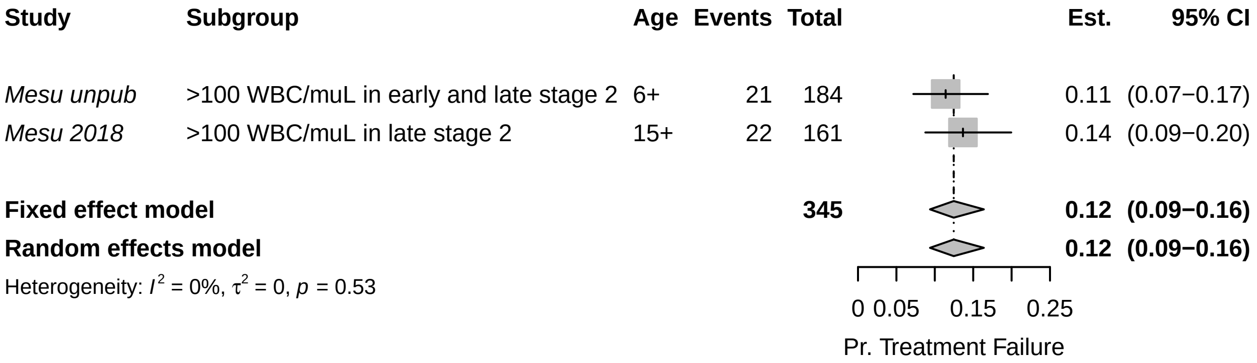


Because the data showed non-significant heterogeneity according to the tau-squared test for heterogeneity, we have chosen to use the fixed-effects combined estimate: 0.125 (0.094, 0.164). The beta parameters of the random-effects estimate Beta(43.56, 300.11) for the probability of severe adverse effects attributable to NECT administration.

##### S6.5.24 Serious adverse events (SAE): pentamidine treatment

- Name in the code: `treat_effect[["sae"]][["sae_prob_s1_pentamidine]]`
- Source: [13, 20, 21]
- Country of estimate: DRC and South Sudan
- Statistical distribution and parameters: Beta(1.43, 551.42)
- Summary statistics (mean and 95% CI or fixed value): 0.002 (<0.001, 0.008)

**Notes**

As part of the WHO guidelines for treatment of HAT in 2019 ([24]), Cochrane performed a systematic review of studies that evaluated the efficacy of NECT compared to fexinidazole studies and presented the probability of serious adverse events. Severe or serious adverse events in studies for S1 treatment were defined as “significant hazard, contra-indication, side effect, or precaution” [13, 20, 21].

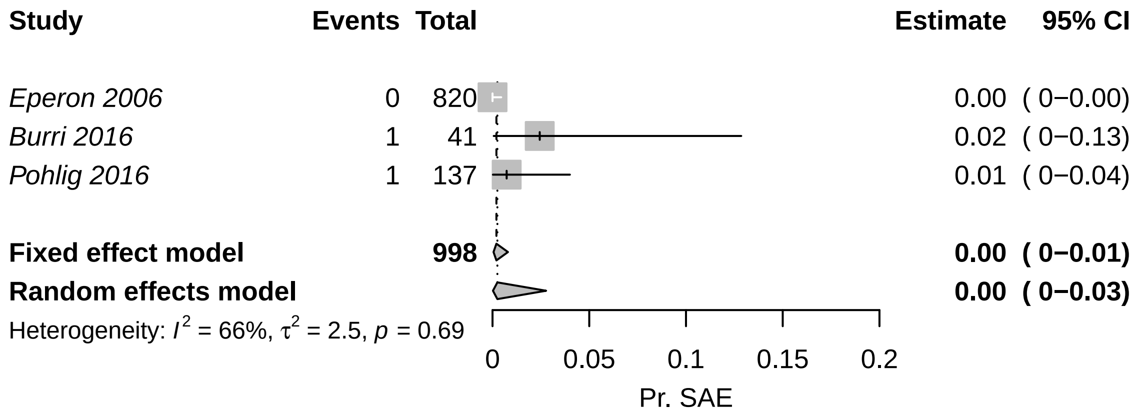

Because the results do not contain evidence of significant heterogeneity, we have chosen to use the fixed (pooled) estimate of 0 (0-0.01), which would result from a beta distribution of Beta(1.43, 551.41).

###### S6.5.25 Serious adverse events (SAE): NECT treatment

- Name in the code: `treat_effect[["sae"]][["sae_prob_s2_nect]]`
- Source: [15–18, 22, 23]
- Country of estimate: DRC
- Statistical distribution and parameters: Beta(40.88, 367.8)
- Summary statistics (mean and 95% CI or fixed value): 0.05 (0.03, 0.08)

**Notes**

The WHO guidelines for treatment of gHAT in 2019 [24], presented all NECT studies to date, as did Kansiime and colleagues [22]. We searched through these studies for evidence of the probability of severe adverse vents (SAEs),descrived as events of Grade 3 or higher according to the National Cancer Institute Common Toxicity Criteria for Adverse Events (CTCAE).

To produce one comprehensive estimate of the probability of SAE, we performed a meta-analysis on proportions within single groups.

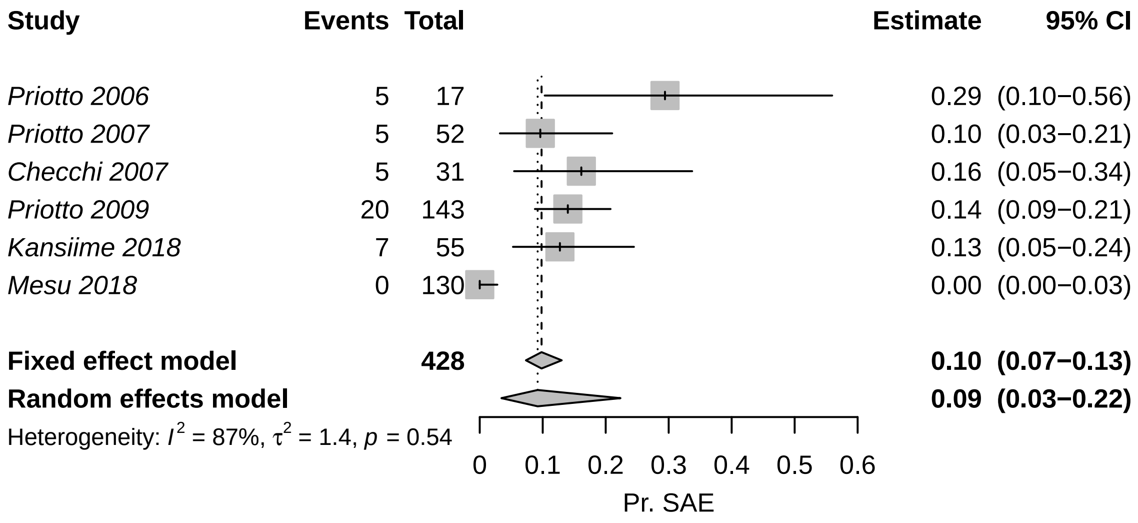


Because the data showed non-significant heterogeneity according to the tau-squared test for heterogeneity, we have chosen to use the fixed-effects estimate: 0.098 (0.073, 0.130), which would result from a beta distribution of Beta(40.88, 367.80).

###### S6.5.26 Serious adverse events (SAE): fexinidazole treatment

- Name in the code: `treat_effect[["sae"]][["sae_prob_s1s2_fexinidazole]]`
- Source: [23]
- Country of estimate: DRC
- Statistical distribution and parameters: Beta(3, 261)
- Summary statistics (mean and 95% CI or fixed value): 0.01 (<0.01, 0.03)

**Notes**

There is only one published study on fexinidazole, so the probability of serious adverse events will be parameterized with the observations from that study: 4 adverse events attributable to fexinidazole in 3 people among 264 people, so we have assigned a distribution of Beta(3, 261).

There were additional data reported in the appendix to the WHO's interim guidelines[24] related to studies that are ongoing. However, since we do not know details about whether those SAEs were attributable to treatment, we have chosen to omit those data.

##### S6.5.27 Days lost to disability: due to stage 1 (S1) disease

- Name in the code: `treat_effect[["yld_yll"]][["yld_s1_days]]`
- Source: [29, 30]
- Country of estimate: Uganda
- Statistical distribution and parameters: Gamma(21.89, 26.07)
- Summary statistics (mean and 95% CI or fixed value): 569.21 (355.50, 831.29)

**Notes**

Checchi and colleagues [31] reported a rate of progression from stage 1 to stage 2 disease of 0.0019 (0.0012, 0.0028), so we used the inverse of the confidence interval to come up with gamma parameters.

Our distribution is therefore Gamma(21.89, 26.07) with a mean and 95% confidence interval of 570.71 (359.18, 827.07), which provides a sufficiently large bound of uncertainty in light of the scarce information we have.

##### S6.5.28 Days lost to disability: due to stage 2 (S2) disease

- Name in the code: `treat_effect[["yld_yll"]][["yld_s2_days]]`
- Source: [30]
- Country of estimate: Uganda
- Statistical distribution and parameters: Gamma(22.18, 12.38)
- Summary statistics (mean and 95% CI or fixed value): 275.57 (172.46, 401.20)

**Notes**

Checchi and colleagues [31] reported a rate of death from stage 2 disease of 0.0040 (0.0025, 0.0058), so used the inverse of the confidence interval to come up with gamma parameters.

Our distribution is therefore Gamma(22.18, 12.38) with a mean and 95% confidence interval of 273.48 (170.99, 400.96), which provides a sufficiently large bound of uncertainty in light of the scarce information we have.

##### S6.5.29 Days lost to disability: due to a serious adverse event (SAE)

- Name in the code: `treat_effect[["yld_yll"]][["sae_yld_s1s2_days]]`
- Source: [27]
- Country of estimate: DRC
- Statistical distribution and parameters: Gamma(1.22, 2.38)
- Summary statistics (mean and 95% CI or fixed value): 2.96 (0.14, 9.99)

**Notes**

Our only source of information for the duration of severe adverse events (SAEs) is Alirol 2013 [27], which lists the most common adverse events and the median duration of these events. Most envents last a median of 1-2 days (with interquartile ranges reaching up to 4 days).

For simplicity, we have fit a gamma distribution with interquartile range of 1-4 days. Our distribution is therefore Gamma(1.22, 2.38) with a mean and 95% confidence interval of 2.92 (0.13, 9.98), which provides a sufficiently large range of values in light of the scarce information we have.

##### S6.5.30 Life-years lost (DALY) parameters

##### S6.5.31 Life expectancy

- Name in the code: `disability[["le_years"]]`
- Source: [32]
- Country of estimate: DRC
- Statistical distribution and parameters: Fixed value
- Summary statistics (mean and 95% CI or fixed value): 60.02

**Notes**

We retrieved the life expectancy at birth from the World Bank's World Development Index [32]. As this should be quite accurate we have decided to enter a fixed estimate into the model.

##### S6.5.32 Disability weights: stage 1 (S1) disease

- Name in the code: `disability[["dw_s1"]]`
- Source: [33]
- Country of estimate: GBD
- Statistical distribution and parameters: Beta(22.96, 147.21)
- Summary statistics (mean and 95% CI or fixed value): 0.14 (0.09, 0.19)

**Notes**

The Global Burden of Disease listed the impact of sleeping sickness as equivalent to the health state labeled "Motor plus cognitive impairments, severe" and their estimate for a disability weight is 0.542 (0.374-0.702), using the 2013 weight values. No distinction was made between stage 1 and 2 of the disease.

While this seems appropriate for the second stage of sleeping sickness, for stage 1 disability we chose to use the disability weights for equivalent to "infectious disease, acute episode, severe", which is described as "has a high fever and pain, and feels very weak, which causes great difficulty with daily activities" and has a much lower disability weight equivalent to 0.133 (0.088-0.190). The distribution for this parameter is therefore Beta(22.96, 147.21).

It should be noted that other cost-effectiveness analyses have used different values for disability weights [34, 35, 8]. These values arise from the 1994 Global Burden of Disease Study but we prefer to consider updated values. Since most of the disability is due to deaths rather than illness during life, we do not believe that this difference is cause for concern.

##### S6.5.33 Disability weights: stage 2 (S2) disease

- Name in the code: `disability[["dw_s2"]]`
- Source: [33]
- Country of estimate: GBD
- Statistical distribution and parameters: Beta(18.37, 15.63)
- Summary statistics (mean and 95% CI or fixed value): 0.54 (0.37, 0.70)

**Notes**

The Global Burden of Disease listed the impact of sleeping sickness as equivalent to the health state labeled "Motor plus cognitive impairments, severe" and their estimate for a disability weight is 0.542 (0.374-0.702), using the 2013 weight values. No distinction was made between stage 1 and 2 of the disease. The distribution for the parameter is Beta(18.37, 15.63).

It should be noted that other cost-effectiveness analyses have used different values for disability weights [34, 35, 8]. These values arise from the 1994 Global Burden of Disease Study but we prefer to consider updated values. Since most of the disability is due to deaths rather than illness during life, we do not believe that this difference is cause for concern.

##### S6.5.34 Disability weights: serious adverse events (SAE)

- Name in the code: `disability[["sae_dw_s1s2"]]`
- Source: [33]
- Country of estimate: GBD
- Statistical distribution and parameters: Uniform(0.04, 0.11)
- Summary statistics (mean and 95% CI or fixed value): 0.08 (0.04, 0.11)

**Notes**

As far as we are aware, no one has considered the disability due to severe adverse events attributable to gHAT treatment, but the most common adverse events are gastrointestinal problems and headaches.

We consulted the Global Burden of Disease for disability weights. The health state labeled "symptomatic tension-type headache" was described as "moderate headache that also affects the neck, which causes difficulty in daily activities" and was estimated to have a disability weight equal to 0.037 (0.022–0.057). The health state labeled "moderate symptomatic gastritis and duodenitis without anemia" was described as "abdominopelvic problem, moderate has pain in the belly and feels nauseous; the person has difficulties with daily activities" and and was estimated to have a disability weight equal to 0.114 (0.078–0.159).

Our distribution is therefore Uniform 0.037-0.114. Since most of the disability is due to deaths rather than illness during life, we do not believe that the uncertainty in this parameter is cause for concern for the purpose of the conclusions of this analysis.

###### S6.5.35 Vector control parameters

###### S6.5.36 Vector control: linear kilometers targets

- Name in the code: `vector_effect[["vc_km"]]`
- Source: [36]
- Country of estimate: DRC
- Statistical distribution and parameters: Fixed value
- Summary statistics (mean and 95% CI or fixed value): Varies by health zone

**Notes**

The parameter for the extent of riverbank covered by vector targets is a fixed value. The values for each health zone are detailed in the section S1.2.1 and sensitivity analysis were described in section S1.7.

###### S6.5.37 Vector control: units per kilometer

- Name in the code: `vector_effect[["vc_units"]]`
- Source: [36]
- Country of estimate: DRC and Uganda
- Statistical distribution and parameters: Fixed value
- Summary statistics (mean and 95% CI or fixed value): Varies by health zone

**Notes**

We used a fixed value for the parameter for the extent of riverbank covered by vector targets. The values for each health zone are detailed in the section S1.2.1 and sensitivity analysis were described in section S1.7.

###### S6.5.38 Vector control: replacement rate of each unit

- Name in the code: `vector_effect[["vc_deployments_yr"]]`
- Source: [36, 37]
- Country of estimate: DRC
- Statistical distribution and parameters: Fixed value
- Summary statistics (mean and 95% CI or fixed value): 2

**Notes**

We have set this parameter as a fixed number, as this is the number of times that one must replace a set of targets in order to provide continuous protection throughout the year [36, 37].

#### S6.6 Notes on cost parameters

###### S6.6.1 Surveillance cost parameters

###### S6.6.2 Active screening: capital costs of a traditional team

- Name in the code: `surv_cost[["as_capital"]]`
- Source: [7–9]
- Country of estimate: DRC: Mosango and Yasa Bonga
- Distribution and parameters: Gamma(81.02, 114.54)
- Summary statistics (mean and 95% CI or fixed value): 9,276 (7,378, 11,375)

**Notes**

Capital costs are denominated in 2018 US dollars.

To our knowledge, no active surveillance costs have been estimated via detailed costing study [38]. We took into account only the two studies that calculated costs using an ingredients approach [7, 8].

Lutumba and colleagues [7] calculated that the total cost of screening 40,000 patients in 2003 was 46,734.29 Euros (1 Euro = 0.86 USD in 2003) (see table 3 of [7]). Of that value, 21 percent of the costs were capital costs, or a cost of 8216.99 in 2018 USD. The publication did not indicate whether that value was annualized or not.

According to Bessel and colleagues [8] the cost for a mobile team that screens 250 patients per day for 220 days a year has capital investments (annualized for five years) of $12,000 for a team that administers CATT and $12,781 for a team that administers RDT (in 2013 USD) (see [8] Table S1). Adjusting for inflation, that would equal 7369.22 in 2018 USD.

According to Snijders et al [9], the capital cost of an active surveillance team was $11,406 in 2018 for a team that administer CATT.

Although no publication gave us a sense of the uncertainty in capital costs, but since we had three studies, we assumed that the range was equivalent to the 95% confidence interval. Therefore, our probability distribution is Gamma(81.02, 114.54), which yields a distribution with mean and confidence intervals of 9,282 (7,342, 11,417).

##### S6.6.3 Active screening: fixed management costs of a traditional team

- Name in the code: `surv_effect[["as_recurrent"]]`
- Source: [7–9]
- Country of estimate: DRC: Mosango and Yasa Bonga
- Distribution and parameters: Gamma(63.31, 630.94)
- Summary statistics (mean and 95% CI or fixed value): 39,955 (30,845, 50,435)

**Notes**

Recurrent management costs are denominated in 2018 US dollars.

To our knowledge, no active surveillance costs have been estimated via detailed costing study [38]. We took into account only the two studies that calculated costs using an ingredients approach [7–9].

Lutumba and colleagues [7] calculated that the total cost of screening 40,000 patients in 2003 was 46,734.29 Euros (1 Euro = 0.86 USD in 2003, for an equivalent of $40,191) (see table 3 of [7]). Of that value, 79 percent of the costs were recurrent fixed costs ($31,751), or a cost of 30,911.52 in 2018 USD after adjusting for inflation and changes in the exchange rate.

According to Bessel and colleagues [8] the cost for a mobile team that screens 250 patients per day for 220 days a year has annual recurrent costs of $30,307 and daily recurrent costs of $97 in 2013 values (see [8] Table S1). Summing those costs ($51,647) and adjusting for inflation and changes in the exchange rate, that would equal 30,717.00 in 2018 USD.

According to Snijders and colleagues [9], the management and recurrent costs of an active surveillance team were $42,408 in 2018, and $7,961 for the management costs from the provincial and central level PNLTHA (including training and supervision) for a total of 50,369.

Although no publication gave us a sense of the uncertainty in recurrent costs, but since we had at least three studies, we assumed that the range of observations was equal to the 95% confidence interval (30,717- 50,359). Therefore, our probability distribution is Gamma(63.31, 630.94), which yields a distribution with mean and confidence intervals of 39,951 (30,969, 50,618).

##### S6.6.4 CATT algorithm: cost per test used

- Name in the code: `surv_cost[["dx_catt"]]`
- Source: [7–9, 38]
- Country of estimate: DRC
- Distribution and parameters: Gamma(25.19, 0.02)
- Summary statistics (mean and 95% CI or fixed value): 0.52 (0.34, 0.75)

**Notes**

To our knowledge, costs for CATT tests are only featured in three sources: 1) the WHO, cited by Keating et al 2015 [38], 2) by Lutumba et al [39], and 3) by Bessel et al [8].

1) Keating et al listed the cost of a CATT test at 0.73 in 1998 USD, equivalent to 0.34 in 2018 USD.

2) Lutumba et al listed the cost of a CATT test at 0.52 in 2003 USD, equivalent to 0.37 in 2018 USD.

3) Bessel et al listed the cost of CATT test at 0.70 for a mobile team and 0.89 for a fixed post in 2013 USD. The reason for the difference in price between screening at a mobile team vs screening at a fixed post was due to the differential wastage and was given in the supplement notes: "Costs at mobile teams and fixed units are different because once a bottle of CATT antigen is open repeat cases and controls must be performed. Materials are the cost of test materials plus the cost of the lancet."

In terms of 2018 USD, the costs are 0.42 for CATT tests in mobile teams 0.53 for CATT tests in fixed posts. We take into account wastage as separate parameter.

4) Snijders et al listed the cost of a CATT test at 0.74 in terms of 2018 USD [9].

Because we had more than two studies on this parameter, we chose gamma distributions where the 2.5th and the 97.5th percentiles would match the minimum and maximum values in the literature, so our parameter distribution is Gamma(25.19, 0.02), which has a mean and confidence interval of 0.52 (0.34, 0.74).

Considering delivery costs (described in S6.6.17), the cost of CATT tests are 0.73 (0.47, 1.05) in 2018 USD. Considering both delivery costs and wastage, the cost of CATT tests are 0.81 (0.51, 1.16) in active screening teams and 0.94 (0.60, 1.36) in passive surveillance (fixed) posts.

###### S6.6.5 Lumbar puncture and laboratory exam: cost per person that needs to be staged

- Name in the code: `surv_cost[["dx_lp_lab"]]`
- Source: [9, 40]
- Country of estimate: DRC and Chad
- Distribution and parameters: Gamma(2.42, 3.66)
- Summary statistics (mean and 95% CI or fixed value): 8.90 (1.45, 23.20)

**Notes**

To our knowledge, costs for lumbar puncture tests were listed in detail only by Bessel and colleagues [8] and will be featured as part of an upcoming publication by Snijders and colleagues [9]. The cost listed by Bessel et al was 2.38 in terms of 2013 USD, equivalent to 1.42 in 2018 USD. Snijders and colleagues report a cost of 23.02 in 2018 USD. Irurzun-Lopez [40] have reported a similar value, so we will assume a value equal to that of Snijders and colleages.

Because we had more than two studies to inform this parameter, we chose gamma distributions where the 2.5th and the 97.5th percentiles would match the minimum and maximum values in the literature, so our parameter distribution is Gamma(2.42, 3.66), which yields a distribution with mean and confidence intervals of 8.83 (1.45, 22.94).

###### S6.6.6 Microscopy (Blood sample, LNA, mAECT): cost per person that needs to be confirmed

- Name in the code: `surv_cost[["dx_microscopy1"]]`
- Source: [12]
- Country of estimate: DRC: Mosango and Yasa Bonga
- Distribution and parameters: Gamma(8.47, 1.27)
- Summary statistics (mean and 95% CI or fixed value): 10.68 (4.70, 18.84)

**Notes**

The per-patient price to confirm a patient with a full microscopy procedure is reported in an upcoming publication by Snijders and colleagues. They report that a microscopy procedure constituted of mAECT and LNA (lymph node aspiration) costs 9.53 in 2018 USD. Because we had no report of the standard error around that estimate, we assigned a gamma distribution that had a confidence interval that spanned half the estimate and twice the estimate, yielding a distribution of Gamma(8.47, 1.27).

###### S6.6.7 RDT algorithm: costs per test used

- Name in the code: `surv_cost[["dx_rdt"]]`
- Source: [12]

- Country of estimate: DRC
- Distribution and parameters: Gamma(8.47, 0.19)
- Summary statistics (mean and 95% CI or fixed value): 1.60 (0.71, 2.83)

**Notes**

To our knowledge, costs for RDT tests were listed in Sutherland et al [35], Bessel et al [8], and an upcoming publication by Snijders and colleagues [12].

1) Both Sutherland et. al and Bessel et. al coincided on a cost of 0.50 USD 2013 for the test in the international market (after a 25 cent subsidy paid for outside of DRC). Bessel also calculated staff costs, shipment, and wastage for 0.32 USD 2013. Because we can split the cost between tradable and non-tradable costs, we inflate and adjust the costs for the staff costs and shipment and then add the cost of the RDT. In terms of 2018 USD costs, the shipment and the staff costs are 0.19, and the total cost is 0.94.
2) Snijders and colleagues reported a cost between 0.85 and 1.97, depending on the company from which the test is purchased. It should be noted that no single company produces enough tests for any single intervention, and so a range of prices must be considered. Snijders' estimate does not include delivery or wastage, so we take that into account separately.

It appears difficult to compare the three estimates, so we will take Snijders' estimate from a recent micro-costing analysis. The mean was $1.58, and we assign a distribution with confidence intervals equal to half and double the costs. The distribution is Gamma(8.47, 0.19).

Considering delivery costs (described in S6.6.17), the cost of RDT tests are 2.30 (1.01, 4.12) in 2018 USD. Considering both delivery costs and wastage, the cost is 2.46 (1.08, 4.39) in passive surveillance teams, and we don't consider the deployment of RDTs in active surveillance teams.

##### S6.6.8 Variable management costs (PNLTHA mark-up)

- Name in the code: `surv_cost[["pnltha_mu"]]`
- Source: [12]
- Country of estimate: DRC
- Distribution and parameters: Uniform(0.1, 0.2)
- Summary statistics (mean and 95% CI or fixed value): 0.15 (0.10, 0.20)

**Notes**

Snijder's and colleagues [12] have assumed that there is a component of management at the national program level that is approximately 15% of the expenses at the local and coordination level (both fixed costs and variable/consumable costs for both active screening and passive surveillance in fixed health posts). However, the mark-up was not applied to the consult in the fixed health post, as these were consultations paid for by patients for symptoms in general, but testing and confirmation for HAT specifically is administered and paid for by PNLTHA, so we have included a mark-up for these items.

##### S6.6.9 Passive surveillance: capital costs of a facility

- Name in the code: `surv_effect[["ps_capital"]]`
- Source: [12]
- Country of estimate: DRC: Mosango and Yasa Bonga
- Distribution and parameters: Gamma(8.47, 209.8)
- Summary statistics (mean and 95% CI or fixed value): 1,777 (778, 3,157)

**Notes**

Capital costs are denominated in 2018 US dollars and apply to each health center or hospital that is capable of HAT diagnosis.

To our knowledge, no passive surveillance costs have been estimated via detailed costing study [38]. We took into account the upcoming results of a micro-costing study in Yasa Bonga and Mosango [9], two health zones of the former Bandundu province and in the current study.

To parameterize the model, we assign a distribution with 95% confidence intervals equal to half and double the costs. The distribution is Gamma(8.47, 209.80), which yields a distribution with median and confidence intervals of 1,705 (780, 3,151).

###### S6.6.10 Passive surveillance: fixed recurrent management costs

- Name in the code: `surv_effect[["ps_recurrent"]]`
- Source: [12]
- Country of estimate: DRC: Mosango and Yasa Bonga
- Distribution and parameters: Gamma(8.47, 985.55)
- Summary statistics (mean and 95% CI or fixed value): 8,368 (3,743, 14,965)

**Notes**

Recurrent management costs are denominated in 2018 USD and apply to health health zone.

To our knowledge, no passive surveillance costs have been estimated via detailed costing study [38]. We took into account the upcoming results of a micro-costing study in Yasa Bonga and Mosango [12], two health zones of the former Bandundu province and two health zones of the current study. The value was 7422 in 2018 USD.

To parameterize the model, we assign a distribution with confidence intervals equal to half and double the costs. The distribution is Gamma(8.47, 985.55), which yields a distribution with median and confidence intervals of 8,051 (3,802, 14,830).

###### S6.6.11 Treatment cost parameters

###### S6.6.12 Hospital stay: cost per day

- Name in the code: `treat_cost[["ip_day"]]`
- Source: [1, 41, 42]
- Country of estimate: DRC
- Distribution and parameters: Gamma(5.81, 0.24)
- Summary statistics (mean and 95% CI or fixed value): 1.39 (0.50, 2.71)

**Notes**

We got the estimates of inpatient treatment costs from the 2010 WHO CHOICE cost estimates (recently updated by [41] and [42]). In 2010, a consult at a primary hospital in DRC would be 2.41 (0.90, 5.73) I$. In 2010, a consult at a secondary hospital in DRC would be 2.59 (0.98, 5.81) I$, and a consult at a tertiary hospital in DRC would be 3.25 (1.32, 7.20) I$.

The equivalent estimates in 2010 USD are 1.27 (0.47, 3.02) per day at a primary hospital, 1.36 (0.52, 3.06) per day at a secondary hospital, and 1.71 (0.70, 3.79) per day at a tertiary hospital. After converting to local currency, applying the inflation index, and converting to 2018 USD, the estimates are 0.95 (0.35, 2.26) per day at a primary hospital, 1.02 (0.39, 2.29) per day at a secondary hospital, and 1.28 (0.52, 2.84) per day at a tertiary hospital.

At the moment, we do not know how many of each kind of hospital the population of HAT patients attend, nor do we understand how costs at district hospitals, referral hospitals, etc resemble those of the two kinds of hospitals under analysis by the WHO CHOICE program. Therefore, we take the estimate with a higher mean (hospital day in a tertiary hospital) in an effort not to under-state the costs of treatment and interventions.

To parameterize the model, we assign a gamma distribution with 95% confidence intervals equal to those reported by WHO CHOICE [1]. The distribution is Gamma(5.81, 0.25), which yields a distribution with median and confidence intervals of 1.45 (0.52, 2.85). Although this yields a higher mean, the uncertainty is adequately characterized.

###### S6.6.13 Outpatient consultation: cost

- Name in the code: `treat_cost[["op"]]`
- Source: [12, 43]
- Country of estimate: DRC
- Distribution and parameters: Uniform(1.37, 3.33)
- Summary statistics (mean and 95% CI or fixed value): 2.34 (1.42, 3.28)

**Notes**

We got the estimates of outpatient consultation costs from two sources:

1) Laokri and colleagues [43] presented an estimate with mean $2.33 and standard deviation 0.27 in 2013 values, or 1.38 in 2018 values.
2) Snijders and colleagues [12] reported that the cost of a consultation is $3.33 in 2018 values.

To parameterize the model, we assign a distribution: Uniform(1.37, 3.33).

###### S6.6.14 Course of pentamidine: cost

- Name in the code: `treat_cost[["rx_pentamidine"]]`
- Source: [38]
- Country of estimate: WHO
- Distribution and parameters: Fixed value
- Summary statistics (mean and 95% CI or fixed value): 54

**Notes**

The cost of pentamidine, for stage 1 disease. Because it is available on the international market, where is it sold in USD, and not subject to the inflationary pressures of any particular country, we have not inflated the cost or converted them to any other currency.

In the future pentamidine treatment it may be replaced with fexinidozole treatment, which would circumvent the need for a lumbar puncture.

###### S6.6.15 Course of nifurtimox eflornithine combination therapy (NECT): cost

- Name in the code: `treat_cost[["rx_nect"]]`
- Source: [44]
- Country of estimate: WHO
- Distribution and parameters: Fixed value
- Summary statistics (mean and 95% CI or fixed value): 460

**Notes**

This represents the cost of NECT to the capital for stage 2 disease. Simarro and colleagues listed a cost of 1440 USD for treatment of four patients.

Because it is available on the international market, where is it sold in USD, and not subject to the inflationary pressures of any particular country, we have not inflated the cost or converted them to any other currency.

In the future it may be replaced with fexinidozole and this would be the drug for treatment failures or very severe patients.

###### S6.6.16 Course of fexinidazole: cost

- Name in the code: `treat_cost[["rx_fexinidazole"]]`
- Source: [35]
- Country of estimate: WHO
- Distribution and parameters: Fixed value
- Summary statistics (mean and 95% CI or fixed value): 50

**Notes**

The cost of fexanidozole, for stage 1 and 2 disease. In the near future this will be the drug of choice for first-line treatment for both stages of disease. It may require hospitalization, but eventually it should be taken on an outpatient basis.

##### S6.6.17 Drug delivery mark-up

- Name in the code: `treat_cost[["rx_delivery"]]`
- Source: [41, 42]
- Country of estimate: DRC
- Distribution and parameters: Beta(45, 55)
- Summary statistics (mean and 95% CI or fixed value): 0.45 (0.35, 0.55)

**Notes**

Because we do not know the delivery price of drugs for each country, we have applied the standard value for the mark up of traded goods recommended by the WHO CHOICE program for AFRO E: `https://www.who.int/choice/cost-effectiveness/inputs/price_multiplier/en/`.

##### S6.6.18 Vector cost parameters

##### S6.6.19 Vector control: operational cost per kilometer of riverbank covered

- Name in the code: `vector_cost[["vc_operation"]]`
- Source: [45]
- Country of estimate: Uganda
- Distribution and parameters: Gamma(8.47, 14.17)
- Summary statistics (mean and 95% CI or fixed value): 120.28 (53.33, 212.26)

**Notes**

Vector control operational costs are denominated in 2018 US dollars on a per-kilometer basis.

To our knowledge, only one vector control micro-costing study has been performed by Shaw and colleagues [45]. In that study, centered in Arua, Uganda, 1551 targets were laid out at 20 targets per kilometer on the river banks [46], for a total of 77.55 kilometers. The total cost of vector control operations for one year was 21,337 in 2013 USD, but it included a deployment and a target maintenance mission for 4,290 USD. Further, the target deployment itself cost 7370 USD. Therefore, the cost of the operation excluding target deployment was 9,677 USD per year, or 124.79 USD per kilometer in 2013 USD in Uganda.

To convert those Uganda costs to DRC costs in 2018 (as most costs were in non-tradeable goods and labor), we turn the USD into Uganda's local currency in 2013. We can take Uganda's local currency and convert that value to PPP (international dollars) values using the PPP conversion factor (local currency units per international dollar) for Uganda. Then we took the operational costs in international dollars and convert them to local currency in DRC with the PPP conversion factor for DRC. Next, we use the CPI to inflate costs to 2018 levels and then use the exchange rate with USD to get 2018 USD values. The equivalent cost for DRC is 106.70 USD per kilometer per year.

As there was no sense of the uncertainty in target deployment costs, we assign a distribution with confidence intervals equal to half and double the costs. The distribution is Gamma(8.47, 14.17), which yields a distribution with mean and confidence intervals of 120.59 (53.15, 213.98).

##### S6.6.20 Vector control: deployment cost per target

- Name in the code: `vector_cost[["vc_deploy"]]`
- Source: [45]
- Country of estimate: Uganda
- Distribution and parameters: Gamma(8.47, 0.54)
- Summary statistics (mean and 95% CI or fixed value): 4.57 (2.02, 8.26)

**Notes**

Target deployment costs are denominated in 2018 US dollars on a per-target basis.

To our knowledge, only one vector control micro-costing study has been performed by Shaw and colleagues [45]. In that study, centered in Arua, Uganda, 1,551 targets were laid out. The target deployment activities cost 7,370 USD, or per target 4.75 USD (in 2013 values).

To convert those Uganda costs to DRC costs in 2018 (as most costs were in non-tradeable goods and labor), we turn the USD into Uganda's local currency in 2013. We can take Uganda's local currency and convert that value to PPP (international dollars) values using the PPP conversion factor (local currency units per international dollar) for Uganda. Then we took the operational costs in international dollars and convert them to local currency in DRC with the PPP conversion factor for DRC. Then, we use the CPI to inflate costs to 2018 levels and then use the exchange rate with USD to get 2018 USD values. The equivalent cost for DRC is 4.06 USD per target.

Uncertainty: as there was no sense of the uncertainty in target deployment costs, we assign a distribution with confidence intervals equal to half and double the costs. The distribution is Gamma(8.47, 0.54), which yields a distribution with mean and confidence intervals of 4.57 (2.07, 8.22).
